# Single nucleus RNA-sequencing reveals altered intercellular communication and dendritic cell activation in nonobstructive hypertrophic cardiomyopathy

**DOI:** 10.1101/2021.12.20.21267954

**Authors:** Christina J. Codden, Amy Larson, Junya Awata, Gayani Perera, Michael T. Chin

**Author notes:** To whom correspondence should be addressed Michael T. Chin, MD, PhD, Molecular Cardiology Research Institute, Tufts Medical Center, 800 Washington Street, Box 80, Boston, MA 02111.

## Abstract

End stage, nonobstructive hypertrophic cardiomyopathy (HCM) is an intractable condition with no disease-specific therapies. To gain insights into the pathogenesis of nonobstructive HCM, we performed single nucleus RNA-sequencing (snRNA-seq) on human HCM hearts explanted at the time of cardiac transplantation and organ donor hearts serving as controls. Differential gene expression analysis revealed 64 differentially expressed genes linked to specific cell types and molecular functions. Analysis of ligand-receptor pair gene expression to delineate potential intercellular communication revealed significant reductions in expressed ligand-receptor pairs affecting the extracellular matrix, growth factor binding, peptidase regulator activity, platelet-derived growth factor binding and protease binding in the HCM tissue. Changes in Integrin-β1 receptor expression were responsible for many changes related to extracellular matrix interactions, by increasing in dendritic, smooth muscle and pericyte cells while decreasing in endothelial and fibroblast cells, suggesting potential mechanisms for fibrosis and microvascular disease in HCM and a potential role for dendritic cells. In contrast, there was an increase in ligand-receptor pair expression associated with adenylate cyclase binding, calcium channel molecular functions, channel inhibitor activity, ion channel inhibitor activity, phosphatase activator activity, protein kinase activator activity and titin binding, suggesting important shifts in various signaling cascades in nonobstructive, end stage HCM.

**Brief summary:** End stage, nonobstructive human HCM is associated with altered intercellular communication and dendritic cell activation, providing novel insights into potential disease mechanisms.

## Introduction

Hypertrophic Cardiomyopathy (HCM) is an inherited disorder characterized by unexplained left ventricular hypertrophy, often asymmetric, often involving the interventricular septum, often associated with left ventricular outflow tract (LVOT) obstruction, fibrosis, microvascular occlusion, and sudden cardiac death. Genetic studies have identified numerous causal mutations in a variety of sarcomere genes such as *MYH7, MYL2, MYL3, MYBPC3, TNNT2, TNNI3* and *TPM1*, leading to the concept that HCM is a disease of the sarcomere ^1^, but genetic testing is only informative in approximately 30% of probands, indicating that other genes and factors significantly contribute to the HCM phenotype. Recently, it has been recognized that in most cases HCM can be considered polygenic with multiple loci contributing to the phenotype or in some cases acting as modifiers of existing sarcomere mutations, by affecting modifiable risk factors such as diastolic blood pressure ^2–4^. Patients with sarcomere gene mutations have been found to have more adverse events than those without sarcomere mutations ^5^, thus implicating a need for better understanding of distinct pathogenic steps in these two groups. The development of fibrosis is associated with an increased risk of sudden cardiac death and is a poor prognostic factor ^6–11^. Excess deposition of extracellular matrix (ECM) proteins can cause cardiac stiffening, which can impede contraction ^6^. It can additionally disrupt electrical coupling, heightening patient susceptibility to arrhythmogenesis. In the end-stages of disease, fibrosis replaces up to 50% of the myocardium and is a key determinant of patient outcome ^9–11^.

Although LVOT obstruction is common and easily treatable in HCM, a subset of patients with nonobstructive HCM develop progressive, symptomatic heart failure, despite guideline directed medical therapy, often leading to heart transplantation ^12^. Histopathological features include myocyte hypertrophy, fibrosis, myocyte disarray and microvascular disease and are common with obstructive HCM. The genetic profiles of patients with nonobstructive HCM are also indistinguishable from those with obstructive HCM, raising questions about the sources of asymmetric hypertrophy. Human tissue for study of nonobstructive HCM can only be obtained postmortem or at the time of heart transplantation or other cardiac procedure.

Single cell and single nucleus transcriptomic analysis have facilitated an understanding of cellular phenotypes and interactions occurring in complex tissues such as the heart and high quality datasets of the normal human heart have been published ^13–15^. We have recently performed single nucleus RNA-seq analysis of obstructive HCM and found significant alterations in intercellular communication pathways that affect the extracellular matrix, involving integrin-β1, thus providing a potential mechanism linking sarcomere dysfunction to extracellular matrix signaling ^16^. Here, we analyze single nucleus transcriptomes in tissue from nonobstructive, end stage HCM and compare with normal tissue. As with obstructive HCM, we find that alterations in intercellular communication pathways affecting the extracellular matrix involve integrin-β1, but there are additional alterations in communication between fibroblasts, vascular cells, and dendritic cells not seen in obstructive HCM. In addition, there are increases in various molecular functions associated with adenylate cyclase signaling, ion channel function, protein phosphatase and protein kinase activity, suggesting a unique intercellular signaling milieu specific to nonobstructive HCM. These findings have important implications for the development of novel therapies for nonobstructive HCM.

## Results

### Patient characteristics

Explanted heart tissue was obtained from six HCM patients with severely symptomatic, nonobstructive, end stage disease who underwent cardiac transplantation. Patient characteristics are summarized in Table 1. Patients gave informed consent for their heart tissue to be used in research. The patients varied in age from 25 to 67. Five of six patients were female. Two of the patients carried pathogenic MYH7 mutations and the remaining four patients had no known mutations pathogenic for HCM. Four organ donors and datasets for the normal human IVS have been previously described ^15^ and are summarized in Table 2 with the two additional donors used in this study.

**Table 1.** HCM Patient Characteristics

| Patient | 2833 | 2890 | 2891 | 2918 | 2930 | 5171 |
| --- | --- | --- | --- | --- | --- | --- |
| Demographics |  |  |  |  |  |  |
| age at transplant | 21-25 | 61-65 | 46-50 | 61-65 | 36-40 | 66-70 |
| female | yes | yes | no | yes | yes | yes |
| nyha class>3 | yes | yes | yes | yes | yes | yes |
| Med Hx |  |  |  |  |  |  |
| Prior AF | no | yes | yes | yes | yes | no |
| Prior VT/VF | yes | no | no | no | no | yes |
| Prior NS VT | no | no | no | no | no | yes |
| Prior syncope | yes | no | yes | no | no | no |
| Fam Hx SCD | no | no | yes | yes | no | yes |
| Fam Hx HCM | no | no | yes | yes | yes | yes |
| Comorbidities | none | HTN, HLD, COPD, DM2, CHB, OA, prior myectomy | OSA, Sarcoidosis | CAD, hypothyroidism | hypothyroidism, apical hypertrophy | asthma, HLD, apical aneurysm |
| Meds |  |  |  |  |  |  |
| beta blocker | yes | yes | no | yes | no | yes |
| calcium channel blocker | no | no | no | no | no | no |
| ACE or ARB | no | no | no | yes | no | yes |
| Diuretic Use | yes | yes | yes | yes | yes | yes |
| loop diuretic | yes | yes | yes | yes | yes | yes |
| thiazide | no | yes | yes | no | no | yes |
| potassium sparing | yes | yes | yes | yes | yes | yes |
| disopyramide | no | no | no | no | no | no |
| amiodarone | no | no | no | no | yes | no |
| milrinone | yes | yes | yes | yes | no | yes |
| dofetilide | no | no | yes | no | no | no |
| ICD | yes | yes | yes | yes | yes | yes |
| Physiological measurements |  |  |  |  |  |  |
| LA size (mm) | 47 | 37 | 36 | 64 | 33 | 51 |
| systolic blood pressure | 95 | 103 | 120 | 87 | 99 | 130 |
| diastolic blood pressure | 57 | 56 | 54 | 66 | 49 | 67 |
| IVS thickness (mm) | 26 | 11 (prior myectomy) | 13 | 18 | 9.5 | 9 |
| Posterior wall thickness | 20 | 0.88 | 13 | 16 | 7.7 | 9 |
| LVEF (%) | 15-20 | 65 | 60 | 45 | 65 | 25-30 |
| LVEDD (mm) | 51 | 46 | 45 | 41 | 45 | 57 |
| LVESD (m) | 49 | 29 | 28 | 35 | 49 | 41 |
| MR | mild | trace | no | trace | mild | mild |
| LGE on MRI | 24% | none | ND | ND | none | ND |
| Pathogenic HCM Variant | NF | NF | MYH7 | NF | NF | MYH7 |

**Table 2.** Organ Donor Characteristics

| Donor | 2867 | 2879 | 2880 | 2884 | 2900 | 3045 |
| --- | --- | --- | --- | --- | --- | --- |
| age | 21-25 | 56-60 | 41-45 | 41-45 | 46-50 | 66-70 |
| female | yes | no | no | yes | no | yes |
| cause of death | asphyxiation | CVA | head trauma | seizure | overdose | CVA |
| cardiac risk factors | none | none | none | HTN, Tobacco | none | HLD |
| CAD | not assessed | 20% LAD | not assessed | not assessed | not assessed | not assessed |
| LVEF | 81% | 60% | not assessed | 55% | not assessed | 70% |
| Medications | antidepressants, OCP | none | unknown | unknown | unknown | statin, levothyroxine |

### Nonobstructive HCM IVS tissue reveals extensive cardiomyocyte and fibroblast diversity

After sequencing and initial data processing with Cell Ranger software ^17^, each sample dataset was processed further to remove called nuclei that were likely droplets with ambient RNA, or droplets that contained two nuclei. The six HCM datasets and six donor heart datasets were combined into one dataset using the Seurat Integration function ^18^. The final combined dataset included 49010 nuclei from nonobstructive HCM hearts and 34358 nuclei from donor hearts. Clustering of the integrated dataset revealed 22 cell populations within the IVS which was visualized using the dimensionality reduction algorithm uniform manifold approximation and projection (UMAP, Fig. 1A). Each point represents a single nucleus colored by cluster identity. Visualizing the integrated dataset by Normal and HCM datasets reveals that all 22 clusters are present in each condition and all samples contribute to all clusters (Fig. 1B, C, D). No cell populations (i.e., clusters) were specific to either condition.

**Fig. 1.**
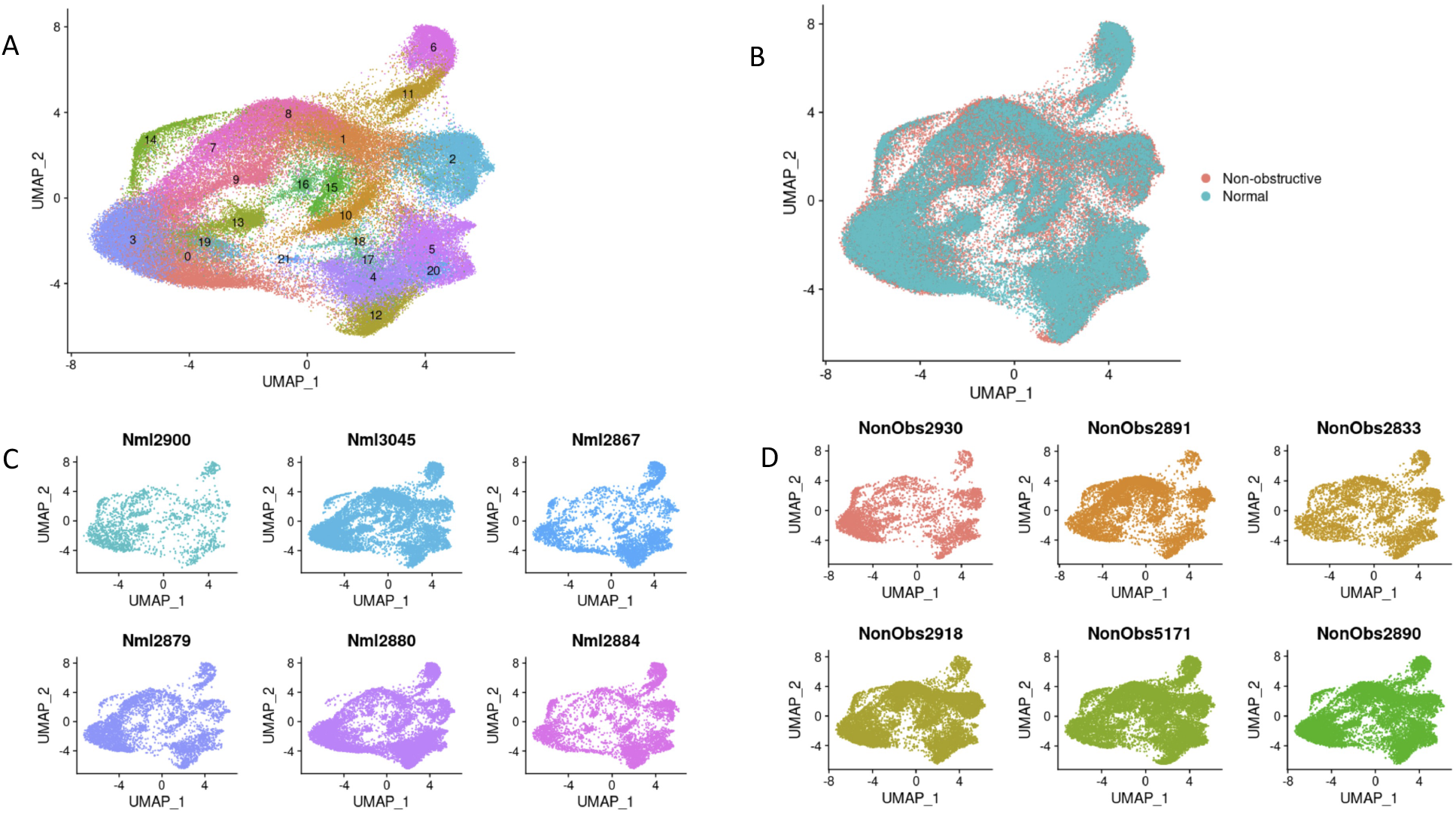
Single nuclei RNA-seq of Normal and HCM IVS Tissue Reveals Cellular Diversity but No HCM-associated Cell Types. **A**. UMAP plot of 22 distinct cell clusters identified in the combined Normal and nonobstructive HCM dataset. **B**. Separating the cell clusters by condition does not identify HCM-specific cell types. **C**. Cluster distribution in each Normal sample. D. Cluster distribution in each nonobstructive HCM sample.

Cell identities were assigned to each cluster using known gene markers of expected cell types, differentially expressed genes queried against panglaoDB ^19^, gene ontology (GO) using GOstats ^20^, and Ingenuity Pathway Analysis ^21^. Supplemental Table 1 lists the gene markers used to identify each cell type. Supplemental Table 2 lists the consensus cell identity assignment using all four methods. Upon assigning cell types, we found that similar cell types expressed similar markers and the associated clusters were expectedly positioned close to each other in UMAP space (Fig. 2A, B). Interestingly, we see ten cardiomyocyte populations (i.e., clusters) and six fibroblast populations, revealing significant cardiomyocyte and fibroblast diversity in the IVS (Fig. 2A, 2B), consistent with previous reports ^15, 16^. Other cell types identified included endothelial, smooth muscle, pericyte, neuronal, leukocyte, and dendritic cell populations. Among normal donor heart samples, assigned cardiomyocytes made up approximately 48.5% of the total cell population. Similarly, among nonobstructive HCM samples cardiomyocytes made up approximately 55.0% of the total cell population. Noncardiomyocytes make up the remaining cell population. Nonobstructive HCM does not appear to be associated with a shift in the relative proportion of cells in the heart.

**Fig. 2.**
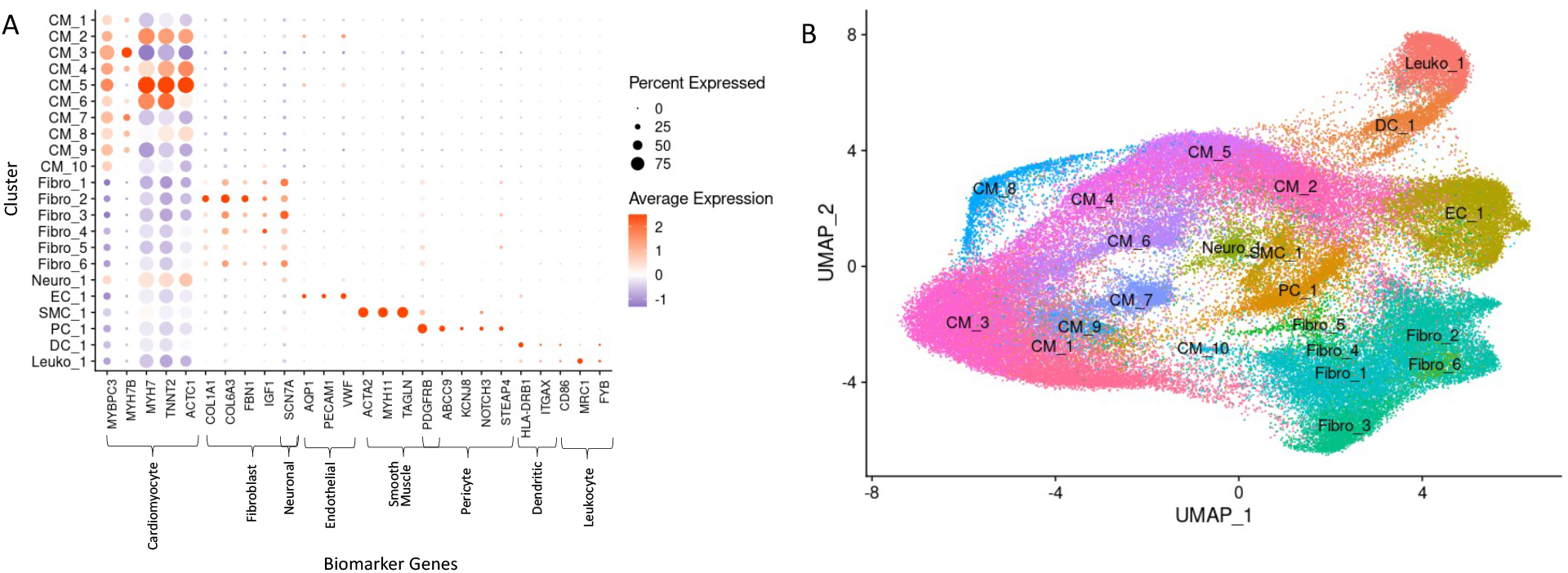
Biomarker Expression and Cell Identification Assignments for each Cluster. **A**. Biomarkers used to assign cell identity and their relative expression in each cluster. **B**. UMAP plot of IVS tissue clusters with cell assignment labels.

### Trajectory analysis and differential gene expression analysis reveal nonobstructive HCM-associated perturbations

Clustering analysis revealed diversity of both cardiomyocytes and fibroblasts. To gain insight into the potential relationships among the different cell populations for each cell type, clusters were projected onto pseudotime trajectory analysis using Monocle3 ^22^. Trajectory analysis was performed on Normal cells only, nonobstructive HCM cells only, and both conditions combined for each of the 8 cell types identified in the above analysis. In single-cell trajectory analysis, a trajectory is a computed path that describes a cell type’s biological progression through a dynamic process. Prior to building trajectory paths, the root node, or beginning, of each trajectory path was determined through hierarchical clustering of our single nuclei data by cell type. Then, trajectory paths for each assigned cell type (assigned in Seurat) were constructed in UMAP space using Monocle3 with Normal and HCM data together and individually. Once trajectory paths were established, pseudotime could be assigned to each cell. For all cell types, root nodes showed consistent placement among Normal only groups, HCM only groups, and Normal and HCM groups. Trajectory paths also showed similar basic structures among Normal only groups, HCM only groups, and Normal and HCM group (data not shown). Therefore, we were not able to distinguish any meaningful changes among trajectory paths. These findings suggest that the relationships between the various subtypes of each cell type do not vary significantly in nonobstructive HCM heart tissue in comparison to Normal heart tissue, as we have reported for obstructive HCM ^16^.

Establishment of trajectories facilitates the analysis of differential gene expression along the trajectory and between conditions along the trajectory. We analyzed differential gene expression for each cell type along their trajectories for the Normal and HCM populations. Differential expression analysis between Normal and HCM cells by assigned cell type and cluster was performed in which generalized linear regression models were fit for each gene. Resulting coefficients were tested for significant differences from zero, with an adjusted p-value cutoff ≤0.05 using the Wald Test. No differentially expressed genes were detected by this method.

Differential gene expression can also be analyzed along trajectory paths in UMAP space through spatial autocorrelation. Moran’s I statistic and adjusted p-values were calculated for each gene in Normal only and HCM only groups within each cell type. When paired with a significant p-value (≤0.05), a Moran’s I statistic value near zero indicated no spatial autocorrelation and a value near 1 indicated perfect positive autocorrelation in which a gene is expressed in a focal region of the UMAP space. Results showed between 115 and 6695 genes are differentially expressed along trajectory paths among cell types and between 18 and 4518 differentially expressed genes overlapped between Normal and HCM groups among cell types (Supplemental Table 3).

Since many genes showed differential expression over UMAP space, further conservative filtering was performed to identify genes of interest. Genes with Moran’s I statistic available for a single condition, Normal or HCM, were filtered by a Moran’s I statistic value above 0.1, while genes with Moran’s I statistics available for both classes were filtered by an absolute difference >0.1. For each cell type, 1 to 18 genes passed the filter with 64 unique genes in total passing the filter among all cell types (Supplemental Table 4). The expression of these filtered genes was then plotted in UMAP space for their associated cell type in Normal and HCM conditions as shown for a subset of identified genes in cardiomyocytes (Supplemental Figure 1). Visual analysis of the UMAP plots revealed 44 genes with pronounced differences in their spatial expression between Normal and nonobstructive HCM conditions (summary in Table 3). Of these 44 genes, 3 (*MYH7, MYL2, TNNT2*) are already known to be associated with human HCM. GO Enrichment analysis of these 44 differentially expressed genes revealed significant enrichment for molecular functions involving actin binding, biological processes involving muscle development and muscle differentiation and cellular components including the sarcomere, myofibril and contractile fiber (Fig. 3A). Analysis of genes from this list showing increased expression in nonobstructive HCM revealed significant enrichment in molecular functions involving structural constituent of muscle and actin binding, no enriched biological processes and enrichment in cellular components involving the sarcomere, myofibril and contractile fiber (Fig. 3B). Analysis of genes from this list showing decreased expression in nonobstructive HCM revealed enrichment for molecular functions involving lipid transport and the ECM structural constituent, biological processes involving muscle differentiation, muscle development, lipid transport and lipid localization, and cellular components involving the endoplasmic reticulum membrane (Fig. 3C).

**Fig. 3.**
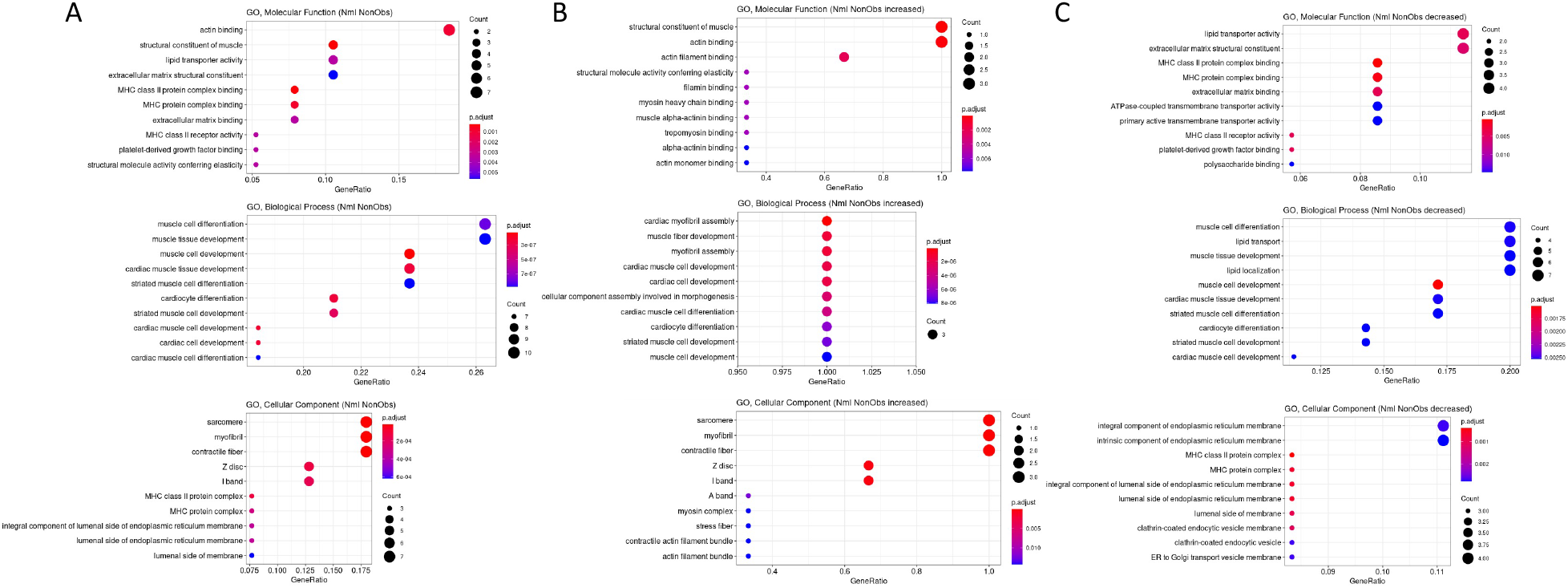
Gene Ontology Enrichment Analysis of Differentially Expressed Genes in nonobstructive HCM. **A**. GO enrichment analysis of all differentially expressed genes in HCM from Table 3. **B**. GO enrichment analysis of Table 3 genes increased in nonobstructive HCM. **C**. GO enrichment analysis of Table 3 genes decreased in nonobstructive HCM.

**Table 3.**
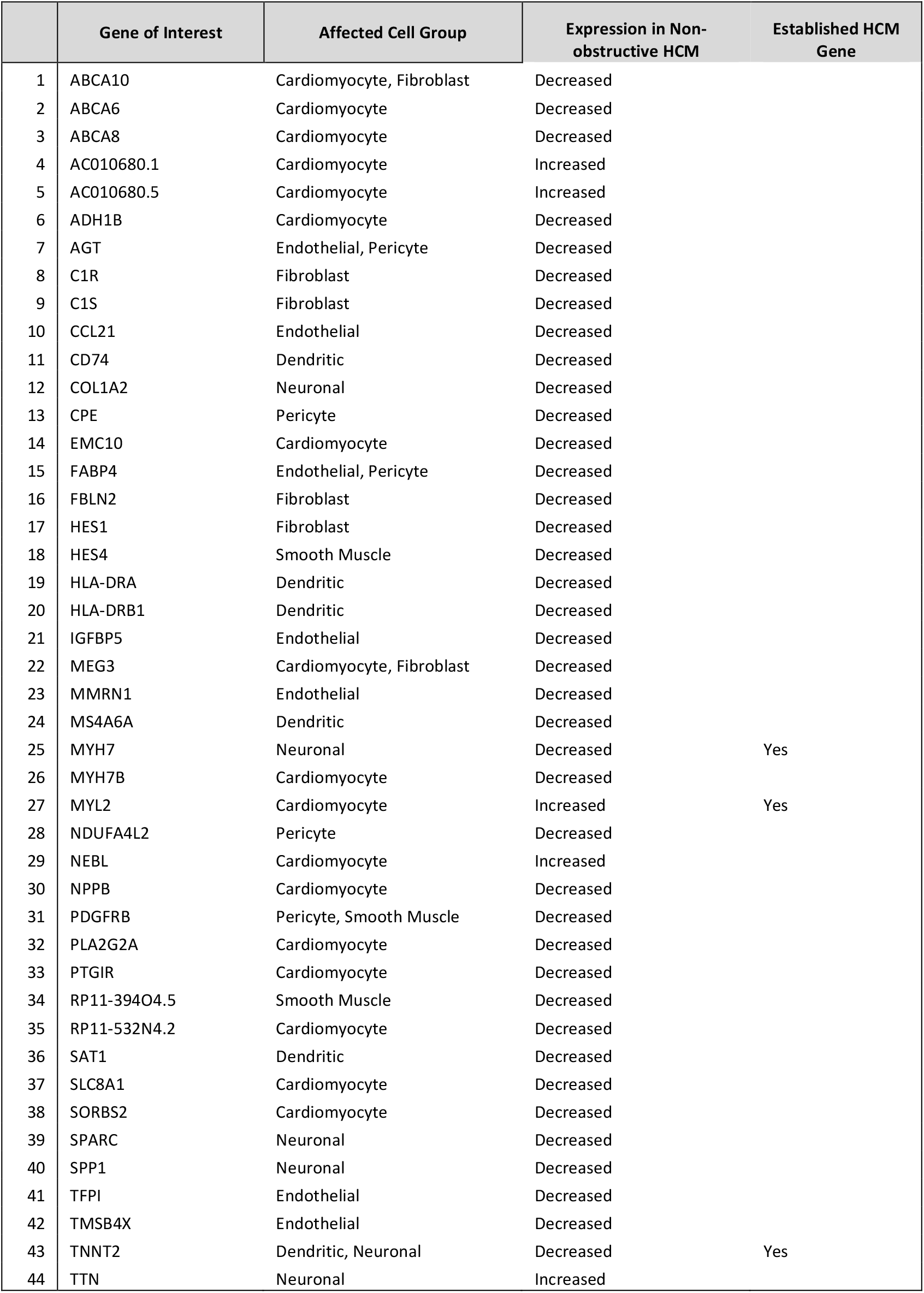
Differentially Expressed Genes in Nonobstructive HCM with Pronounced Changes in Spatial Expression

### Ligand-receptor pair gene expression indicates alteration of intercellular communication in HCM

#### HCM is associated with a general decrease in cell-cell communication but an increase in fibroblast, smooth muscle cell and pericyte to dendritic communication and an increase in smooth muscle cell to leukocyte communication

To quantify potential cardiac cell-cell communication in Normal and nonobstructive HCM IVS tissue, we quantified the number of possible expressed ligand-receptor pairs among cell types as previously described ^23, 24^. We examined the expression of a curated list of 3627 unique human ligand-receptor (L-R) pairs derived by combining a collection of 2557 human L-R pairs ^25^ with another set of 3398 human L-R pairs ^26^ and eliminating duplicate pairs. Ligands and receptors were considered expressed if their associated gene was detectable in ≥ 20% of cells in a cell type. Notably, quantification of cell-cell communication in this study represents potential communication as we account for only expressed ligand-receptor pairs and not the position or boundaries of cell types. Results showed that nonobstructive HCM IVS tissue demonstrated a reduced intercellular communication network among our 8 identified cell types compared to normal IVS tissue (Fig. 4). Quantitatively, the total number of expressed ligand-receptor pairs in the nonobstructive HCM tissue (n=405) was much lower than in Normal tissue (n=710, Fig. 4A, B). Analysis of the broadcast ligands by individual cell types demonstrates that 1) all cells except for smooth muscle cells show a broad decrease in both paracrine and autocrine ligand broadcasting in nonobstructive HCM (Fig. 4B, C, D) and 2) fibroblasts broadcast the greatest number of ligands to the other cell types in both Normal and nonobstructive HCM tissue. Analysis of receptor expression again showed a broad decrease across cell types, except for dendritic cells (Fig. 4B, C, D). Fibroblast communication is particularly high with fibroblast, endothelial, pericyte, smooth muscle, neuronal, and cardiomyocyte cells (Fig. 4). While intercellular communication was generally reduced in nonobstructive HCM tissue compared to Normal tissue, there was several cases of increased nonobstructive HCM communication, from fibroblasts, smooth muscle cells, pericytes and cardiomyocytes to dendritic cells and from smooth muscle cells to leukocytes (Fig 4C, D).

**Fig. 4.**
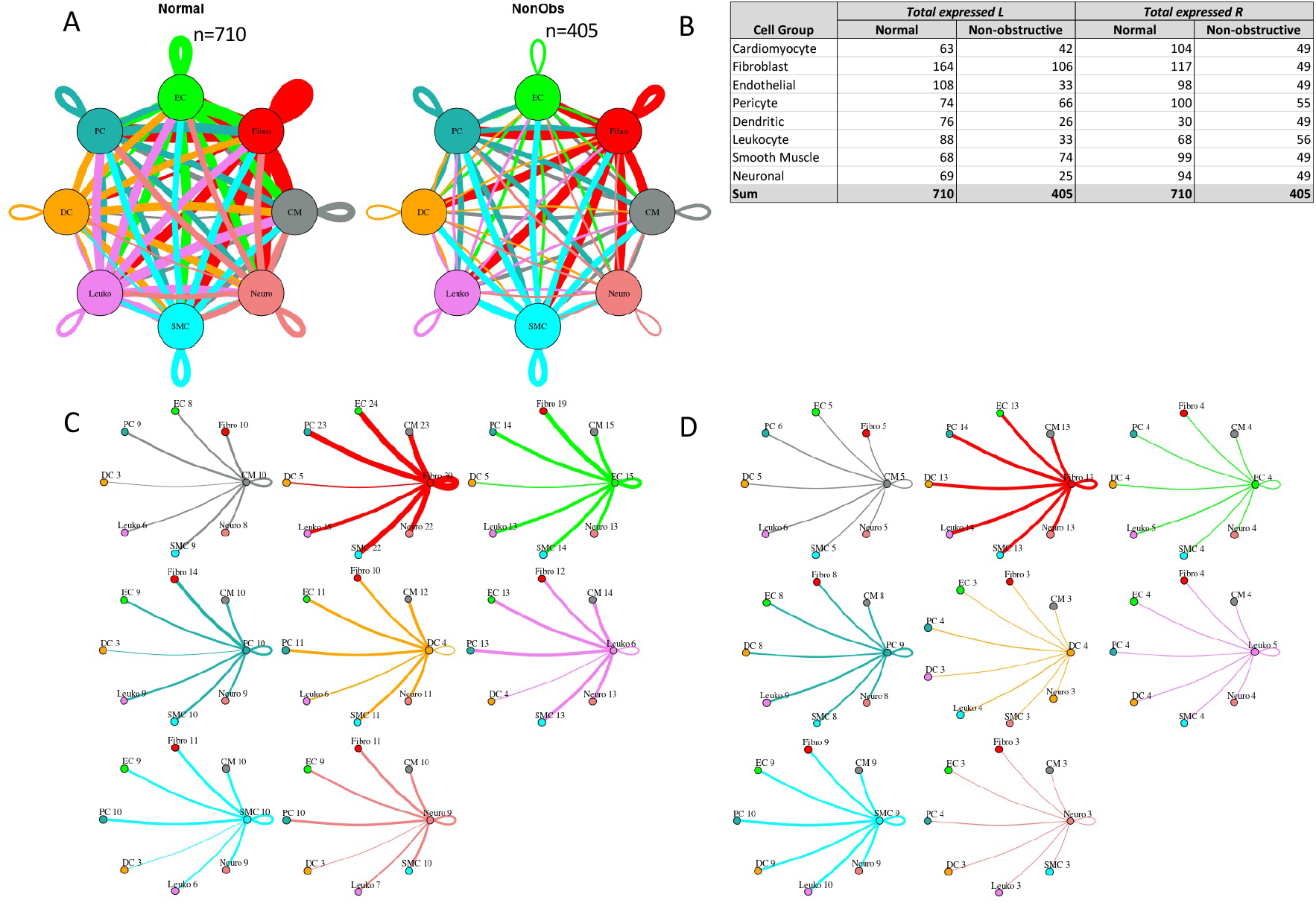
Intercellular communication networks are reduced in nonobstructive HCM. **A**. Cell-cell communication networks between cardiac cell types in normal control (left) and nonobstructive HCM (right) conditions. Line color indicates ligand broadcast by the cell population with the same color. Lines connect to cell types which expressed cognate receptors. Line thickness is proportional to the number of uniquely expressed ligand-receptor pairs. Loops indicates communication within a cell type. **B**. Quantity of ligands and receptors in expressed ligand-receptor pairs described by cell type and condition (Normal or nonobstructive HCM). **C**., **D**. Cell-cell communication networks broken down by cell type in normal control (**C**) and nonobstructive HCM (**D**) conditions. Figure formatting follows panel A. Numbers indicate the quantity of uniquely expressed ligand-receptor pairs between the broadcasting cell type (expressing ligand) and receiving cell type (expressing receptor).

Communication between fibroblasts and dendritic cells increased from 5 to 13 L-R pairs in nonobstructive HCM tissue due primarily to the dendritic cells in nonobstructive HCM tissue gaining *ITGB1* receptor expression to enable communication with several cognate ligands (COL1A2, COL3A1, COL4A1, COL6A1, COL6A2, COL6A3, FN1, LAMA2, LGALS1, LUM; Supplemental Table 5). The increase in smooth muscle cell to dendritic cell communication (3 to 9 L-R pairs) is also primarily due to dendritic cells in nonobstructive HCM tissue gaining *ITGB1* receptor expression, to enable communication with several smooth muscle cell cognate ligands (COL1A2, COL4A1, COL6A1, COL6A2, FN1, LGALS1; Supplemental Table 5). Similarly, increased pericyte to dendritic cell interaction (3 to 8 L-R pairs) is also due primarily to nonobstructive tissue dendritic cells gaining ITGB1 receptor expression, to enable communication with several pericyte cognate ligands (COL1A2, COL4A1, COL6A1, COL6A2, FN1, LGALS1; Supplemental Table 5). The increase in smooth muscle cell to leukocyte communication in nonobstructive HCM (6 to 10 L-R pairs) is due to leukocytes gaining ITGB1 receptor expression, to enable communication with several smooth muscle cell cognate ligands (COL1A2, COL4A1, COL6A1, COL6A2, FN1) (Supplemental Table 6).

#### Changes in ligand-receptor pair gene expression imply alterations in molecular function

To assess the molecular functions potentially affected by changes in ligand-receptor signaling among our 8 identified cell types, GO enrichment analysis was performed on the ligands and receptors in expressed ligand-receptor pairs from both Normal and nonobstructive HCM tissue (Fig. 5). We observed a decrease in most identified ligand molecular functions (34 of 47) in the nonobstructive HCM heart tissue compared to the Normal heart tissue, with complete loss of functions involving insulin receptor binding, insulin-like growth factor receptor binding, lipoprotein particle receptor binding, and structural molecule activity conferring elasticity (note, these are low count normal MFs; Figure 5A). Large decreases in nonobstructive molecular functions relative to Normal (>100 count difference) were found for extracellular matrix structural constituent, extracellular matrix structural constituent conferring tensile strength (56%), growth factor binding (57%), peptidase regulator activity (75%), platelet-derived growth factor binding (56%), protease binding (74%; Figure 5A). Large increases in molecular functions in nonobstructive HCM tissue relative to normal tissue (>50 count difference) were noted for adenylate cyclase binding, calcium channel inhibitor activity, calcium channel regulator activity, channel inhibitor activity, protein kinase activator activity, protein N-terminus binding, protein phosphatase activator activity, protein serine threonine kinase activator activity and titin binding. When ligand molecular function is assessed in individual cell types, a general decrease in most nonobstructive molecular functions is observed compared to the normal condition (Figure 5B-I) with generally more drastic decreases in cardiomyocytes, endothelial cells, dendritic cells, leukocytes, and neuronal cell ligand molecular functions (Figure 5). Less drastic decreases and increases are observed in non-obstructive ligand molecular functions among fibroblasts, pericytes, and smooth muscle cells (Figure 5).

**Fig. 5.**
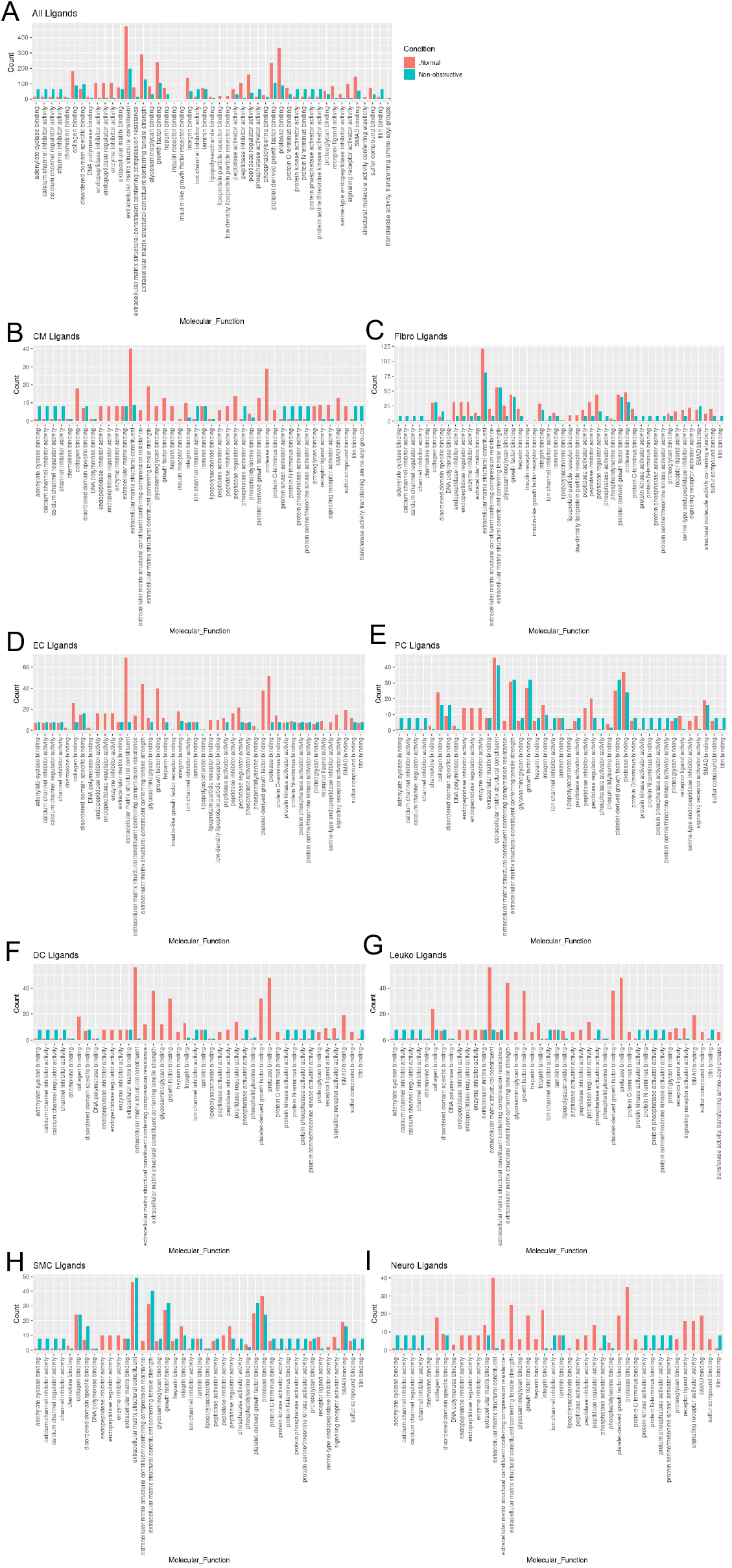
Bar plot representing the total count of ligands (in expressed ligand-receptor pairs) associated with different cellular processes in Normal and nonobstructive HCM IVS Cells. Bar color distinguishes ligand count in normal or nonobstructive HCM conditions. **A**. Comparison of molecular functions across all cell types. **B**. Comparison in cardiomyocytes. **C**. Fibroblasts. **D**. Endothelial Cells. **E**. Pericytes. **F**. Dendritic Cells. **G**. Leukocytes. **H**. Smooth Muscle Cells. **I**. Neurons.

Molecular functions associated with changes in receptor expression align with changes in ligand expression (Fig. 6A) including the finding of a general decrease in most nonobstructive molecular functions compared to the Normal condition (46 of 65 molecular functions; Figure 6). The largest decreases (>100 count difference) in nonobstructive molecular functions compared to Normal included amyloid beta binding, coreceptor activity, lipoprotein, peptide binding, and scavenger receptor activity (Figure 6A). The largest increases (> 100 count difference) in nonobstructive molecular functions relative to Normal included amide binding, calcium molecular functions, calmodulin binding, channel/gated channel molecular functions, divalent inorganic cation transmembrane transporter activity, metal ion transmembrane transporter activity, protein kinase A activity, and sulfur compound binding (Figure 6A). Trends in receptor molecular functions in individual cell types are notable for increases related to signal transduction in dendritic cells and leukocytes, particularly calcium channel activity and protein kinase A activity in dendritic cells.

**Fig. 6.**
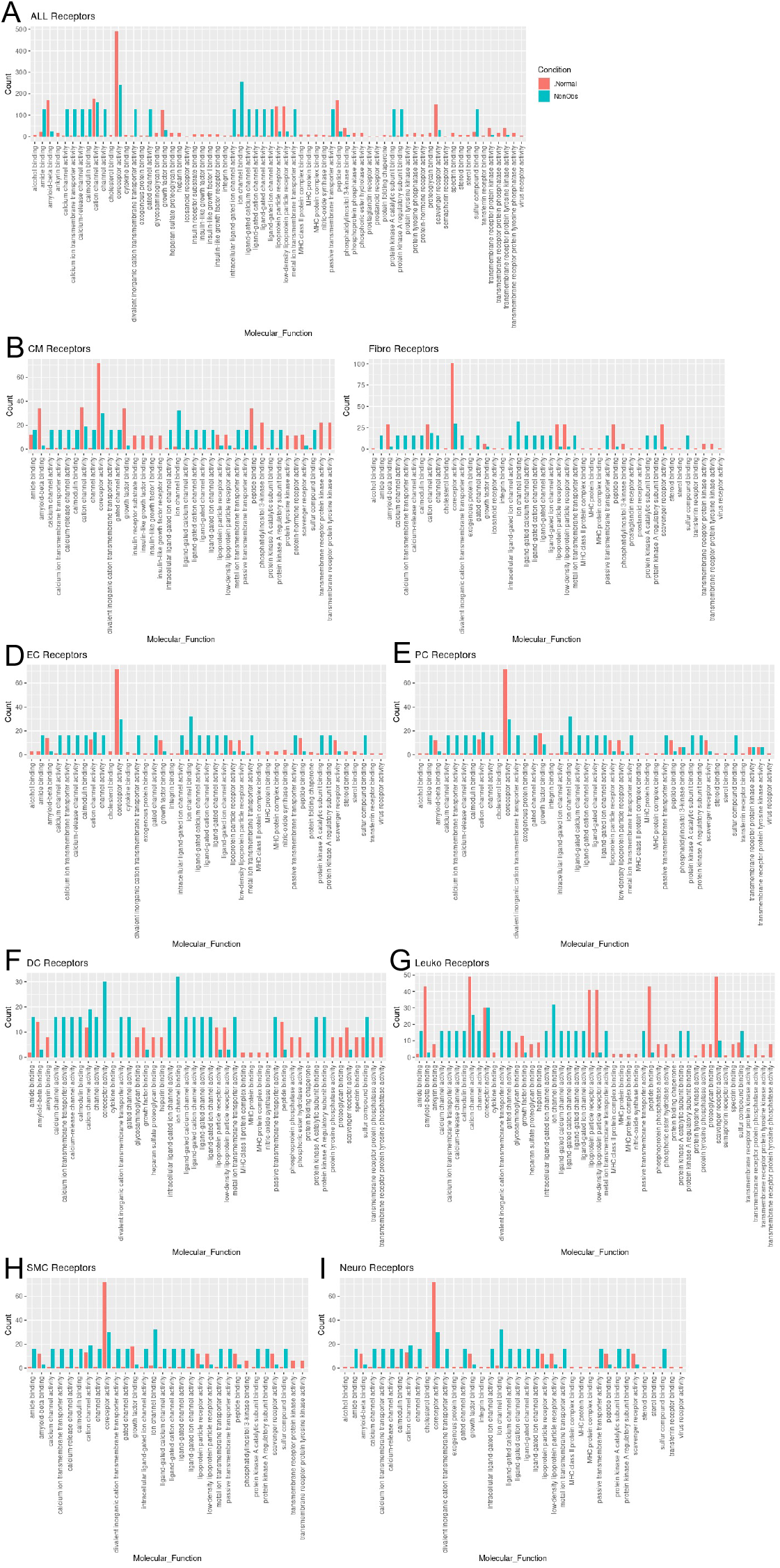
Bar plot representing the total count of receptors (in expressed ligand-receptor pairs) associated with different cellular processes in Normal and nonobstructive HCM IVS Cells. Bar color distinguishes receptor count in normal or nonobstructive HCM conditions. **A**. Comparison of molecular functions across all cell types. **B**. Comparison in cardiomyocytes. **C**. Fibroblasts. **D**. Endothelial Cells. **E**. Pericytes. **F**. Dendritic Cells. **G**. Leukocytes. **H**. Smooth Muscle Cells. **I**. Neurons.

#### Fibroblast subtypes show subtype-specific changes in cell-cell communication

To better understand the interaction between the different fibroblast populations and other cell types in the IVS, we examined the potential ligand-receptor communication through cell network plots for all fibroblast subtypes alongside the other previously identified 7 cell types (Fig. 7A, C, D). In Normal tissue, all fibroblast subtypes broadcast ligands extensively to generate a dense intercellular network. In nonobstructive HCM tissue, the intercellular communication network for fibroblast subtypes shows a reduced number of broadcasting ligands compared to Normal tissue, (1502 versus 2636 respectively; Fig. 7A, B). Fibroblast cluster 2 generated the highest number of broadcasting ligands in both conditions. The largest decreases in potential interactions occurred between fibroblast cluster 2 and fibroblast clusters 2 through 6 (Supplemental Table 7) and between fibroblast cluster 5 and fibroblast clusters 2 through 6 (Supplemental Table 8). In fibroblast cluster 2 from nonobstructive HCM tissue, there was a disproportionate reduction in expression of cognate ligands for the ITGB1 receptor (COL18A1, COL5A1, COL5A2, FBLN1, FBN1, HSPG2, LAMC1, LGALS3BP, NID1, THBS2) and also of ligands for the LRP1 receptor (APP, C3, CALR, CTGF, HSPG2, LRPAP1, SERPINE2, SPERPING1, TFPI). *ITGB1* and *LRP1* were expressed in both normal and nonobstructive HCM fibroblasts from cluster 2. Interaction between fibroblast cluster 2 and fibroblast clusters 3-5 was reduced in nonobstructive HCM primarily due to loss of the LRP1 receptor and several cognate ligands for the ITGB1 receptor (COL18A1, COL5A1, COL5A2, FBLN1, FBN1, HSPG2, LAMC1, LGALS3BP, NID1, THBS2). *ITGB1* was expressed in fibroblast clusters 3-5 in both normal and nonobstructive HCM tissue.

**Fig. 7.**
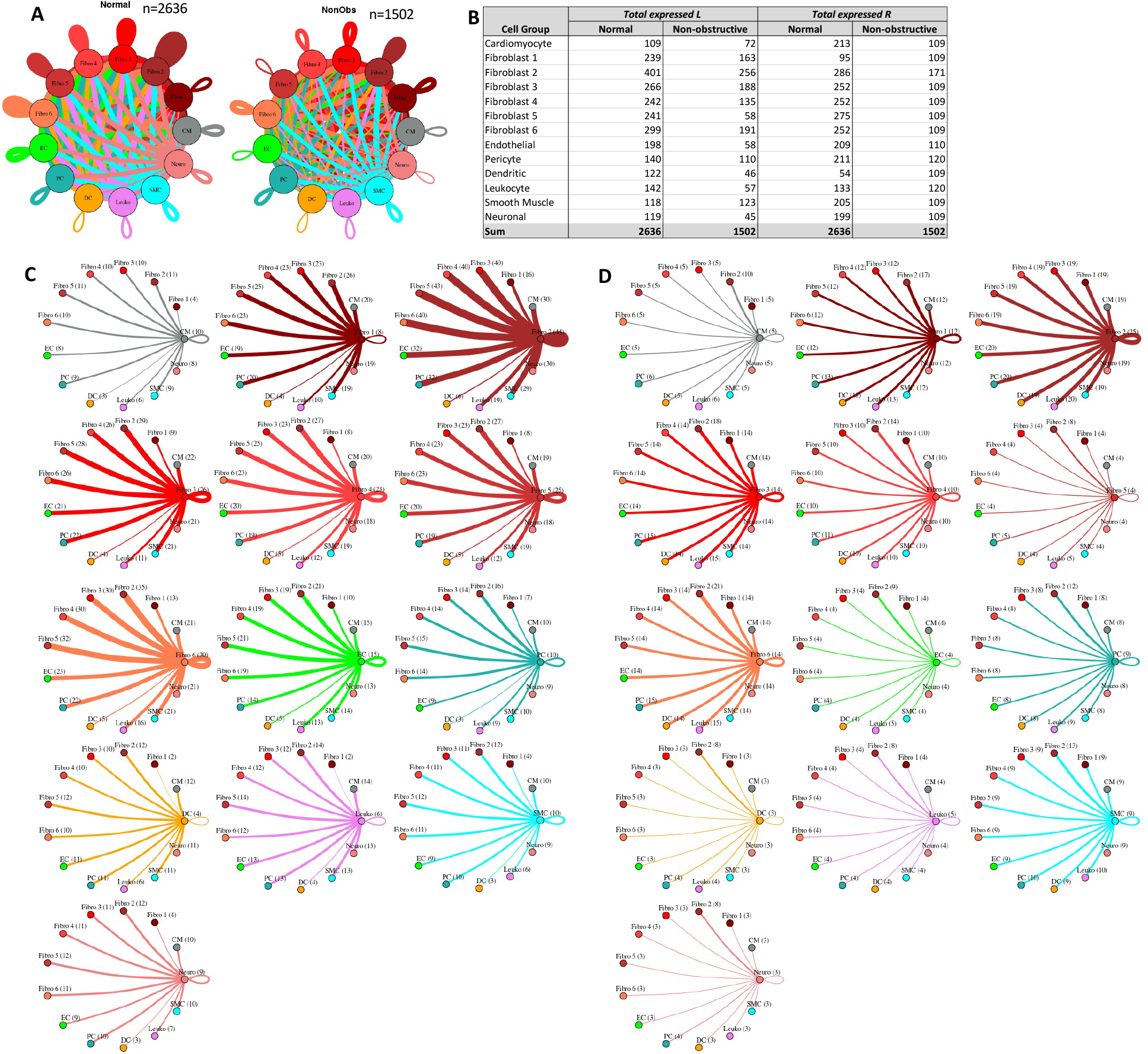
Cell-cell communication networks between fibroblast subtypes and other heart cells in normal control and nonobstructive HCM conditions. **A**. Comparison of Normal (left) and nonobstructive HCM (right) communication networks. Line color indicates ligand broadcast by the cell population with the same color. Lines connect to cell types which expressed cognate receptors. Line thickness is proportional to the number of uniquely expressed ligand-receptor pairs. Loops indicates communication within a cell type. **B**. Quantity of ligands and receptors in expressed ligand-receptor pairs described by cell type and condition (Normal or nonobstructive HCM). **C, D**. Cell-cell communication networks broken down by cell type and fibroblast cluster in normal control (**C**) and nonobstructive HCM (**D**) conditions. Figure formatting follows panel A. Numbers indicate the quantity of uniquely expressed ligand-receptor pairs between the broadcasting cell type (expressing ligand) and receiving cell type (expressing receptor).

#### Cardiomyocyte subtypes show subtype-specific changes in cell-cell communication

To better understand the interaction between the different cardiomyocyte populations and other cell types in the IVS, we performed L-R analysis for all ten cardiomyocyte subtypes and the other 7 cell types (Fig. 8). In nonobstructive HCM tissue, there is again a general decrease in intercellular communication (2634 L-R pairs in normal, reduced to 2026 in nonobstructive HCM; Fig. 8A, B), but the degree of reduction is lower than seen when subtypes are not considered or when fibroblast subtypes are considered. The largest decreases occur in ligands broadcast from endothelial cells and in receptors expressed by fibroblasts and the cardiomyocyte 4 cluster. Notably, there are increases in ligands broadcast by cardiomyocyte clusters 1, 2, 5, 9 and by smooth muscle cells and in receptors in cardiomyocyte clusters 1, 3 and 9 and dendritic cells in nonobstructive HCM (Fig. 8B). The greatest reductions in intercellular L-R connections involve fibroblasts broadcasting to fibroblasts and cardiomyocyte cluster 4 (Supplemental Table 9), and from endothelial cells broadcasting to fibroblasts, cardiomyocyte cluster 4 and cardiomyocyte cluster 8 (Supplemental Table 10). In both cases, reduction in intercellular communication is due to loss of LRP1 receptor expression in fibroblasts as well as loss of ligands for ITGB1 (COL11A1, FBLN1, FBN1, HSPG2, LAMB1, LAMC1, NID1, VCAN) in fibroblasts and endothelial cells. Notably, *ITGB1* is expressed in fibroblasts and cardiomyocyte clusters 4 and 8 in both Normal tissue and nonobstructive HCM tissue. Increases in communication from cardiomyocyte clusters 2, 5, fibroblasts and smooth muscle cells to cardiomyocyte cluster 9 is due to gain of expression of *ITGB1* in cardiomyocyte cluster 9 (Supplemental Table 11). Increased communication from cardiomyocyte cluster 5 to dendritic cells is also due to gain of expression of *ITGB1* in dendritic cells (Supplemental Table 12).

**Fig. 8.**
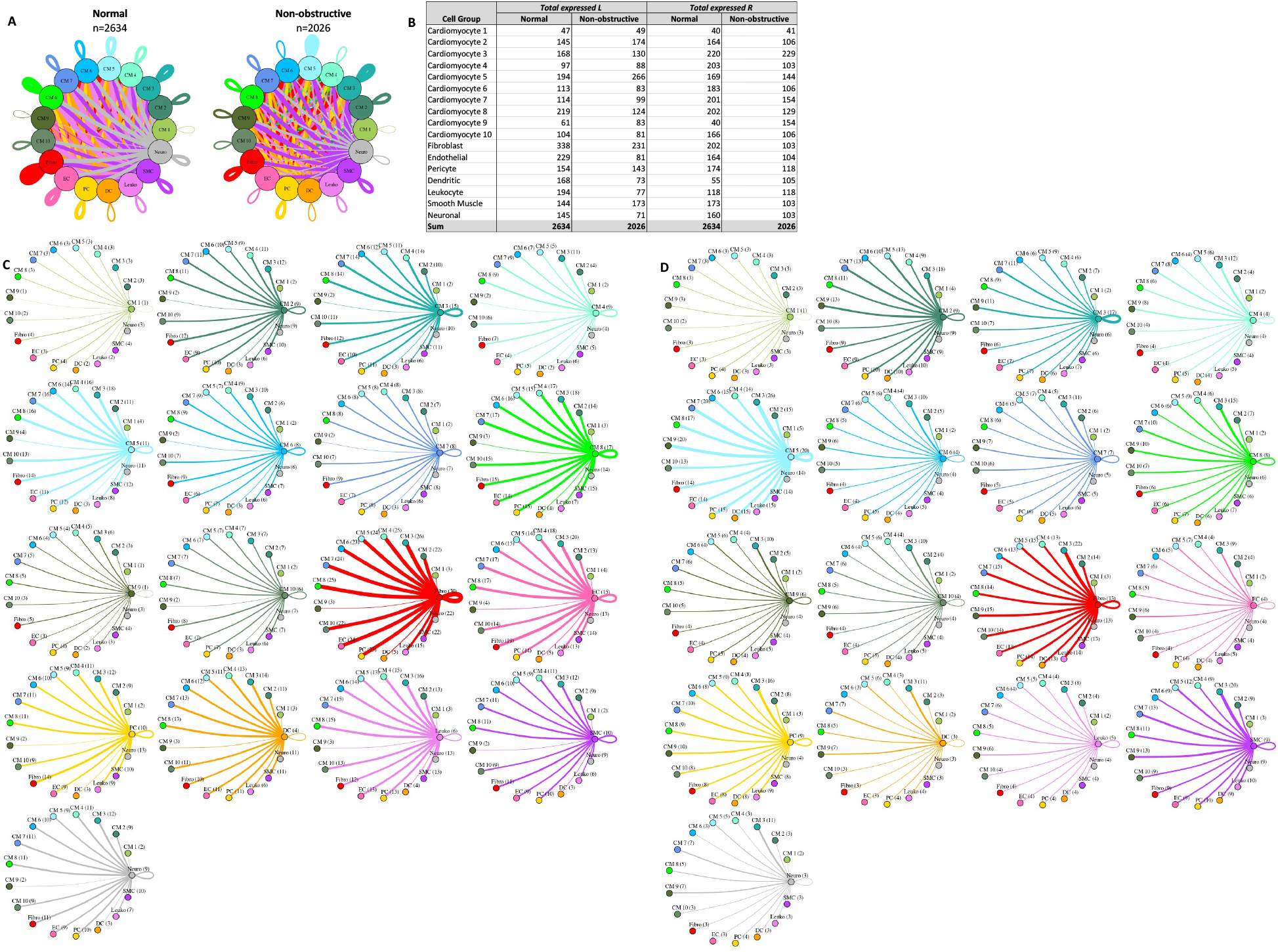
Cell-cell communication networks between cardiomyocyte subtypes and other heart cells in normal control and nonobstructive HCM conditions. **A**. Comparison of Normal (left) and nonobstructive HCM (right) communication networks. Line color indicates ligand broadcast by the cell population with the same color. Lines connect to cell types which expressed cognate receptors. Line thickness is proportional to the number of uniquely expressed ligand-receptor pairs. Loops indicates communication within a cell type. **B**. Quantity of ligands and receptors in expressed ligand-receptor pairs described by cell type and condition (Normal or nonobstructive HCM). **C, D**. Cell-cell communication networks broken down by cell type and cardiomyocyte cluster in normal control (**C**) and nonobstructive HCM (**D**) conditions. Figure formatting follows panel A. Numbers indicate the quantity of uniquely expressed ligand-receptor pairs between the broadcasting cell type (expressing ligand) and receiving cell type (expressing receptor).

#### Cardiomyocyte subtypes and fibroblast subtypes together show cell subtype-specific alterations in cell-cell communication

Given that cardiomyocytes and fibroblasts are the most numerous cells in the heart and demonstrate the most subtypes in our study, we examined the intercellular networks between cardiomyocyte subtypes and fibroblast subtypes in normal tissue and nonobstructive HCM. The total number of expressed ligand-receptor pairs in the Normal tissue was greater than in the nonobstructive HCM tissue (3354 and 2399 expressed ligand-receptor pairs respectively, Fig. 9). The largest decreases in cellular communication involved fibroblast cluster 5, through reduction in both broadcasted ligands and expressed receptors. The largest reductions in L-R pairs occurred between fibroblast clusters 2 or 5 and fibroblast clusters 2-6 (Supplemental Table 13) and resulted from loss of ligands for ITGB1, which was still expressed in the recipient cells in both normal and nonobstructive HCM, or from loss of the LRP1 receptor in recipient fibroblast clusters except for fibroblast cluster 2. The largest increases in communication due to ligand broadcasting involved cardiomyocyte clusters 2, 5 and 9 and increases due to receptor expression involved cardiomyocyte clusters 1, 3, 9 and fibroblast cluster 1 (Fig. 9B). Increased communication to cardiomyocyte cluster 9 from fibroblast clusters 1-5 and cardiomyocyte clusters 2, 3 and 5 were due to gain of ITGB1 expression in cardiomyocyte cluster 9 (Supplemental Table 14).

**Fig. 9.**
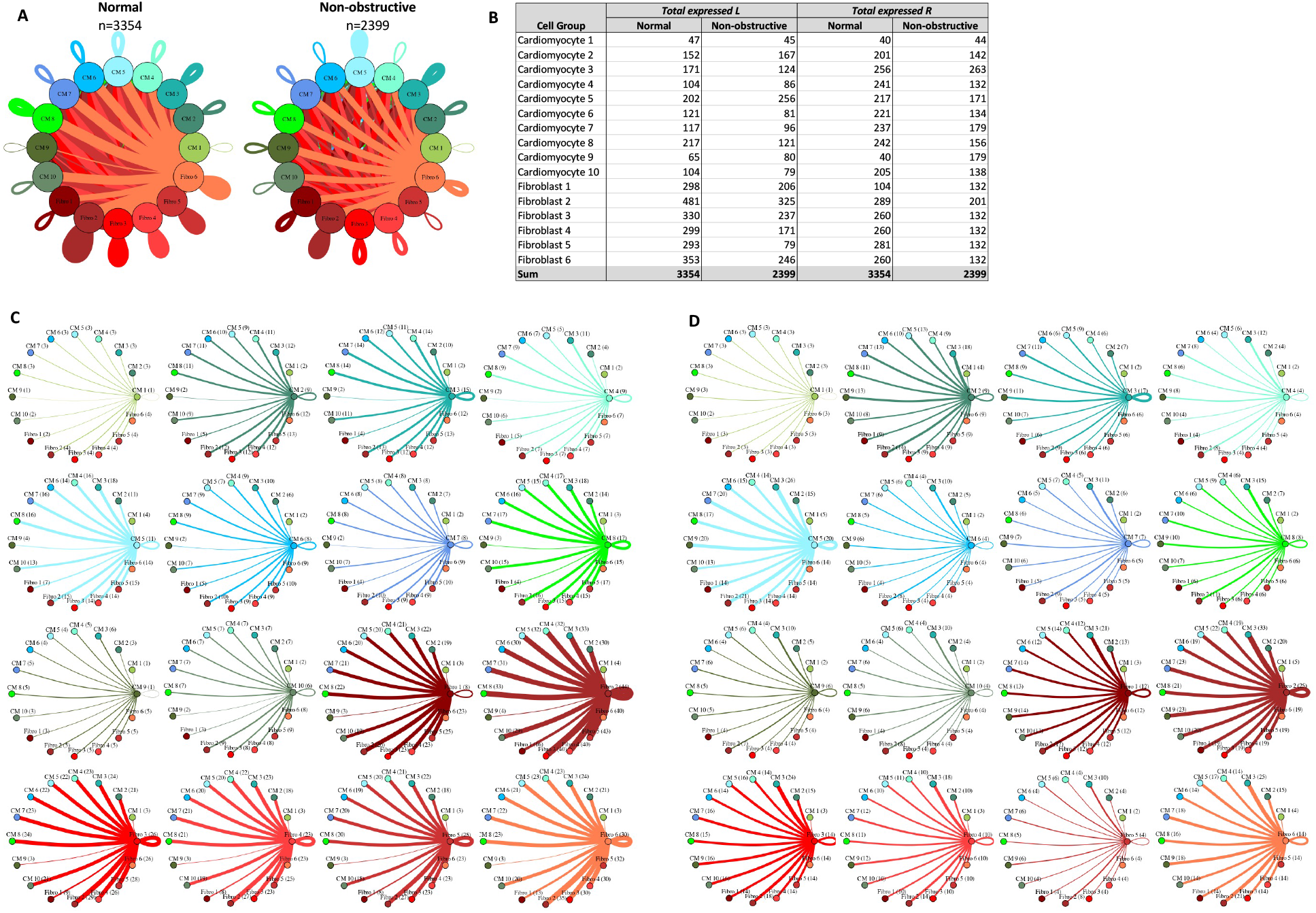
Cell-cell communication networks between cardiac fibroblast and cardiomyocyte subtypes in normal control and HCM conditions. **A**. Overall communication network between cardiomyocytes and fibroblasts. Line color indicates ligand broadcast by the cell population with the same color. Lines connect to cell types which expressed cognate receptors. Line thickness is proportional to the number of uniquely expressed ligand-receptor pairs. Loops indicates communication within a cell type. **B**. Quantity of ligands and receptors in expressed ligand-receptor pairs described by cell type and condition (Normal or nonobstructive HCM). **C, D**. Cell-cell communication networks broken down by cardiomyocyte cluster and fibroblast cluster in normal control (**C**) and nonobstructive HCM (**D**) conditions. Figure formatting follows panel A. Numbers indicate the quantity of uniquely expressed ligand-receptor pairs between the broadcasting cell type (expressing ligand) and receiving cell type (expressing receptor).

## Discussion

HCM has long been considered a disease of the sarcomere based on the association of sarcomere gene mutations with HCM, but the contributions from other loci are increasingly appreciated ^2, 3^, as are potential mechanisms aside from sarcomere dysfunction ^27^. We have previously examined single nuclei gene expression patterns in obstructive HCM. In this prior study, we did not identify any HCM-specific cell populations but observed a profound reduction in intercellular communication, especially in pathways mediated by ITGB1, and altered extracellular matrix gene expression, which may explain some of the nonmyocyte phenotypes seen in HCM such as fibrosis and link these phenotypes to alterations in mechanical signal transduction ^16^. In the current study, we performed a single nucleus transcriptome analysis of tissue from patients with nonobstructive HCM, using tissue from the IVS harvested at the time of cardiac transplantation. This patient population is distinct from those with obstructive HCM in that they do not have LVOT obstruction and generally have more severe, intractable heart failure that results in cardiac transplantation. Determining gene expression patterns at the single cell level in nonobstructive HCM thus has the potential for facilitating precision medicine approaches for this specific population, through overlapping and unique pathogenic mechanisms relative to obstructive HCM.

The single nuclei gene expression patterns for nonobstructive HCM parallel those for obstructive HCM in that there are no disease-specific cell clusters and there is a reduction in intercellular communication, particularly in pathways involving ITGB1 and its associated ECM ligands. As with obstructive HCM, changes in gene expression involve sarcomere genes known to be associated with HCM and GO analysis indicates that gene expression changes are relevant to the extracellular matrix and sarcomere function, indicating that these are common, fundamental disease processes in HCM, regardless of LVOT obstruction. We have previously noted that a reduction in ECM associated gene expression and molecular function in obstructive HCM is counterintuitive based on the association between HCM and increased fibrosis and have suggested that the reduction in gene expression may represent a negative feedback loop that follows the accumulation of ECM protein in tissue ^16^. Concordant findings in nonobstructive HCM can be explained similarly. Precision medicine approaches that target integrin signaling and ECM interaction may one day prove useful for both obstructive and nonobstructive HCM.

A notable difference between obstructive and nonobstructive HCM in our current study, however, involves the unexpected increases in communication between fibroblasts, smooth muscle cells and pericytes with dendritic cells and between smooth muscle cells and leukocytes in the nonobstructive HCM condition relative to the control condition. Immune cells have been demonstrated to play important roles in development of cardiac hypertrophy and fibrosis in a pressure overload model of rodent hypertrophy (reviewed in ^28^). Dendritic cells function as antigen presenting cells that activate T cell responses and depletion of these cells reduces cardiomyocyte hypertrophy, cardiac fibrosis, LV remodeling and inflammatory cell infiltration after pressure overload ^29^. A role for dendritic cells in human heart failure is less well-established, although a trend toward elevated numbers in blood has been reported ^30^. Integrin-mediated signaling has been shown to be an important determinant of dendritic cell function, with ITGB1 determining dendritic morphology ^31^ and promoting an anti-inflammatory phenotype in bone-marrow-derived dendritic cells ^32^. Our data linking dendritic cells to fibroblasts, smooth muscle cells and pericytes by ITGB1 signaling is novel and suggests an important role for these cells in the pathological hypertrophy, cardiac fibrosis and vascular abnormalities seen in nonobstructive HCM. Our additional novel observation that signal transduction related molecular functions such as calcium channel activity and protein kinase A activity are increased in dendritic cells from nonobstructive HCM hearts also provides additional potential pathways for precision-medicine guided therapy for nonobstructive HCM.

We have previously noted extensive cardiomyocyte and fibroblast diversity in the human IVS, likely due to the diverse origins of the cells within this anatomic region ^15, 16^. We have also noted that the different cardiomyocyte and fibroblast subtypes exhibit specific intercellular communication profiles ^16, 24^ and the phenomenon is observed again in this study. Specific cardiomyocyte and fibroblast subtypes appear to adopt distinct functional roles relevant to the disease process by shifting patterns of gene expression to increase or decrease interactions of cell types. An ongoing challenge is to determine which of these shifts is most relevant to the disease process and thus identify potential therapeutic targets. Future spatial transcriptomic experiments assigning single cell populations to anatomic pathological features of HCM, such as areas of myocyte disarray, fibrosis, focal myocyte hypertrophy, and vascular abnormalities will likely be informative. Our study is the first to characterize transcription patterns in nonobstructive HCM at single cell resolution and provides an important resource for future investigation of the complex cellular interplay in nonobstructive HCM.

## Methods

### Study Patients, Sample Collection and Processing

A total of 6 patients with end stage, nonobstructive HCM scheduled for cardiac transplantation were approached for written informed consent to allow their tissue to be used for research. Those who consented underwent surgery and tissue was collected. Explanted heart tissue sample processing was done as previously described ^15, 16^. 100 mg of collected interventricular septum tissue was minced into 1 mm^3^ pieces, placed in 0.5 mL of CryoStor CS10 Freeze Media (STEMCELL Technologies), and stored in a MrFrosty (ThermoFisher) at 4°C for 10 minutes and then transferred to −80°C overnight. Bulk RNA was isolated from a piece of tissue using the Qiagen RNeasy Plus Micro kit and then assessed on the Agilent Bioanalyzer 2100. Samples with an RNA Integrity Number greater than 8.5 were used in library preparation. Sample collection was approved by the Tufts University/Medical Center Health Sciences Institutional Review Board under IRB protocol # 9487. Patient characteristics were obtained from the medical record and are shown in Table 1. Tissue from organ donor patients without underlying cardiac disease was obtained and processed as described previously ^15^.

### Nuclei isolation, library preparation, and sequencing

Generation of single nuclei sequencing libraries was performed as previously described ^15, 16^. Cryopreserved samples were thawed at 37°C and placed on ice. Nuclei were isolated via Dounce homogenization as previously described ^33^. Homogenates were filtered through a Pluristrainer 10 µM cell strainer (Fisher Scientific) into a pre-chilled tube. Nuclei were pelleted by centrifuging at 500 x g for 5 min at 4°C. Nuclei pellets were washed and pelleted according to manufacturer protocol (10x Genomics). Nuclei were stained with trypan blue and counted on a hemocytometer to determine concentration prior to loading of the 10x Chromium device and samples were diluted to capture ~10,000 nuclei. Nuclei were separated into Gel Bead Emulsion droplets using the 10x Chromium device according to the manufacturer protocol (10x Genomics). Sequencing libraries were prepared using the Chromium Single Cell 3’ reagent V2 kit according to manufacturer’s protocol. Libraries were multiplexed and sequenced on a NovaSeq S4 (Illumina) to produce ~50,000 reads per nucleus. Single nuclei RNAseq data for normal IVS tissue from 4 donors is available in the Gene Expression Omnibus database under accession number GSE161921 ^15^. Data for nonobstructive HCM IVS tissue and additional normal heart donors is available under accession number GSE181764.

### Clustering of Cells by Gene Expression Pattern and Assignment of Cell Type Identity

Clustering of cells and assignment of cell identity was done as previously described ^15, 16^. Sequencing reads were processed using Cell Ranger version 6.0.1 ^17^. The gene expression matrix was subset to only include reads from the nuclear genome. Quality control (QC) filtering, clustering, dimensionality reduction, visualization, and differential gene expression were performed using the R package Seurat version 3.0. Each dataset was filtered so that genes that were expressed in three nuclei or more were included in the final dataset. The dataset was further sublet to exclude nuclei that had fewer than 200 genes expressed to remove droplets containing only ambient RNA, and to exclude nuclei with greater than 2000 genes to remove droplets that contained two nuclei. Datasets were individually normalized and integrated using Seurat’s SCTransform development workflow to reduce batch effects ^18^. Optimal clustering resolution was determined using Clustree version 0.4.3 ^34^ to identify the resolution where the number of clusters stays stable and was determined to be 0.9 for the integrated dataset. Assignment of cell identity to each cluster was performed using four separate analyses. Expression of known cell-specific gene markers were used to identify major cell types, as done previously ^13, 15, 24, 35^. The top 20-30 differentially expressed genes in each cluster were also compared with cell type gene expression markers from the PanglaoDB database https://panglaodb.se ^19^ to independently assign cell type. Entire sets of differentially expressed genes for each cluster were also subjected to Ingenuity Pathway Analysis ^21^ and their inferred functions were used to identify cluster cell types independently. Upregulated genes from each cluster were also subject to Gene Ontology biological process association using GoStats ^20^ and these associations were used to refine cell type assignment further.

### Trajectory Analysis and Identification of Differentially Expressed Genes

Trajectory analysis was performed using Monocle3 ^22^ to determine the relationship between subtypes of cells identified in our clustering analysis as previously described ^16^. Since our data do not represent a developmental time course, we determined the root nodes for each subtype by hierarchical clustering prior to generating trajectories and assigning pseudotime to each cell. Each cell type was analyzed in three cohorts: 1. Normal and nonobstructive HCM cells together; 2. Normal cells alone; 3. Nonobstructive HCM cells alone. Once trajectories were established, differential expression between Normal and HCM cells by cell type and cluster was performed by fitting a generalized linear regression model to each gene according to the formula log(yi) = β0 + βtxt. The coefficients β0 and βt were extracted from each model and tested for significant difference from zero using the Wald test, which assesses constraints on statistical parameters based on the weighted distance between the unrestricted estimate and its hypothesized value under the null hypothesis, where the weight is the precision of the estimate. A gene was determined to be differentially expressed between Normal and HCM conditions in a cell type or cluster if the Wald test produced an adjusted p-value ≤0.05.

Differentially expressed genes over trajectory paths in UMAP space (i.e., spatial autocorrelation) was performed in Monocle3 using Moran’s I statistic. Moran’s I statistic is a value that varies from −1 to 1, where −1 indicates perfect dispersion, 0 indicates no spatial autocorrelation, and 1 indicates perfect positive autocorrelation (i.e., nearby cells in have similar gene expression values in focal region of UMAP space). For each Normal and nonobstructive HCM cell type, a gene was determined to be differentially expressed over space if the associated Moran’s I statistic value was positive, paired with a significant adjusted p-value ≤ 0.05, and expressed in ≥ 1% of associated cells. Since many genes showed differential expression over space, further conservative filtering was performed in which genes with Moran’s I statistic available in a single class (i.e., Normal or HCM) were filtered by Moran’s I statistic values >0.1. For genes with Moran’s I statistics available in both classes (i.e., Normal and HCM), genes were filtered by an absolute difference >0.1. GO analysis of molecular function and biological process associated with differentially expressed genes was done using the online tools at uniprot.org/uniprotkb ^36^.

### Analysis of Ligand-Receptor Pair Gene Expression to Discover Intercellular Communication Pathways

To quantify potential cardiac cell-cell communication in Normal and HCM hearts, cell communication networks were plotted in igraph version 1.2.6 ^37^ and compared on the basis of ligand-receptor pair gene expression. Our cell-cell communication networks were derived as described previously ^23^, using a list of 2557 human ligand-receptor pairs ^25^ combined with another list of 3398 human ligand-receptor pairs ^26^, to give a total of 3627 unique human ligand-receptor pairs, largely as described previously ^16^. A ligand or receptor for each cell type or cluster was considered expressed if the corresponding gene showed an above zero gene count in ≥ 20% of cells in our snRNAseq data. We initially analyzed the potential signaling interactions between the 8 cell types identified in our snRNAseq data. Lines in our cell networks connect two cell types and represent expressed human ligand-receptor pairs (i.e., potential cell-cell communication between a broadcasting (ligand) and recipient (receptor) cell types. Line color in our networks represents the broadcasting ligand source. Line thickness is proportional to the number of uniquely expressed ligand-receptor pairs. Cell-cell communication networks were also analyzed by fibroblast cluster along with other cell types, by cardiomyocyte cluster and other cell types and by fibroblast clusters and cardiomyocyte clusters. GO analysis of differentially expressed ligand receptor pairs was performed using the R package clusterProfiler ^38^.

### Statistics

We used mixed effects models to analyze cell type specific differential expression, while taking into account the variability between and within subjects. A sample size of 6 cases and 6 controls, with an average of 3,000 cells per subject, will provide 80% power to detect fold change of expression ranging between 1.3 for an intracluster correlation f 0.01, and 2 for an intracluster correlation of 0.1. The power calculations were pretty stable for sample sizes ranging between 500 and 3000 cells per subject. We used a Bonferroni correction for 10,000 tests to fix the level of significance. The power calculations were conducted using Power Analysis and Sample Size software (PASS).

Other statistical methods to cluster cells in UMAP space and control for batch effects using the built-in functionality of Seurat ^18^, to determine cluster stability by Clustree ^34^ and to perform Gene Ontology Analysis using well documented software programs ^38^ are described in above methods sections and in cited references. Statistical methods to compare gene expression along pseudotime trajectories using linear regression or spatial autocorrelation using the built-in functionality of Monocle3 are described above and in the original cited reference ^22^.

### Study Approval

Explanted HCM heart sample collection was approved by the Tufts University/Medical Center Health Sciences Institutional Review Board under IRB protocol # 9487. All subjects gave their informed consent for inclusion before they participated in the study. The study was conducted in accordance with the Declaration of Helsinki. Unused donor hearts were obtained in deidentified fashion from New England Donor Services under a Tufts University/Medical Center Health Sciences IRB approved research protocol after being designated as not human subjects research.

## Data Availability

All data produced are available online in the Gene Expression Omnibus database under accession numbers GSE161921 and GSE181764

https://www.ncbi.nlm.nih.gov/geo/query/acc.cgi?acc=gse181764

https://www.ncbi.nlm.nih.gov/geo/query/acc.cgi

## Author contributions

Conceptualization: MTC

Data Curation: CJC, AL, GP, MTC

Formal Analysis: CJC, MTC

Methodology: CJC, AL, JA, GP, MTC

Investigation: AL, CJC, JA, GP, MTC

Visualization: CJC, AL, MTC

Funding acquisition: MTC, AL

Project administration: MTC

Resources: AL, MTC

Software: AL, CJC

Supervision: MTC

Writing – original draft: MTC

Writing – review & editing: AL, CJC, JA, GP, MTC

## Acknowledgements

We thank Paola Sebastiani for helpful discussions regarding statistics.

## Funding

American Heart Association Innovative Project Award 18IPA34170294 (MTC)

National Center for Advancing Translational Sciences, National Institutes of Health, Award Number UL1TR002544 (MTC)

National Heart, Lung, and Blood Institute of the National Institutes of Health, Award Number F32HL147492 (AL)

Beals Goodfellow Award for CardioVascular Research at Tufts Medical Center (AL)

## Notes

The authors have declared that no conflict of interest exists.

### Competing Interest Statement

The authors have declared no competing interest.

### Funding Statement

This study was funded by
American Heart Association Innovative Project Award 18IPA34170294 (MTC)
National Center for Advancing Translational Sciences, National Institutes of Health, Award Number UL1TR002544 (MTC)
National Heart, Lung, and Blood Institute of the National Institutes of Health, Award Number F32HL147492 (AL)

### Author Declarations

The IRB of Tufts Medical Center gave ethical approval for this work.

### Summary of Updates

Reference 16 has been updated.

