## supplemental tables and figure for "Single nucleus RNA-sequencing reveals altered intercellular communication and dendritic cell activation in nonobstructive hypertrophic cardiomyopathy"

### Supplemental Table 1. Biomarker list with citations

| Cell type | Biomarker | Reference | Note |
|---|---|---|---|
| Cardiomyocyte | MYBPC3 | Selewa et al., 2020; Litviňuková et al., 2020 | |
| | MYH7B | Cui et al., 2019 | |
| | MYH7 | Cui et al., 2019; Wang et al., 2020; Litviňuková et al., 2020 | |
| | TNNT2 | Selewa et al., 2020; Lother et al., 2018 | |
| | LDB3 | Jia et al., 2018 | |
| | FHOD3 | Tucker et al., 2021 | |
| | PLN | Lother et al., 2018; Litviňuková et al., 2020 | Mature |
| | ACTC1 | Wang et al., 2020; Litviňuková et al., 2020 | Mature |
| | TNNC | McLellan et al., 2020; Jia et al., 2018 | Mature |
| | GATA4 | Jia et al., 2018; Hu et al., 2018 | |
| | MYOCD | Jia et al., 2018; Hu et al., 2019 | Developing |
| | MHRT | McLellan et al., 2020; Hu et al., 2019 | Developing |
| | NPPA | Cui et al., 2019; Wang et al., 2020; Litviňuková et al., 2020 | Trabecular |
| | MYH6 | Lother et al., 2018; Cui et al., 2019 | |
| | ATP2A2 | Lother et al., 2018; Wang et al., 2020 | |
| | RYR2 | Lother et al., 2018; Litviňuková et al., 2020; Hu et al., 2019 | |
| | MYL2 | Wang et al., 2020; Litviňuková et al., 2020 | |
| Fibroblasts | FMO2 | Tucker et al., 2021 | |
| | POSTN | Cui et al., 2019; Selewa et al., 2020; McLellan et al., 2020; Litviňuková et al., 2020 | |
| | COL6A3 | Wang et al., 2020 | |
| | COL3A1 | McLellan et al., 2020; Cui et al., 2019 | |
| | FBN1 | McLellan et al., 2020; Hu et al., 2018 | |
| | COL1A1 | Lother et al., 2018; McLellan et al., 2020; Cui et al., 2019 | |
| | DDR2 | Wang et al., 2020 | |
| | PLEKHH2 | Tucker et al., 2021 | |
| | IGF1 | Tucker et al., 2021 | |
| | SCN7A | Poulsen et al., 2020 | Also neuronal marker |
| | COL1A2 | Cui et al., 2019 | |
| Myofibroblasts | DES | Wang et al., 2020 | Repeated in cardiomyocytes |
| | COL5A2 | Selewa et al., 2020 | |
| | ACTA2 | Tarbit et al., 2019 | Also smooth muscle |
| | PALLD | Tucker et al., 2021 | |
| Endothelial | PECAM1 | Lother et al., 2018; Litviňuková et al., 2020; Cui et al., 2019 | |
| | VWF | Lother et al., 2018; Litviňuková et al., 2020 | |
| | AQP1 | Wang et al., 2020 | |
| | CDH5 | Lother et al., 2018; Litviňuková et al., 2020 | |
| | TEK | Lother et al., 2018; Wang et al., 2020; Jia et al., 2018 | |
| | KDR | Lother et al., 2018; Wang et al., 2020 | |
| | SOX7 | Lother et al., 2018 | Venous |
| | NF2F2 | Lother et al., 2018 | Venous |
| Smooth Muscle | MYH11 | McLellan et al., 2020; Cui et al., 2019 | Classic |
| | ACTA2 | McLellan et al., 2020; Litviňuková et al., 2020; Wang et al., 2020 | Classic, repeated in myofibroblasts |
| | TAGLN | McLellan et al., 2020 | Classic |
| | CNN1 | Skelton et al., 2014 | |
| | OLFR558 | McLellan et al., 2020 | |
| | LMOD1 | McLellan et al., 2020 | |
| | NRIP2 | McLellan et al., 2020 | |
| | PI16 | Cui et al., 2019 | Immature |
| | CPZ | Cui et al., 2019 | Immature |
| | PDGFRL | Cui et al., 2019 | Immature |
| Pericytes | PDGFRB | Tucker et al., 2021; McLellan et al., 2020 | Smooth Muscle and Pericyte marker, repeated in smooth muscle |
| | PDGFRA | McLellan et al., 2020 | Repeated in fibroblasts |
| | COLEC11 | McLellan et al., 2020 | |
| | ABCC9 | McLellan et al., 2020 | |
| | KCNJ8 | McLellan et al., 2020 | |
| | VTN | McLellan et al., 2020 | |
| | STEAP4 | McLellan et al., 2020 | |
| | NOTCH3 | Wang et al., 2020; Litviňuková et al., 2020 | Smooth Muscle and Pericyte marker, repeated in smooth muscle |
| Lymphatic | LYVE1 | McLellan et al., 2020 | |
| | MMRN1 | McLellan et al., 2020 | |
| | CCL21A | McLellan et al., 2020 | |
| | CLDN5 | McLellan et al., 2020 | |
| | FLT1 | McLellan et al., 2020 | |
| Dendritic | CD11C | Novershtern et al., 2011; Merad et al., 2013 | |
| | HLA-DR | | |
| Neuronal | SCN7A | Tucker et al., 2021 | |
| | NOVA1 | Tucker et al., 2021 | |
| | VIM | Tucker et al., 2021 | Schwann, repeated in myofibroblasts, fibroblasts |
| | ANXA1 | Tucker et al., 2021 | Schwann |
| Leukocyte | MRC1 | Tucker et al., 2021 | |
| | FYB | Tucker et al., 2021 | |
| | CD86 | Tucker et al., 2021 | |
| Natural Killer | CD3 (not expressed) | Montaldo et al., 2013; Chen et al., 2015; Colucci et al., 2003; Farag &Caligiuri, 2006; Humana Press, 2007 | |
| | CD56 | | |
| | CD94 | | |
| | NKp46 | | |
| T | CD3 | Humana Press, 2007; Balon et al., 2006; Finak et al., 2016 | |
| B | CD19 | Humana Press, 2007; Kaminski et al., 2012; Orlic et al., 1993; Bendall et al., 2014; Wood, 2004 | |
| | CD79A | McLellan et al., 2020 | |
| | LY6D | McLellan et al., 2020 | |
| | H2-DMB2 | McLellan et al., 2020 | |
| | CD79B | McLellan et al., 2020 | |
| | MS4A1 | McLellan et al., 2020 | |
| Monocyte | CD14 | Novershtern et al., 2011; Wood, 2004; Yang et al., 2014 | |
| | PTPRC (CD45) | | |
| | CSFR3R | | |
| Macrophage | CD11B | Murray et al., 2011; Pilling et al., 2009 | |
| | CD68 | | |
| | CD163 | | |
| | CSF1R | McLellan et al., 2020 | |
| | ADGRE1 | McLellan et al., 2020 | |
| | PLD4 | McLellan et al., 2020 | |
| | MS4A6C | McLellan et al., 2020 | |
| | MGL2 | McLellan et al., 2020 | |
| Neutrophil | CD11B | Humana Press, 2007; Wood, 2004; Elghetany & Elghetany, 2002; Mantovani et al., 2011; Behnen et al., 2014 | |
| | CD16 | | |
| | CD18 | | |
| | CD32 | | |
| | CD55 | | |

Supplemental Table 2. Consensus Cell Identity Assignments for Each Cluster (blue)

| Cluster | *Cell Assignment Method* | | | |
|---|---|---|---|---|
| | **Biomarkers** | **pangloadb overexpressed genes** | **Gene Ontology** | **Inginuity Pathway Analysis** |
| **0** | Cardiomyocyte | Uknown | Cardiomyocyte | Cardiomyocyte |
| **1** | Cardiomyocyte | Unknown | Cardiomyocyte | Cardiomyocyte |
| **2** | Endothelial | Endothelial | Endothelial | Endothelial |
| **3** | Cardiomyocyte | Uknown | Cardiomyocyte | Cardiomyocyte |
| **4** | Fibroblast | Fibroblast | Unknown | Unknown |
| **5** | Fibroblast | Fibroblast | Fibroblast | Fibroblast |
| **6** | Leukocyte | Dendritic, Unknown | Leukocyte | Unknown |
| **7** | Cardiomyocyte | Unknown | Cardiomyocyte | Cardiomyocyte |
| **8** | Cardiomyocyte | Unknown | Cardiomyocyte | Cardiomyocyte |
| **9** | Cardiomyocyte | Unknown | Unknown | Cardiomyocyte |
| **10** | Pericyte | Unknown | Pericyte | Unknown |
| **11** | Dendritic, Leukocyte | Dendritic | Dendritic | Unknown |
| **12** | Fibroblast | Fibroblast | Cardiomyocyte | Unknown |
| **13** | Cardiomyocyte | Unknown | Cardiomyocyte | Cardiomyocyte |
| **14** | Cardiomyocyte | Unknown | Unknown | Cardiomyocyte |
| **15** | Smooth Muscle | Smooth Muscle | Smooth Muscle | Unknown |
| **16** | Cardiomyocyte, Neuronal | Uknown | Neuronal | Neuronal |
| **17** | Fibroblast | Fibroblast | Unknown | Fibroblast |
| **18** | Fibroblast | Fibroblast | Unknown | Unknown |
| **19** | Cardiomyocyte | Uknown | Unknown | Cardiomyocyte |
| **20** | Fibroblast | Fibroblast | Unknown | Unknown |
| **21** | Cardiomyocyte | Unknown | Cardiomyocyte | Unknown |

| Cell Type | Condition | *Number of* | | |
|---|---|---|---|---|
| | | **Nuclei** | **Differentially expressed genes** | **Gene overlap** |
| Cardiomyocyte | Normal | 16659 | 6695 | 4518 |
| | Non-obstructive | 26964 | 4851 | |
| Fibroblast | Normal | 9147 | 831 | 206 |
| | Non-obstructive | 9251 | 324 | |
| Endothelial | Normal | 2214 | 578 | 164 |
| | Non-obstructive | 5491 | 359 | |
| Pericyte | Normal | 1351 | 407 | 48 |
| | Non-obstructive | 2260 | 159 | |
| Dendritic | Normal | 1558 | 450 | 51 |
| | Non-obstructive | 1604 | 142 | |
| Leukocyte | Normal | 2457 | 226 | 25 |
| | Non-obstructive | 1754 | 136 | |
| Smooth Muscle | Normal | 563 | 250 | 33 |
| | Non-obstructive | 961 | 173 | |
| Neuronal | Normal | 409 | 295 | 18 |
| | Non-obstructive | 725 | 115 | |

**Supplemental Table 3**. Number of differentially expressed genes by cell type and condition. Differentially expressed genes were determined by positive Moran's I statistic values when respective adjusted p-values were $\leq 0.05$ and when genes were expressed in $\geq 1\%$ of all cells in a cell type. The number of cells per cell class and number of overlapping differentially expressed genes among cell classes are also listed.

Supplemental Table 4. Differentially Expressed Genes in Nonobstructive HCM

| | Filtered Differentially Expressed Genes Over Space | Cell Type |
|---|---|---|
| 1 | ABCA10 | Cardiomyocyte, Fibroblast |
| 2 | ABCA6 | Cardiomyocyte |
| 3 | ABCA8 | Cardiomyocyte |
| 4 | AC010680.1 | Cardiomyocyte |
| 5 | AC010680.5 | Cardiomyocyte |
| 7 | ADH1B | Cardiomyocyte |
| 8 | AGT | Endothelial, Pericyte |
| 11 | B2M | Dendritic, Neuronal |
| 12 | C1R | Fibroblast |
| 14 | CARMN | Smooth Muscle |
| 18 | CD69 | Dendritic |
| 20 | CLU | Endothelial |
| 21 | COL1A2 | Neuronal |
| 23 | DNM3OS | Cardiomyocyte |
| 24 | EDN1 | Endothelial |
| 25 | EMC10 | Cardiomyocyte |
| 27 | FBLN2 | Fibroblast |
| 28 | GZMA | Dendritic |
| 30 | HES4 | Smooth Muscle |
| 31 | HLA-DPA1 | Dendritic |
| 33 | HLA-DRB1 | Dendritic |
| 36 | IGFBP7 | Endothelial, Pericyte |
| 37 | LINC00861 | Dendritic |
| 38 | MEG3 | Cardiomyocyte, Fibroblast |
| 39 | MMRN1 | Endothelial |
| 40 | MS4A6A | Dendritic |
| 41 | MYH7 | Neuronal |
| 42 | MYH7B | Cardiomyocyte |
| 43 | MYL2 | Cardiomyocyte |
| 44 | NDUFA4L2 | Pericyte |
| 45 | NEBL | Cardiomyocyte,Neuronal |
| 46 | NPPB | Cardiomyocyte |
| 47 | PDGFRB | Pericyte, Smooth Muscle |
| 50 | RGS1 | Leukocyte |
| 51 | RGS5 | Endothelial, Pericyte |
| 52 | RP11-394O4.5 | Smooth Muscle |
| 53 | RP11-532N4.2 | Cardiomyocyte |
| 54 | SAT1 | Dendritic |
| 55 | SLC8A1 | Cardiomyocyte |
| 57 | SPARC | Neuronal |
| 58 | SPARCL1 | Pericyte |
| 59 | SPP1 | Neuronal |
| 60 | TFPI | Endothelial |
| 61 | TMSB4X | Endothelial |
| 63 | TTN | Neuronal |
| 64 | VIM | Neuronal |

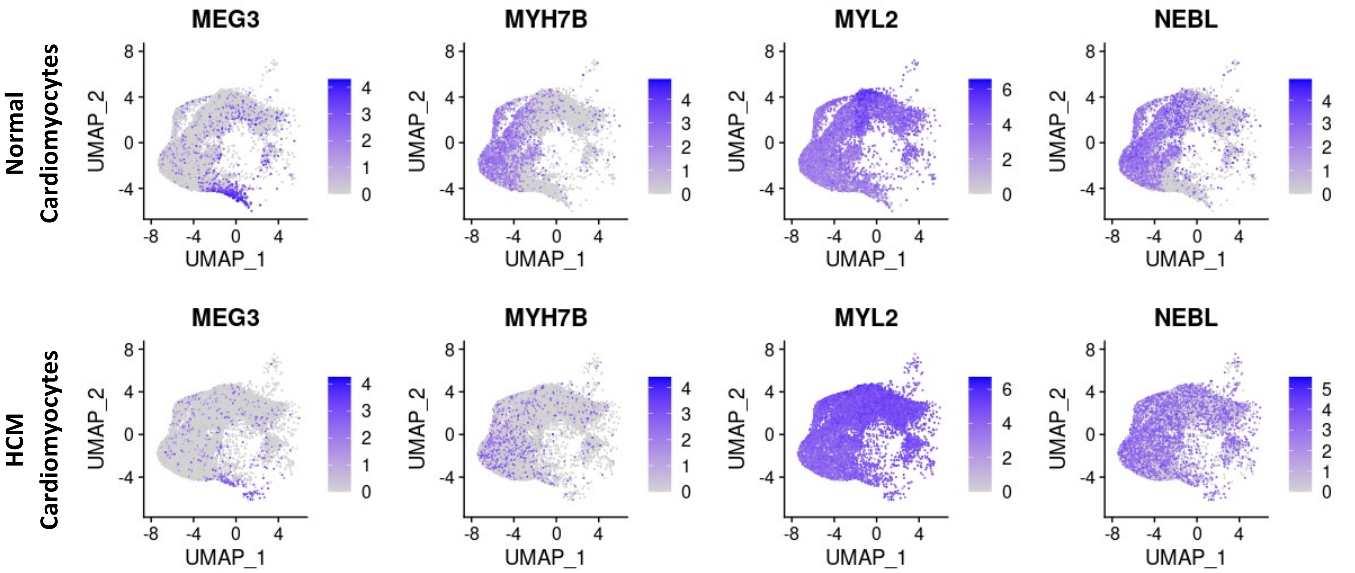


Supplemental Figure 1. Representative Differentially Expressed Genes in Cardiomyocytes, Determined by Spatial Autocorrelation, Plotted in UMAP Space

Supplemental Table 5. Increased Receptor Signaling to Dendritic Cells

| Normal | | | | |
|---|---|---|---|---|
| Pair_Name | Ligand | Receptor | L_cell | R_cell |
| COL1A2_CD36 | COL1A2 | CD36 | CM | DC |
| LGALS1_PTPRC | LGALS1 | PTPRC | CM | DC |
| TIMP1_CD63 | TIMP1 | CD63 | CM | DC |
| APP_CD74 | APP | CD74 | Fibro | DC |
| COL1A1_CD36 | COL1A1 | CD36 | Fibro | DC |
| COL1A2_CD36 | COL1A2 | CD36 | Fibro | DC |
| LGALS1_PTPRC | LGALS1 | PTPRC | Fibro | DC |
| TIMP1_CD63 | TIMP1 | CD63 | Fibro | DC |
| APP_CD74 | APP | CD74 | EC | DC |
| COL1A1_CD36 | COL1A1 | CD36 | EC | DC |
| COL1A2_CD36 | COL1A2 | CD36 | EC | DC |
| LGALS1_PTPRC | LGALS1 | PTPRC | EC | DC |
| TIMP1_CD63 | TIMP1 | CD63 | EC | DC |
| COL1A2_CD36 | COL1A2 | CD36 | PC | DC |
| LGALS1_PTPRC | LGALS1 | PTPRC | PC | DC |
| TIMP1_CD63 | TIMP1 | CD63 | PC | DC |
| COL1A1_CD36 | COL1A1 | CD36 | DC | DC |
| COL1A2_CD36 | COL1A2 | CD36 | DC | DC |
| LGALS1_PTPRC | LGALS1 | PTPRC | DC | DC |
| TIMP1_CD63 | TIMP1 | CD63 | DC | DC |
| COL1A1_CD36 | COL1A1 | CD36 | Leuko | DC |
| COL1A2_CD36 | COL1A2 | CD36 | Leuko | DC |
| LGALS1_PTPRC | LGALS1 | PTPRC | Leuko | DC |
| TIMP1_CD63 | TIMP1 | CD63 | Leuko | DC |
| COL1A2_CD36 | COL1A2 | CD36 | SMC | DC |
| LGALS1_PTPRC | LGALS1 | PTPRC | SMC | DC |
| TIMP1_CD63 | TIMP1 | CD63 | SMC | DC |
| COL1A2_CD36 | COL1A2 | CD36 | Neuro | DC |
| LGALS1_PTPRC | LGALS1 | PTPRC | Neuro | DC |
| TIMP1_CD63 | TIMP1 | CD63 | Neuro | DC |

| Non-obstructive | | | | |
|---|---|---|---|---|
| Pair_Name | Ligand | Receptor | L_cell | R_cell |
| CALM1_RYR2 | CALM1 | RYR2 | CM | DC |
| LAMA2_ITGB1 | LAMA2 | ITGB1 | CM | DC |
| LGALS1_ITGB1 | LGALS1 | ITGB1 | CM | DC |
| S100A1_RYR2 | S100A1 | RYR2 | CM | DC |
| TGM2_ITGB1 | TGM2 | ITGB1 | CM | DC |
| CALM1_RYR2 | CALM1 | RYR2 | Fibro | DC |
| COL1A2_CD36 | COL1A2 | CD36 | Fibro | DC |
| COL1A2_ITGB1 | COL1A2 | ITGB1 | Fibro | DC |
| COL3A1_ITGB1 | COL3A1 | ITGB1 | Fibro | DC |
| COL4A1_ITGB1 | COL4A1 | ITGB1 | Fibro | DC |
| COL6A1_ITGB1 | COL6A1 | ITGB1 | Fibro | DC |
| COL6A2_ITGB1 | COL6A2 | ITGB1 | Fibro | DC |
| COL6A3_ITGB1 | COL6A3 | ITGB1 | Fibro | DC |
| FN1_ITGB1 | FN1 | ITGB1 | Fibro | DC |
| LAMA2_ITGB1 | LAMA2 | ITGB1 | Fibro | DC |
| LGALS1_ITGB1 | LGALS1 | ITGB1 | Fibro | DC |
| LUM_ITGB1 | LUM | ITGB1 | Fibro | DC |
| S100A1_RYR2 | S100A1 | RYR2 | Fibro | DC |
| CALM1_RYR2 | CALM1 | RYR2 | EC | DC |
| FN1_ITGB1 | FN1 | ITGB1 | EC | DC |
| LGALS1_ITGB1 | LGALS1 | ITGB1 | EC | DC |
| S100A1_RYR2 | S100A1 | RYR2 | EC | DC |
| CALM1_RYR2 | CALM1 | RYR2 | PC | DC |
| COL1A2_CD36 | COL1A2 | CD36 | PC | DC |
| COL1A2_ITGB1 | COL1A2 | ITGB1 | PC | DC |
| COL4A1_ITGB1 | COL4A1 | ITGB1 | PC | DC |
| COL6A1_ITGB1 | COL6A1 | ITGB1 | PC | DC |
| FN1_ITGB1 | FN1 | ITGB1 | PC | DC |
| LGALS1_ITGB1 | LGALS1 | ITGB1 | PC | DC |
| S100A1_RYR2 | S100A1 | RYR2 | PC | DC |
| CALM1_RYR2 | CALM1 | RYR2 | DC | DC |
| LGALS1_ITGB1 | LGALS1 | ITGB1 | DC | DC |
| S100A1_RYR2 | S100A1 | RYR2 | DC | DC |
| CALM1_RYR2 | CALM1 | RYR2 | Leuko | DC |
| LGALS1_ITGB1 | LGALS1 | ITGB1 | Leuko | DC |
| LUM_ITGB1 | LUM | ITGB1 | Leuko | DC |
| S100A1_RYR2 | S100A1 | RYR2 | Leuko | DC |
| CALM1_RYR2 | CALM1 | RYR2 | SMC | DC |
| COL1A2_CD36 | COL1A2 | CD36 | SMC | DC |
| COL1A2_ITGB1 | COL1A2 | ITGB1 | SMC | DC |
| COL4A1_ITGB1 | COL4A1 | ITGB1 | SMC | DC |
| COL6A1_ITGB1 | COL6A1 | ITGB1 | SMC | DC |
| COL6A2_ITGB1 | COL6A2 | ITGB1 | SMC | DC |
| FN1_ITGB1 | FN1 | ITGB1 | SMC | DC |
| LGALS1_ITGB1 | LGALS1 | ITGB1 | SMC | DC |
| S100A1_RYR2 | S100A1 | RYR2 | SMC | DC |
| CALM1_RYR2 | CALM1 | RYR2 | Neuro | DC |
| LGALS1_ITGB1 | LGALS1 | ITGB1 | Neuro | DC |
| S100A1_RYR2 | S100A1 | RYR2 | Neuro | DC |

Supplemental Table 6. Increased Smooth Muscle to Leukocyte Communication in Nonobstructive HCM

| ***Normal*** | | | | |
|---|---|---|---|---|
| **Pair_Name** | **Ligand** | **Receptor** | **L_cell** | **R_cell** |
| A2M_LRP1 | A2M | LRP1 | SMC | Leuko |
| COL1A2_CD36 | COL1A2 | CD36 | SMC | Leuko |
| HMGB1_CD163 | HMGB1 | CD163 | SMC | Leuko |
| LGALS1_PTPRC | LGALS1 | PTPRC | SMC | Leuko |
| PSAP_LRP1 | PSAP | LRP1 | SMC | Leuko |
| TIMP1_CD63 | TIMP1 | CD63 | SMC | Leuko |

| ***Non-obstructive*** | | | | |
|---|---|---|---|---|
| **Pair_Name** | **Ligand** | **Receptor** | **L_cell** | **R_cell** |
| CALM1_RYR2 | CALM1 | RYR2 | SMC | Leuko |
| COL1A2_CD36 | COL1A2 | CD36 | SMC | Leuko |
| COL1A2_ITGB1 | COL1A2 | ITGB1 | SMC | Leuko |
| COL4A1_ITGB1 | COL4A1 | ITGB1 | SMC | Leuko |
| COL6A1_ITGB1 | COL6A1 | ITGB1 | SMC | Leuko |
| COL6A2_ITGB1 | COL6A2 | ITGB1 | SMC | Leuko |
| FN1_ITGB1 | FN1 | ITGB1 | SMC | Leuko |
| HMGB1_CD163 | HMGB1 | CD163 | SMC | Leuko |
| LGALS1_ITGB1 | LGALS1 | ITGB1 | SMC | Leuko |
| S100A1_RYR2 | S100A1 | RYR2 | SMC | Leuko |

### Supplemental Table 7. Fibroblast Cluster 2 Communication with Fibroblast Clusters 2-6

| *Normal* | | | | |
|---|---|---|---|---|
| **Pair_Name** | **Ligand** | **Receptor** | **L_cell** | **R_cell** |
| APP_APLP2 | APP | APLP2 | Fibro 2 | Fibro 2 |
| APP_LRP1 | APP | LRP1 | Fibro 2 | Fibro 2 |
| C3_CD81 | C3 | CD81 | Fibro 2 | Fibro 2 |
| C3_LRP1 | C3 | LRP1 | Fibro 2 | Fibro 2 |
| CALR_LRP1 | CALR | LRP1 | Fibro 2 | Fibro 2 |
| CD99_CD81 | CD99 | CD81 | Fibro 2 | Fibro 2 |
| COL18A1_ITGB1 | COL18A1 | ITGB1 | Fibro 2 | Fibro 2 |
| COL1A1_ITGB1 | COL1A1 | ITGB1 | Fibro 2 | Fibro 2 |
| COL1A2_ITGB1 | COL1A2 | ITGB1 | Fibro 2 | Fibro 2 |
| COL3A1_ITGB1 | COL3A1 | ITGB1 | Fibro 2 | Fibro 2 |
| COL4A1_ITGB1 | COL4A1 | ITGB1 | Fibro 2 | Fibro 2 |
| COL5A1_ITGB1 | COL5A1 | ITGB1 | Fibro 2 | Fibro 2 |
| COL5A2_ITGB1 | COL5A2 | ITGB1 | Fibro 2 | Fibro 2 |
| COL6A1_ITGB1 | COL6A1 | ITGB1 | Fibro 2 | Fibro 2 |
| COL6A2_ITGB1 | COL6A2 | ITGB1 | Fibro 2 | Fibro 2 |
| COL6A3_ITGB1 | COL6A3 | ITGB1 | Fibro 2 | Fibro 2 |
| CTGF_LRP1 | CTGF | LRP1 | Fibro 2 | Fibro 2 |
| FBLN1_ITGB1 | FBLN1 | ITGB1 | Fibro 2 | Fibro 2 |
| FBN1_ITGB1 | FBN1 | ITGB1 | Fibro 2 | Fibro 2 |
| FN1_ITGB1 | FN1 | ITGB1 | Fibro 2 | Fibro 2 |
| FN1_SDC2 | FN1 | SDC2 | Fibro 2 | Fibro 2 |
| HLA-A_APLP2 | HLA-A | APLP2 | Fibro 2 | Fibro 2 |
| HSP90AA1_LRP1 | HSP90AA1 | LRP1 | Fibro 2 | Fibro 2 |
| HSP90B1_LRP1 | HSP90B1 | LRP1 | Fibro 2 | Fibro 2 |
| HSPG2_ITGB1 | HSPG2 | ITGB1 | Fibro 2 | Fibro 2 |
| HSPG2_LRP1 | HSPG2 | LRP1 | Fibro 2 | Fibro 2 |
| LAMA2_ITGB1 | LAMA2 | ITGB1 | Fibro 2 | Fibro 2 |
| LAMB1_ITGB1 | LAMB1 | ITGB1 | Fibro 2 | Fibro 2 |
| LAMC1_ITGB1 | LAMC1 | ITGB1 | Fibro 2 | Fibro 2 |
| LGALS1_ITGB1 | LGALS1 | ITGB1 | Fibro 2 | Fibro 2 |
| LGALS3BP_ITGB1 | LGALS3BP | ITGB1 | Fibro 2 | Fibro 2 |
| LRPAP1_LRP1 | LRPAP1 | LRP1 | Fibro 2 | Fibro 2 |
| LUM_ITGB1 | LUM | ITGB1 | Fibro 2 | Fibro 2 |
| MFGE8_PDGFRB | MFGE8 | PDGFRB | Fibro 2 | Fibro 2 |
| MMP2_SDC2 | MMP2 | SDC2 | Fibro 2 | Fibro 2 |
| NID1_ITGB1 | NID1 | ITGB1 | Fibro 2 | Fibro 2 |
| PSAP_LRP1 | PSAP | LRP1 | Fibro 2 | Fibro 2 |
| SERPINE2_LRP1 | SERPINE2 | LRP1 | Fibro 2 | Fibro 2 |
| SERPING1_LRP1 | SERPING1 | LRP1 | Fibro 2 | Fibro 2 |
| TFPI_LRP1 | TFPI | LRP1 | Fibro 2 | Fibro 2 |
| THBS2_ITGB1 | THBS2 | ITGB1 | Fibro 2 | Fibro 2 |
| TIMP1_CD63 | TIMP1 | CD63 | Fibro 2 | Fibro 2 |
| TIMP2_ITGB1 | TIMP2 | ITGB1 | Fibro 2 | Fibro 2 |
| VCAN_ITGB1 | VCAN | ITGB1 | Fibro 2 | Fibro 2 |
| APP_LRP1 | APP | LRP1 | Fibro 2 | Fibro 3 |
| C3_CD81 | C3 | CD81 | Fibro 2 | Fibro 3 |
| C3_LRP1 | C3 | LRP1 | Fibro 2 | Fibro 3 |
| CALR_LRP1 | CALR | LRP1 | Fibro 2 | Fibro 3 |
| CD99_CD81 | CD99 | CD81 | Fibro 2 | Fibro 3 |
| COL18A1_ITGB1 | COL18A1 | ITGB1 | Fibro 2 | Fibro 3 |
| COL1A1_ITGB1 | COL1A1 | ITGB1 | Fibro 2 | Fibro 3 |
| COL1A2_ITGB1 | COL1A2 | ITGB1 | Fibro 2 | Fibro 3 |
| COL3A1_ITGB1 | COL3A1 | ITGB1 | Fibro 2 | Fibro 3 |
| COL4A1_ITGB1 | COL4A1 | ITGB1 | Fibro 2 | Fibro 3 |
| COL5A1_ITGB1 | COL5A1 | ITGB1 | Fibro 2 | Fibro 3 |
| COL5A2_ITGB1 | COL5A2 | ITGB1 | Fibro 2 | Fibro 3 |
| COL6A1_ITGB1 | COL6A1 | ITGB1 | Fibro 2 | Fibro 3 |
| COL6A2_ITGB1 | COL6A2 | ITGB1 | Fibro 2 | Fibro 3 |
| COL6A3_ITGB1 | COL6A3 | ITGB1 | Fibro 2 | Fibro 3 |
| CTGF_LRP1 | CTGF | LRP1 | Fibro 2 | Fibro 3 |
| FBLN1_ITGB1 | FBLN1 | ITGB1 | Fibro 2 | Fibro 3 |
| FBN1_ITGB1 | FBN1 | ITGB1 | Fibro 2 | Fibro 3 |
| FN1_ITGB1 | FN1 | ITGB1 | Fibro 2 | Fibro 3 |
| HSP90AA1_LRP1 | HSP90AA1 | LRP1 | Fibro 2 | Fibro 3 |
| HSP90B1_LRP1 | HSP90B1 | LRP1 | Fibro 2 | Fibro 3 |
| HSPG2_ITGB1 | HSPG2 | ITGB1 | Fibro 2 | Fibro 3 |
| HSPG2_LRP1 | HSPG2 | LRP1 | Fibro 2 | Fibro 3 |
| LAMA2_ITGB1 | LAMA2 | ITGB1 | Fibro 2 | Fibro 3 |
| LAMB1_ITGB1 | LAMB1 | ITGB1 | Fibro 2 | Fibro 3 |
| LAMC1_ITGB1 | LAMC1 | ITGB1 | Fibro 2 | Fibro 3 |
| LGALS1_ITGB1 | LGALS1 | ITGB1 | Fibro 2 | Fibro 3 |
| LGALS3BP_ITGB1 | LGALS3BP | ITGB1 | Fibro 2 | Fibro 3 |
| LRPAP1_LRP1 | LRPAP1 | LRP1 | Fibro 2 | Fibro 3 |
| LUM_ITGB1 | LUM | ITGB1 | Fibro 2 | Fibro 3 |
| MFGE8_PDGFRB | MFGE8 | PDGFRB | Fibro 2 | Fibro 3 |
| NID1_ITGB1 | NID1 | ITGB1 | Fibro 2 | Fibro 3 |
| PSAP_LRP1 | PSAP | LRP1 | Fibro 2 | Fibro 3 |
| SERPINE2_LRP1 | SERPINE2 | LRP1 | Fibro 2 | Fibro 3 |
| SERPING1_LRP1 | SERPING1 | LRP1 | Fibro 2 | Fibro 3 |
| TFPI_LRP1 | TFPI | LRP1 | Fibro 2 | Fibro 3 |
| THBS2_ITGB1 | THBS2 | ITGB1 | Fibro 2 | Fibro 3 |
| TIMP1_CD63 | TIMP1 | CD63 | Fibro 2 | Fibro 3 |
| TIMP2_ITGB1 | TIMP2 | ITGB1 | Fibro 2 | Fibro 3 |
| VCAN_ITGB1 | VCAN | ITGB1 | Fibro 2 | Fibro 3 |
| APP_LRP1 | APP | LRP1 | Fibro 2 | Fibro 4 |
| C3_CD81 | C3 | CD81 | Fibro 2 | Fibro 4 |
| C3_LRP1 | C3 | LRP1 | Fibro 2 | Fibro 4 |
| CALR_LRP1 | CALR | LRP1 | Fibro 2 | Fibro 4 |
| CD99_CD81 | CD99 | CD81 | Fibro 2 | Fibro 4 |
| COL18A1_ITGB1 | COL18A1 | ITGB1 | Fibro 2 | Fibro 4 |
| COL1A1_ITGB1 | COL1A1 | ITGB1 | Fibro 2 | Fibro 4 |
| COL1A2_ITGB1 | COL1A2 | ITGB1 | Fibro 2 | Fibro 4 |
| COL3A1_ITGB1 | COL3A1 | ITGB1 | Fibro 2 | Fibro 4 |
| COL4A1_ITGB1 | COL4A1 | ITGB1 | Fibro 2 | Fibro 4 |
| COL5A1_ITGB1 | COL5A1 | ITGB1 | Fibro 2 | Fibro 4 |
| COL5A2_ITGB1 | COL5A2 | ITGB1 | Fibro 2 | Fibro 4 |
| COL6A1_ITGB1 | COL6A1 | ITGB1 | Fibro 2 | Fibro 4 |
| COL6A2_ITGB1 | COL6A2 | ITGB1 | Fibro 2 | Fibro 4 |
| COL6A3_ITGB1 | COL6A3 | ITGB1 | Fibro 2 | Fibro 4 |
| CTGF_LRP1 | CTGF | LRP1 | Fibro 2 | Fibro 4 |
| FBLN1_ITGB1 | FBLN1 | ITGB1 | Fibro 2 | Fibro 4 |
| FBN1_ITGB1 | FBN1 | ITGB1 | Fibro 2 | Fibro 4 |
| FN1_ITGB1 | FN1 | ITGB1 | Fibro 2 | Fibro 4 |
| HSP90AA1_LRP1 | HSP90AA1 | LRP1 | Fibro 2 | Fibro 4 |
| HSP90B1_LRP1 | HSP90B1 | LRP1 | Fibro 2 | Fibro 4 |
| HSPG2_ITGB1 | HSPG2 | ITGB1 | Fibro 2 | Fibro 4 |
| HSPG2_LRP1 | HSPG2 | LRP1 | Fibro 2 | Fibro 4 |
| LAMA2_ITGB1 | LAMA2 | ITGB1 | Fibro 2 | Fibro 4 |
| LAMB1_ITGB1 | LAMB1 | ITGB1 | Fibro 2 | Fibro 4 |
| LAMC1_ITGB1 | LAMC1 | ITGB1 | Fibro 2 | Fibro 4 |
| LGALS1_ITGB1 | LGALS1 | ITGB1 | Fibro 2 | Fibro 4 |
| LGALS3BP_ITGB1 | LGALS3BP | ITGB1 | Fibro 2 | Fibro 4 |
| LRPAP1_LRP1 | LRPAP1 | LRP1 | Fibro 2 | Fibro 4 |
| LUM_ITGB1 | LUM | ITGB1 | Fibro 2 | Fibro 4 |
| MFGE8_PDGFRB | MFGE8 | PDGFRB | Fibro 2 | Fibro 4 |
| NID1_ITGB1 | NID1 | ITGB1 | Fibro 2 | Fibro 4 |
| PSAP_LRP1 | PSAP | LRP1 | Fibro 2 | Fibro 4 |
| SERPINE2_LRP1 | SERPINE2 | LRP1 | Fibro 2 | Fibro 4 |
| SERPING1_LRP1 | SERPING1 | LRP1 | Fibro 2 | Fibro 4 |
| TFPI_LRP1 | TFPI | LRP1 | Fibro 2 | Fibro 4 |
| THBS2_ITGB1 | THBS2 | ITGB1 | Fibro 2 | Fibro 4 |
| TIMP1_CD63 | TIMP1 | CD63 | Fibro 2 | Fibro 4 |
| TIMP2_ITGB1 | TIMP2 | ITGB1 | Fibro 2 | Fibro 4 |
| VCAN_ITGB1 | VCAN | ITGB1 | Fibro 2 | Fibro 4 |
| APP_LRP1 | APP | LRP1 | Fibro 2 | Fibro 5 |
| C3_CD81 | C3 | CD81 | Fibro 2 | Fibro 5 |
| C3_LRP1 | C3 | LRP1 | Fibro 2 | Fibro 5 |
| CALR_LRP1 | CALR | LRP1 | Fibro 2 | Fibro 5 |
| CD99_CD81 | CD99 | CD81 | Fibro 2 | Fibro 5 |
| COL18A1_ITGB1 | COL18A1 | ITGB1 | Fibro 2 | Fibro 5 |
| COL1A1_CD36 | COL1A1 | CD36 | Fibro 2 | Fibro 5 |
| COL1A1_ITGB1 | COL1A1 | ITGB1 | Fibro 2 | Fibro 5 |
| COL1A2_CD36 | COL1A2 | CD36 | Fibro 2 | Fibro 5 |
| COL1A2_ITGB1 | COL1A2 | ITGB1 | Fibro 2 | Fibro 5 |
| COL3A1_ITGB1 | COL3A1 | ITGB1 | Fibro 2 | Fibro 5 |
| COL4A1_ITGB1 | COL4A1 | ITGB1 | Fibro 2 | Fibro 5 |
| COL5A1_ITGB1 | COL5A1 | ITGB1 | Fibro 2 | Fibro 5 |
| COL5A2_ITGB1 | COL5A2 | ITGB1 | Fibro 2 | Fibro 5 |
| COL6A1_ITGB1 | COL6A1 | ITGB1 | Fibro 2 | Fibro 5 |
| COL6A2_ITGB1 | COL6A2 | ITGB1 | Fibro 2 | Fibro 5 |
| COL6A3_ITGB1 | COL6A3 | ITGB1 | Fibro 2 | Fibro 5 |
| CTGF_LRP1 | CTGF | LRP1 | Fibro 2 | Fibro 5 |
| FBLN1_ITGB1 | FBLN1 | ITGB1 | Fibro 2 | Fibro 5 |
| FBN1_ITGB1 | FBN1 | ITGB1 | Fibro 2 | Fibro 5 |
| FN1_ITGB1 | FN1 | ITGB1 | Fibro 2 | Fibro 5 |
| HSP90AA1_LRP1 | HSP90AA1 | LRP1 | Fibro 2 | Fibro 5 |
| HSP90B1_LRP1 | HSP90B1 | LRP1 | Fibro 2 | Fibro 5 |
| HSPG2_ITGB1 | HSPG2 | ITGB1 | Fibro 2 | Fibro 5 |
| HSPG2_LRP1 | HSPG2 | LRP1 | Fibro 2 | Fibro 5 |
| LAMA2_ITGB1 | LAMA2 | ITGB1 | Fibro 2 | Fibro 5 |
| LAMB1_ITGB1 | LAMB1 | ITGB1 | Fibro 2 | Fibro 5 |
| LAMC1_ITGB1 | LAMC1 | ITGB1 | Fibro 2 | Fibro 5 |
| LGALS1_ITGB1 | LGALS1 | ITGB1 | Fibro 2 | Fibro 5 |
| LGALS3BP_ITGB1 | LGALS3BP | ITGB1 | Fibro 2 | Fibro 5 |
| LRPAP1_LRP1 | LRPAP1 | LRP1 | Fibro 2 | Fibro 5 |
| LUM_ITGB1 | LUM | ITGB1 | Fibro 2 | Fibro 5 |
| MFGE8_PDGFRB | MFGE8 | PDGFRB | Fibro 2 | Fibro 5 |
| NID1_ITGB1 | NID1 | ITGB1 | Fibro 2 | Fibro 5 |
| PSAP_LRP1 | PSAP | LRP1 | Fibro 2 | Fibro 5 |
| SERPINE2_LRP1 | SERPINE2 | LRP1 | Fibro 2 | Fibro 5 |
| SERPING1_LRP1 | SERPING1 | LRP1 | Fibro 2 | Fibro 5 |
| TFPI_LRP1 | TFPI | LRP1 | Fibro 2 | Fibro 5 |
| THBS2_CD36 | THBS2 | CD36 | Fibro 2 | Fibro 5 |
| THBS2_ITGB1 | THBS2 | ITGB1 | Fibro 2 | Fibro 5 |
| TIMP1_CD63 | TIMP1 | CD63 | Fibro 2 | Fibro 5 |
| TIMP2_ITGB1 | TIMP2 | ITGB1 | Fibro 2 | Fibro 5 |
| VCAN_ITGB1 | VCAN | ITGB1 | Fibro 2 | Fibro 5 |
| APP_LRP1 | APP | LRP1 | Fibro 2 | Fibro 6 |
| C3_CD81 | C3 | CD81 | Fibro 2 | Fibro 6 |
| C3_LRP1 | C3 | LRP1 | Fibro 2 | Fibro 6 |
| CALR_LRP1 | CALR | LRP1 | Fibro 2 | Fibro 6 |
| CD99_CD81 | CD99 | CD81 | Fibro 2 | Fibro 6 |
| COL18A1_ITGB1 | COL18A1 | ITGB1 | Fibro 2 | Fibro 6 |
| COL1A1_ITGB1 | COL1A1 | ITGB1 | Fibro 2 | Fibro 6 |
| COL1A2_ITGB1 | COL1A2 | ITGB1 | Fibro 2 | Fibro 6 |
| COL3A1_ITGB1 | COL3A1 | ITGB1 | Fibro 2 | Fibro 6 |
| COL4A1_ITGB1 | COL4A1 | ITGB1 | Fibro 2 | Fibro 6 |
| COL5A1_ITGB1 | COL5A1 | ITGB1 | Fibro 2 | Fibro 6 |
| COL5A2_ITGB1 | COL5A2 | ITGB1 | Fibro 2 | Fibro 6 |
| COL6A1_ITGB1 | COL6A1 | ITGB1 | Fibro 2 | Fibro 6 |
| COL6A2_ITGB1 | COL6A2 | ITGB1 | Fibro 2 | Fibro 6 |
| COL6A3_ITGB1 | COL6A3 | ITGB1 | Fibro 2 | Fibro 6 |
| CTGF_LRP1 | CTGF | LRP1 | Fibro 2 | Fibro 6 |
| FBLN1_ITGB1 | FBLN1 | ITGB1 | Fibro 2 | Fibro 6 |
| FBN1_ITGB1 | FBN1 | ITGB1 | Fibro 2 | Fibro 6 |
| FN1_ITGB1 | FN1 | ITGB1 | Fibro 2 | Fibro 6 |
| HSP90AA1_LRP1 | HSP90AA1 | LRP1 | Fibro 2 | Fibro 6 |
| HSP90B1_LRP1 | HSP90B1 | LRP1 | Fibro 2 | Fibro 6 |
| HSPG2_ITGB1 | HSPG2 | ITGB1 | Fibro 2 | Fibro 6 |
| HSPG2_LRP1 | HSPG2 | LRP1 | Fibro 2 | Fibro 6 |
| LAMA2_ITGB1 | LAMA2 | ITGB1 | Fibro 2 | Fibro 6 |
| LAMB1_ITGB1 | LAMB1 | ITGB1 | Fibro 2 | Fibro 6 |
| LAMC1_ITGB1 | LAMC1 | ITGB1 | Fibro 2 | Fibro 6 |
| LGALS1_ITGB1 | LGALS1 | ITGB1 | Fibro 2 | Fibro 6 |
| LGALS3BP_ITGB1 | LGALS3BP | ITGB1 | Fibro 2 | Fibro 6 |
| LRPAP1_LRP1 | LRPAP1 | LRP1 | Fibro 2 | Fibro 6 |
| LUM_ITGB1 | LUM | ITGB1 | Fibro 2 | Fibro 6 |
| MFGE8_PDGFRB | MFGE8 | PDGFRB | Fibro 2 | Fibro 6 |
| NID1_ITGB1 | NID1 | ITGB1 | Fibro 2 | Fibro 6 |
| PSAP_LRP1 | PSAP | LRP1 | Fibro 2 | Fibro 6 |
| SERPINE2_LRP1 | SERPINE2 | LRP1 | Fibro 2 | Fibro 6 |
| SERPING1_LRP1 | SERPING1 | LRP1 | Fibro 2 | Fibro 6 |
| TFPI_LRP1 | TFPI | LRP1 | Fibro 2 | Fibro 6 |
| THBS2_ITGB1 | THBS2 | ITGB1 | Fibro 2 | Fibro 6 |
| TIMP1_CD63 | TIMP1 | CD63 | Fibro 2 | Fibro 6 |
| TIMP2_ITGB1 | TIMP2 | ITGB1 | Fibro 2 | Fibro 6 |
| VCAN_ITGB1 | VCAN | ITGB1 | Fibro 2 | Fibro 6 |

| *Non-obstructive* | | | | |
|---|---|---|---|---|
| **Pair_Name** | **Ligand** | **Receptor** | **L_cell** | **R_cell** |
| A2M_LRP1 | A2M | LRP1 | Fibro 2 | Fibro 2 |
| CALM1_RYR2 | CALM1 | RYR2 | Fibro 2 | Fibro 2 |
| CCN2_LRP1 | CCN2 | LRP1 | Fibro 2 | Fibro 2 |
| COL1A1_CD36 | COL1A1 | CD36 | Fibro 2 | Fibro 2 |
| COL1A1_ITGB1 | COL1A1 | ITGB1 | Fibro 2 | Fibro 2 |
| COL1A2_CD36 | COL1A2 | CD36 | Fibro 2 | Fibro 2 |
| COL1A2_ITGB1 | COL1A2 | ITGB1 | Fibro 2 | Fibro 2 |
| COL3A1_ITGB1 | COL3A1 | ITGB1 | Fibro 2 | Fibro 2 |
| COL4A1_ITGB1 | COL4A1 | ITGB1 | Fibro 2 | Fibro 2 |
| COL6A1_ITGB1 | COL6A1 | ITGB1 | Fibro 2 | Fibro 2 |
| COL6A2_ITGB1 | COL6A2 | ITGB1 | Fibro 2 | Fibro 2 |
| COL6A3_ITGB1 | COL6A3 | ITGB1 | Fibro 2 | Fibro 2 |
| FN1_ITGB1 | FN1 | ITGB1 | Fibro 2 | Fibro 2 |
| HSP90AA1_LRP1 | HSP90AA1 | LRP1 | Fibro 2 | Fibro 2 |
| HSP90B1_LRP1 | HSP90B1 | LRP1 | Fibro 2 | Fibro 2 |
| LAMA2_ITGB1 | LAMA2 | ITGB1 | Fibro 2 | Fibro 2 |
| LAMB1_ITGB1 | LAMB1 | ITGB1 | Fibro 2 | Fibro 2 |
| LGALS1_ITGB1 | LGALS1 | ITGB1 | Fibro 2 | Fibro 2 |
| LPL_LRP1 | LPL | LRP1 | Fibro 2 | Fibro 2 |
| LUM_ITGB1 | LUM | ITGB1 | Fibro 2 | Fibro 2 |
| PSAP_LRP1 | PSAP | LRP1 | Fibro 2 | Fibro 2 |
| S100A1_RYR2 | S100A1 | RYR2 | Fibro 2 | Fibro 2 |
| TIMP1_CD63 | TIMP1 | CD63 | Fibro 2 | Fibro 2 |
| TIMP2_ITGB1 | TIMP2 | ITGB1 | Fibro 2 | Fibro 2 |
| VCAN_ITGB1 | VCAN | ITGB1 | Fibro 2 | Fibro 2 |
| CALM1_RYR2 | CALM1 | RYR2 | Fibro 2 | Fibro 3 |
| COL1A1_CD36 | COL1A1 | CD36 | Fibro 2 | Fibro 3 |
| COL1A1_ITGB1 | COL1A1 | ITGB1 | Fibro 2 | Fibro 3 |
| COL1A2_CD36 | COL1A2 | CD36 | Fibro 2 | Fibro 3 |
| COL1A2_ITGB1 | COL1A2 | ITGB1 | Fibro 2 | Fibro 3 |
| COL3A1_ITGB1 | COL3A1 | ITGB1 | Fibro 2 | Fibro 3 |
| COL4A1_ITGB1 | COL4A1 | ITGB1 | Fibro 2 | Fibro 3 |
| COL6A1_ITGB1 | COL6A1 | ITGB1 | Fibro 2 | Fibro 3 |
| COL6A2_ITGB1 | COL6A2 | ITGB1 | Fibro 2 | Fibro 3 |
| COL6A3_ITGB1 | COL6A3 | ITGB1 | Fibro 2 | Fibro 3 |
| FN1_ITGB1 | FN1 | ITGB1 | Fibro 2 | Fibro 3 |
| LAMA2_ITGB1 | LAMA2 | ITGB1 | Fibro 2 | Fibro 3 |
| LAMB1_ITGB1 | LAMB1 | ITGB1 | Fibro 2 | Fibro 3 |
| LGALS1_ITGB1 | LGALS1 | ITGB1 | Fibro 2 | Fibro 3 |
| LUM_ITGB1 | LUM | ITGB1 | Fibro 2 | Fibro 3 |
| S100A1_RYR2 | S100A1 | RYR2 | Fibro 2 | Fibro 3 |
| TIMP1_CD63 | TIMP1 | CD63 | Fibro 2 | Fibro 3 |
| TIMP2_ITGB1 | TIMP2 | ITGB1 | Fibro 2 | Fibro 3 |
| VCAN_ITGB1 | VCAN | ITGB1 | Fibro 2 | Fibro 3 |
| CALM1_RYR2 | CALM1 | RYR2 | Fibro 2 | Fibro 4 |
| COL1A1_CD36 | COL1A1 | CD36 | Fibro 2 | Fibro 4 |
| COL1A1_ITGB1 | COL1A1 | ITGB1 | Fibro 2 | Fibro 4 |
| COL1A2_CD36 | COL1A2 | CD36 | Fibro 2 | Fibro 4 |
| COL1A2_ITGB1 | COL1A2 | ITGB1 | Fibro 2 | Fibro 4 |
| COL3A1_ITGB1 | COL3A1 | ITGB1 | Fibro 2 | Fibro 4 |
| COL4A1_ITGB1 | COL4A1 | ITGB1 | Fibro 2 | Fibro 4 |
| COL6A1_ITGB1 | COL6A1 | ITGB1 | Fibro 2 | Fibro 4 |
| COL6A2_ITGB1 | COL6A2 | ITGB1 | Fibro 2 | Fibro 4 |
| COL6A3_ITGB1 | COL6A3 | ITGB1 | Fibro 2 | Fibro 4 |
| FN1_ITGB1 | FN1 | ITGB1 | Fibro 2 | Fibro 4 |
| LAMA2_ITGB1 | LAMA2 | ITGB1 | Fibro 2 | Fibro 4 |
| LAMB1_ITGB1 | LAMB1 | ITGB1 | Fibro 2 | Fibro 4 |
| LGALS1_ITGB1 | LGALS1 | ITGB1 | Fibro 2 | Fibro 4 |
| LUM_ITGB1 | LUM | ITGB1 | Fibro 2 | Fibro 4 |
| S100A1_RYR2 | S100A1 | RYR2 | Fibro 2 | Fibro 4 |
| TIMP1_CD63 | TIMP1 | CD63 | Fibro 2 | Fibro 4 |
| TIMP2_ITGB1 | TIMP2 | ITGB1 | Fibro 2 | Fibro 4 |
| VCAN_ITGB1 | VCAN | ITGB1 | Fibro 2 | Fibro 4 |
| CALM1_RYR2 | CALM1 | RYR2 | Fibro 2 | Fibro 5 |
| COL1A1_CD36 | COL1A1 | CD36 | Fibro 2 | Fibro 5 |
| COL1A1_ITGB1 | COL1A1 | ITGB1 | Fibro 2 | Fibro 5 |
| COL1A2_CD36 | COL1A2 | CD36 | Fibro 2 | Fibro 5 |
| COL1A2_ITGB1 | COL1A2 | ITGB1 | Fibro 2 | Fibro 5 |
| COL3A1_ITGB1 | COL3A1 | ITGB1 | Fibro 2 | Fibro 5 |
| COL4A1_ITGB1 | COL4A1 | ITGB1 | Fibro 2 | Fibro 5 |
| COL6A1_ITGB1 | COL6A1 | ITGB1 | Fibro 2 | Fibro 5 |
| COL6A2_ITGB1 | COL6A2 | ITGB1 | Fibro 2 | Fibro 5 |
| COL6A3_ITGB1 | COL6A3 | ITGB1 | Fibro 2 | Fibro 5 |
| FN1_ITGB1 | FN1 | ITGB1 | Fibro 2 | Fibro 5 |
| LAMA2_ITGB1 | LAMA2 | ITGB1 | Fibro 2 | Fibro 5 |
| LAMB1_ITGB1 | LAMB1 | ITGB1 | Fibro 2 | Fibro 5 |
| LGALS1_ITGB1 | LGALS1 | ITGB1 | Fibro 2 | Fibro 5 |
| LUM_ITGB1 | LUM | ITGB1 | Fibro 2 | Fibro 5 |
| S100A1_RYR2 | S100A1 | RYR2 | Fibro 2 | Fibro 5 |
| TIMP1_CD63 | TIMP1 | CD63 | Fibro 2 | Fibro 5 |
| TIMP2_ITGB1 | TIMP2 | ITGB1 | Fibro 2 | Fibro 5 |
| VCAN_ITGB1 | VCAN | ITGB1 | Fibro 2 | Fibro 5 |
| CALM1_RYR2 | CALM1 | RYR2 | Fibro 2 | Fibro 6 |
| COL1A1_CD36 | COL1A1 | CD36 | Fibro 2 | Fibro 6 |
| COL1A1_ITGB1 | COL1A1 | ITGB1 | Fibro 2 | Fibro 6 |
| COL1A2_CD36 | COL1A2 | CD36 | Fibro 2 | Fibro 6 |
| COL1A2_ITGB1 | COL1A2 | ITGB1 | Fibro 2 | Fibro 6 |
| COL3A1_ITGB1 | COL3A1 | ITGB1 | Fibro 2 | Fibro 6 |
| COL4A1_ITGB1 | COL4A1 | ITGB1 | Fibro 2 | Fibro 6 |
| COL6A1_ITGB1 | COL6A1 | ITGB1 | Fibro 2 | Fibro 6 |
| COL6A2_ITGB1 | COL6A2 | ITGB1 | Fibro 2 | Fibro 6 |
| COL6A3_ITGB1 | COL6A3 | ITGB1 | Fibro 2 | Fibro 6 |
| FN1_ITGB1 | FN1 | ITGB1 | Fibro 2 | Fibro 6 |
| LAMA2_ITGB1 | LAMA2 | ITGB1 | Fibro 2 | Fibro 6 |
| LAMB1_ITGB1 | LAMB1 | ITGB1 | Fibro 2 | Fibro 6 |
| LGALS1_ITGB1 | LGALS1 | ITGB1 | Fibro 2 | Fibro 6 |
| LUM_ITGB1 | LUM | ITGB1 | Fibro 2 | Fibro 6 |
| S100A1_RYR2 | S100A1 | RYR2 | Fibro 2 | Fibro 6 |
| TIMP1_CD63 | TIMP1 | CD63 | Fibro 2 | Fibro 6 |
| TIMP2_ITGB1 | TIMP2 | ITGB1 | Fibro 2 | Fibro 6 |
| VCAN_ITGB1 | VCAN | ITGB1 | Fibro 2 | Fibro 6 |

### Supplemental Table 8. Fibroblast Cluster 5 Communication with Fibroblast Clusters 2-6

| *Normal* | | | | |
|---|---|---|---|---|
| **Pair_Name** | **Ligand** | **Receptor** | **L_cell** | **R_cell** |
| APP_LRP1 | APP | LRP1 | Fibro 5 | Fibro 2 |
| COL1A1_ITGB1 | COL1A1 | ITGB1 | Fibro 5 | Fibro 2 |
| COL1A2_ITGB1 | COL1A2 | ITGB1 | Fibro 5 | Fibro 2 |
| COL3A1_ITGB1 | COL3A1 | ITGB1 | Fibro 5 | Fibro 2 |
| COL6A1_ITGB1 | COL6A1 | ITGB1 | Fibro 5 | Fibro 2 |
| COL6A2_ITGB1 | COL6A2 | ITGB1 | Fibro 5 | Fibro 2 |
| COL6A3_ITGB1 | COL6A3 | ITGB1 | Fibro 5 | Fibro 2 |
| FBLN1_ITGB1 | FBLN1 | ITGB1 | Fibro 5 | Fibro 2 |
| FBN1_ITGB1 | FBN1 | ITGB1 | Fibro 5 | Fibro 2 |
| FN1_ITGB1 | FN1 | ITGB1 | Fibro 5 | Fibro 2 |
| FN1_SDC2 | FN1 | SDC2 | Fibro 5 | Fibro 2 |
| HLA-A_APLP2 | HLA-A | APLP2 | Fibro 5 | Fibro 2 |
| HSPG2_ITGB1 | HSPG2 | ITGB1 | Fibro 5 | Fibro 2 |
| HSPG2_LRP1 | HSPG2 | LRP1 | Fibro 5 | Fibro 2 |
| LAMA2_ITGB1 | LAMA2 | ITGB1 | Fibro 5 | Fibro 2 |
| LAMC1_ITGB1 | LAMC1 | ITGB1 | Fibro 5 | Fibro 2 |
| LGALS1_ITGB1 | LGALS1 | ITGB1 | Fibro 5 | Fibro 2 |
| LUM_ITGB1 | LUM | ITGB1 | Fibro 5 | Fibro 2 |
| MFGE8_PDGFRB | MFGE8 | PDGFRB | Fibro 5 | Fibro 2 |
| MMP2_SDC2 | MMP2 | SDC2 | Fibro 5 | Fibro 2 |
| PSAP_LRP1 | PSAP | LRP1 | Fibro 5 | Fibro 2 |
| SERPINE2_LRP1 | SERPINE2 | LRP1 | Fibro 5 | Fibro 2 |
| SERPING1_LRP1 | SERPING1 | LRP1 | Fibro 5 | Fibro 2 |
| TFPI_LRP1 | TFPI | LRP1 | Fibro 5 | Fibro 2 |
| TIMP1_CD63 | TIMP1 | CD63 | Fibro 5 | Fibro 2 |
| VCAN_ITGB1 | VCAN | ITGB1 | Fibro 5 | Fibro 2 |
| APP_LRP1 | APP | LRP1 | Fibro 5 | Fibro 3 |
| COL1A1_ITGB1 | COL1A1 | ITGB1 | Fibro 5 | Fibro 3 |
| COL1A2_ITGB1 | COL1A2 | ITGB1 | Fibro 5 | Fibro 3 |
| COL3A1_ITGB1 | COL3A1 | ITGB1 | Fibro 5 | Fibro 3 |
| COL6A1_ITGB1 | COL6A1 | ITGB1 | Fibro 5 | Fibro 3 |
| COL6A2_ITGB1 | COL6A2 | ITGB1 | Fibro 5 | Fibro 3 |
| COL6A3_ITGB1 | COL6A3 | ITGB1 | Fibro 5 | Fibro 3 |
| FBLN1_ITGB1 | FBLN1 | ITGB1 | Fibro 5 | Fibro 3 |
| FBN1_ITGB1 | FBN1 | ITGB1 | Fibro 5 | Fibro 3 |
| FN1_ITGB1 | FN1 | ITGB1 | Fibro 5 | Fibro 3 |
| HSPG2_ITGB1 | HSPG2 | ITGB1 | Fibro 5 | Fibro 3 |
| HSPG2_LRP1 | HSPG2 | LRP1 | Fibro 5 | Fibro 3 |
| LAMA2_ITGB1 | LAMA2 | ITGB1 | Fibro 5 | Fibro 3 |
| LAMC1_ITGB1 | LAMC1 | ITGB1 | Fibro 5 | Fibro 3 |
| LGALS1_ITGB1 | LGALS1 | ITGB1 | Fibro 5 | Fibro 3 |
| LUM_ITGB1 | LUM | ITGB1 | Fibro 5 | Fibro 3 |
| MFGE8_PDGFRB | MFGE8 | PDGFRB | Fibro 5 | Fibro 3 |
| PSAP_LRP1 | PSAP | LRP1 | Fibro 5 | Fibro 3 |
| SERPINE2_LRP1 | SERPINE2 | LRP1 | Fibro 5 | Fibro 3 |
| SERPING1_LRP1 | SERPING1 | LRP1 | Fibro 5 | Fibro 3 |
| TFPI_LRP1 | TFPI | LRP1 | Fibro 5 | Fibro 3 |
| TIMP1_CD63 | TIMP1 | CD63 | Fibro 5 | Fibro 3 |
| VCAN_ITGB1 | VCAN | ITGB1 | Fibro 5 | Fibro 3 |
| APP_LRP1 | APP | LRP1 | Fibro 5 | Fibro 4 |
| COL1A1_ITGB1 | COL1A1 | ITGB1 | Fibro 5 | Fibro 4 |
| COL1A2_ITGB1 | COL1A2 | ITGB1 | Fibro 5 | Fibro 4 |
| COL3A1_ITGB1 | COL3A1 | ITGB1 | Fibro 5 | Fibro 4 |
| COL6A1_ITGB1 | COL6A1 | ITGB1 | Fibro 5 | Fibro 4 |
| COL6A2_ITGB1 | COL6A2 | ITGB1 | Fibro 5 | Fibro 4 |
| COL6A3_ITGB1 | COL6A3 | ITGB1 | Fibro 5 | Fibro 4 |
| FBLN1_ITGB1 | FBLN1 | ITGB1 | Fibro 5 | Fibro 4 |
| FBN1_ITGB1 | FBN1 | ITGB1 | Fibro 5 | Fibro 4 |
| FN1_ITGB1 | FN1 | ITGB1 | Fibro 5 | Fibro 4 |
| HSPG2_ITGB1 | HSPG2 | ITGB1 | Fibro 5 | Fibro 4 |
| HSPG2_LRP1 | HSPG2 | LRP1 | Fibro 5 | Fibro 4 |
| LAMA2_ITGB1 | LAMA2 | ITGB1 | Fibro 5 | Fibro 4 |
| LAMC1_ITGB1 | LAMC1 | ITGB1 | Fibro 5 | Fibro 4 |
| LGALS1_ITGB1 | LGALS1 | ITGB1 | Fibro 5 | Fibro 4 |
| LUM_ITGB1 | LUM | ITGB1 | Fibro 5 | Fibro 4 |
| MFGE8_PDGFRB | MFGE8 | PDGFRB | Fibro 5 | Fibro 4 |
| PSAP_LRP1 | PSAP | LRP1 | Fibro 5 | Fibro 4 |
| SERPINE2_LRP1 | SERPINE2 | LRP1 | Fibro 5 | Fibro 4 |
| SERPING1_LRP1 | SERPING1 | LRP1 | Fibro 5 | Fibro 4 |
| TFPI_LRP1 | TFPI | LRP1 | Fibro 5 | Fibro 4 |
| TIMP1_CD63 | TIMP1 | CD63 | Fibro 5 | Fibro 4 |
| VCAN_ITGB1 | VCAN | ITGB1 | Fibro 5 | Fibro 4 |
| APP_LRP1 | APP | LRP1 | Fibro 5 | Fibro 5 |
| COL1A1_CD36 | COL1A1 | CD36 | Fibro 5 | Fibro 5 |
| COL1A1_ITGB1 | COL1A1 | ITGB1 | Fibro 5 | Fibro 5 |
| COL1A2_CD36 | COL1A2 | CD36 | Fibro 5 | Fibro 5 |
| COL1A2_ITGB1 | COL1A2 | ITGB1 | Fibro 5 | Fibro 5 |
| COL3A1_ITGB1 | COL3A1 | ITGB1 | Fibro 5 | Fibro 5 |
| COL6A1_ITGB1 | COL6A1 | ITGB1 | Fibro 5 | Fibro 5 |
| COL6A2_ITGB1 | COL6A2 | ITGB1 | Fibro 5 | Fibro 5 |
| COL6A3_ITGB1 | COL6A3 | ITGB1 | Fibro 5 | Fibro 5 |
| FBLN1_ITGB1 | FBLN1 | ITGB1 | Fibro 5 | Fibro 5 |
| FBN1_ITGB1 | FBN1 | ITGB1 | Fibro 5 | Fibro 5 |
| FN1_ITGB1 | FN1 | ITGB1 | Fibro 5 | Fibro 5 |
| HSPG2_ITGB1 | HSPG2 | ITGB1 | Fibro 5 | Fibro 5 |
| HSPG2_LRP1 | HSPG2 | LRP1 | Fibro 5 | Fibro 5 |
| LAMA2_ITGB1 | LAMA2 | ITGB1 | Fibro 5 | Fibro 5 |
| LAMC1_ITGB1 | LAMC1 | ITGB1 | Fibro 5 | Fibro 5 |
| LGALS1_ITGB1 | LGALS1 | ITGB1 | Fibro 5 | Fibro 5 |
| LUM_ITGB1 | LUM | ITGB1 | Fibro 5 | Fibro 5 |
| MFGE8_PDGFRB | MFGE8 | PDGFRB | Fibro 5 | Fibro 5 |
| PSAP_LRP1 | PSAP | LRP1 | Fibro 5 | Fibro 5 |
| SERPINE2_LRP1 | SERPINE2 | LRP1 | Fibro 5 | Fibro 5 |
| SERPING1_LRP1 | SERPING1 | LRP1 | Fibro 5 | Fibro 5 |
| TFPI_LRP1 | TFPI | LRP1 | Fibro 5 | Fibro 5 |
| TIMP1_CD63 | TIMP1 | CD63 | Fibro 5 | Fibro 5 |
| VCAN_ITGB1 | VCAN | ITGB1 | Fibro 5 | Fibro 5 |
| APP_LRP1 | APP | LRP1 | Fibro 5 | Fibro 6 |
| COL1A1_ITGB1 | COL1A1 | ITGB1 | Fibro 5 | Fibro 6 |
| COL1A2_ITGB1 | COL1A2 | ITGB1 | Fibro 5 | Fibro 6 |
| COL3A1_ITGB1 | COL3A1 | ITGB1 | Fibro 5 | Fibro 6 |
| COL6A1_ITGB1 | COL6A1 | ITGB1 | Fibro 5 | Fibro 6 |
| COL6A2_ITGB1 | COL6A2 | ITGB1 | Fibro 5 | Fibro 6 |
| COL6A3_ITGB1 | COL6A3 | ITGB1 | Fibro 5 | Fibro 6 |
| FBLN1_ITGB1 | FBLN1 | ITGB1 | Fibro 5 | Fibro 6 |
| FBN1_ITGB1 | FBN1 | ITGB1 | Fibro 5 | Fibro 6 |
| FN1_ITGB1 | FN1 | ITGB1 | Fibro 5 | Fibro 6 |
| HSPG2_ITGB1 | HSPG2 | ITGB1 | Fibro 5 | Fibro 6 |
| HSPG2_LRP1 | HSPG2 | LRP1 | Fibro 5 | Fibro 6 |
| LAMA2_ITGB1 | LAMA2 | ITGB1 | Fibro 5 | Fibro 6 |
| LAMC1_ITGB1 | LAMC1 | ITGB1 | Fibro 5 | Fibro 6 |
| LGALS1_ITGB1 | LGALS1 | ITGB1 | Fibro 5 | Fibro 6 |
| LUM_ITGB1 | LUM | ITGB1 | Fibro 5 | Fibro 6 |
| MFGE8_PDGFRB | MFGE8 | PDGFRB | Fibro 5 | Fibro 6 |
| PSAP_LRP1 | PSAP | LRP1 | Fibro 5 | Fibro 6 |
| SERPINE2_LRP1 | SERPINE2 | LRP1 | Fibro 5 | Fibro 6 |
| SERPING1_LRP1 | SERPING1 | LRP1 | Fibro 5 | Fibro 6 |
| TFPI_LRP1 | TFPI | LRP1 | Fibro 5 | Fibro 6 |
| TIMP1_CD63 | TIMP1 | CD63 | Fibro 5 | Fibro 6 |
| VCAN_ITGB1 | VCAN | ITGB1 | Fibro 5 | Fibro 6 |

| *Non-obstructive* | | | | |
|---|---|---|---|---|
| **Pair_Name** | **Ligand** | **Receptor** | **L_cell** | **R_cell** |
| A2M_LRP1 | A2M | LRP1 | Fibro 5 | Fibro 2 |
| CALM1_RYR2 | CALM1 | RYR2 | Fibro 5 | Fibro 2 |
| FN1_ITGB1 | FN1 | ITGB1 | Fibro 5 | Fibro 2 |
| HSP90AA1_LRP1 | HSP90AA1 | LRP1 | Fibro 5 | Fibro 2 |
| LPL_LRP1 | LPL | LRP1 | Fibro 5 | Fibro 2 |
| LUM_ITGB1 | LUM | ITGB1 | Fibro 5 | Fibro 2 |
| PSAP_LRP1 | PSAP | LRP1 | Fibro 5 | Fibro 2 |
| S100A1_RYR2 | S100A1 | RYR2 | Fibro 5 | Fibro 2 |
| CALM1_RYR2 | CALM1 | RYR2 | Fibro 5 | Fibro 3 |
| FN1_ITGB1 | FN1 | ITGB1 | Fibro 5 | Fibro 3 |
| LUM_ITGB1 | LUM | ITGB1 | Fibro 5 | Fibro 3 |
| S100A1_RYR2 | S100A1 | RYR2 | Fibro 5 | Fibro 3 |
| CALM1_RYR2 | CALM1 | RYR2 | Fibro 5 | Fibro 4 |
| FN1_ITGB1 | FN1 | ITGB1 | Fibro 5 | Fibro 4 |
| LUM_ITGB1 | LUM | ITGB1 | Fibro 5 | Fibro 4 |
| S100A1_RYR2 | S100A1 | RYR2 | Fibro 5 | Fibro 4 |
| CALM1_RYR2 | CALM1 | RYR2 | Fibro 5 | Fibro 5 |
| FN1_ITGB1 | FN1 | ITGB1 | Fibro 5 | Fibro 5 |
| LUM_ITGB1 | LUM | ITGB1 | Fibro 5 | Fibro 5 |
| S100A1_RYR2 | S100A1 | RYR2 | Fibro 5 | Fibro 5 |
| CALM1_RYR2 | CALM1 | RYR2 | Fibro 5 | Fibro 6 |
| FN1_ITGB1 | FN1 | ITGB1 | Fibro 5 | Fibro 6 |
| LUM_ITGB1 | LUM | ITGB1 | Fibro 5 | Fibro 6 |
| S100A1_RYR2 | S100A1 | RYR2 | Fibro 5 | Fibro 6 |

#### Supplemental Table 9. Fibroblast Communication with Fibroblasts and Cardiomyocyte Cluster 4

| ***Normal*** | | | | |
|---|---|---|---|---|
| **Pair_Name** | **Ligand** | **Receptor** | **L_cell** | **R_cell** |
| APP_LRP1 | APP | LRP1 | Fibro | Fibro |
| C3_CD81 | C3 | CD81 | Fibro | Fibro |
| C3_LRP1 | C3 | LRP1 | Fibro | Fibro |
| COL1A1_ITGB1 | COL1A1 | ITGB1 | Fibro | Fibro |
| COL1A2_ITGB1 | COL1A2 | ITGB1 | Fibro | Fibro |
| COL3A1_ITGB1 | COL3A1 | ITGB1 | Fibro | Fibro |
| COL4A1_ITGB1 | COL4A1 | ITGB1 | Fibro | Fibro |
| COL6A1_ITGB1 | COL6A1 | ITGB1 | Fibro | Fibro |
| COL6A2_ITGB1 | COL6A2 | ITGB1 | Fibro | Fibro |
| COL6A3_ITGB1 | COL6A3 | ITGB1 | Fibro | Fibro |
| CTGF_LRP1 | CTGF | LRP1 | Fibro | Fibro |
| FBLN1_ITGB1 | FBLN1 | ITGB1 | Fibro | Fibro |
| FBN1_ITGB1 | FBN1 | ITGB1 | Fibro | Fibro |
| FN1_ITGB1 | FN1 | ITGB1 | Fibro | Fibro |
| HSP90B1_LRP1 | HSP90B1 | LRP1 | Fibro | Fibro |
| HSPG2_ITGB1 | HSPG2 | ITGB1 | Fibro | Fibro |
| HSPG2_LRP1 | HSPG2 | LRP1 | Fibro | Fibro |
| LAMA2_ITGB1 | LAMA2 | ITGB1 | Fibro | Fibro |
| LAMB1_ITGB1 | LAMB1 | ITGB1 | Fibro | Fibro |
| LAMC1_ITGB1 | LAMC1 | ITGB1 | Fibro | Fibro |
| LGALS1_ITGB1 | LGALS1 | ITGB1 | Fibro | Fibro |
| LUM_ITGB1 | LUM | ITGB1 | Fibro | Fibro |
| MFGE8_PDGFRB | MFGE8 | PDGFRB | Fibro | Fibro |
| NID1_ITGB1 | NID1 | ITGB1 | Fibro | Fibro |
| PSAP_LRP1 | PSAP | LRP1 | Fibro | Fibro |
| SERPINE2_LRP1 | SERPINE2 | LRP1 | Fibro | Fibro |
| SERPING1_LRP1 | SERPING1 | LRP1 | Fibro | Fibro |
| TFPI_LRP1 | TFPI | LRP1 | Fibro | Fibro |
| TIMP1_CD63 | TIMP1 | CD63 | Fibro | Fibro |
| VCAN_ITGB1 | VCAN | ITGB1 | Fibro | Fibro |
| APP_CAV1 | APP | CAV1 | Fibro | CM 4 |
| CALM2_CACNA1C | CALM2 | CACNA1C | Fibro | CM 4 |
| CALM2_INSR | CALM2 | INSR | Fibro | CM 4 |
| COL1A1_CD36 | COL1A1 | CD36 | Fibro | CM 4 |
| COL1A1_ITGB1 | COL1A1 | ITGB1 | Fibro | CM 4 |
| COL1A2_CD36 | COL1A2 | CD36 | Fibro | CM 4 |
| COL1A2_ITGB1 | COL1A2 | ITGB1 | Fibro | CM 4 |
| COL3A1_ITGB1 | COL3A1 | ITGB1 | Fibro | CM 4 |
| COL4A1_ITGB1 | COL4A1 | ITGB1 | Fibro | CM 4 |
| COL6A1_ITGB1 | COL6A1 | ITGB1 | Fibro | CM 4 |
| COL6A2_ITGB1 | COL6A2 | ITGB1 | Fibro | CM 4 |
| COL6A3_ITGB1 | COL6A3 | ITGB1 | Fibro | CM 4 |
| FBLN1_ITGB1 | FBLN1 | ITGB1 | Fibro | CM 4 |
| FBN1_ITGB1 | FBN1 | ITGB1 | Fibro | CM 4 |
| FN1_ITGB1 | FN1 | ITGB1 | Fibro | CM 4 |
| HSPG2_ITGB1 | HSPG2 | ITGB1 | Fibro | CM 4 |
| IGF1_INSR | IGF1 | INSR | Fibro | CM 4 |
| LAMA2_ITGB1 | LAMA2 | ITGB1 | Fibro | CM 4 |
| LAMB1_ITGB1 | LAMB1 | ITGB1 | Fibro | CM 4 |
| LAMC1_ITGB1 | LAMC1 | ITGB1 | Fibro | CM 4 |
| LGALS1_ITGB1 | LGALS1 | ITGB1 | Fibro | CM 4 |
| LUM_ITGB1 | LUM | ITGB1 | Fibro | CM 4 |
| NID1_ITGB1 | NID1 | ITGB1 | Fibro | CM 4 |
| TIMP1_CD63 | TIMP1 | CD63 | Fibro | CM 4 |
| VCAN_ITGB1 | VCAN | ITGB1 | Fibro | CM 4 |

| ***Non-obstructive*** | | | | |
|---|---|---|---|---|
| **Pair_Name** | **Ligand** | **Receptor** | **L_cell** | **R_cell** |
| CALM1_RYR2 | CALM1 | RYR2 | Fibro | Fibro |
| COL1A2_CD36 | COL1A2 | CD36 | Fibro | Fibro |
| COL1A2_ITGB1 | COL1A2 | ITGB1 | Fibro | Fibro |
| COL3A1_ITGB1 | COL3A1 | ITGB1 | Fibro | Fibro |
| COL4A1_ITGB1 | COL4A1 | ITGB1 | Fibro | Fibro |
| COL6A1_ITGB1 | COL6A1 | ITGB1 | Fibro | Fibro |
| COL6A2_ITGB1 | COL6A2 | ITGB1 | Fibro | Fibro |
| COL6A3_ITGB1 | COL6A3 | ITGB1 | Fibro | Fibro |
| FN1_ITGB1 | FN1 | ITGB1 | Fibro | Fibro |
| LAMA2_ITGB1 | LAMA2 | ITGB1 | Fibro | Fibro |
| LGALS1_ITGB1 | LGALS1 | ITGB1 | Fibro | Fibro |
| LUM_ITGB1 | LUM | ITGB1 | Fibro | Fibro |
| S100A1_RYR2 | S100A1 | RYR2 | Fibro | Fibro |
| CALM1_RYR2 | CALM1 | RYR2 | Fibro | CM 4 |
| COL1A2_CD36 | COL1A2 | CD36 | Fibro | CM 4 |
| COL1A2_ITGB1 | COL1A2 | ITGB1 | Fibro | CM 4 |
| COL3A1_ITGB1 | COL3A1 | ITGB1 | Fibro | CM 4 |
| COL4A1_ITGB1 | COL4A1 | ITGB1 | Fibro | CM 4 |
| COL6A1_ITGB1 | COL6A1 | ITGB1 | Fibro | CM 4 |
| COL6A2_ITGB1 | COL6A2 | ITGB1 | Fibro | CM 4 |
| COL6A3_ITGB1 | COL6A3 | ITGB1 | Fibro | CM 4 |
| FN1_ITGB1 | FN1 | ITGB1 | Fibro | CM 4 |
| LAMA2_ITGB1 | LAMA2 | ITGB1 | Fibro | CM 4 |
| LGALS1_ITGB1 | LGALS1 | ITGB1 | Fibro | CM 4 |
| LUM_ITGB1 | LUM | ITGB1 | Fibro | CM 4 |
| S100A1_RYR2 | S100A1 | RYR2 | Fibro | CM 4 |

### Supplemental Table 10. Endothelial Cell Communication to Fibroblasts, Cardiomyocyte Cluster 4 and Cardiomyocyte Cluster 8

| *Normal* | | | | |
|---|---|---|---|---|
| **Pair_Name** | **Ligand** | **Receptor** | **L_cell** | **R_cell** |
| A2M_LRP1 | A2M | LRP1 | EC | Fibro |
| APP_LRP1 | APP | LRP1 | EC | Fibro |
| COL1A1_ITGB1 | COL1A1 | ITGB1 | EC | Fibro |
| COL1A2_ITGB1 | COL1A2 | ITGB1 | EC | Fibro |
| COL3A1_ITGB1 | COL3A1 | ITGB1 | EC | Fibro |
| COL6A1_ITGB1 | COL6A1 | ITGB1 | EC | Fibro |
| COL6A2_ITGB1 | COL6A2 | ITGB1 | EC | Fibro |
| FN1_ITGB1 | FN1 | ITGB1 | EC | Fibro |
| GNAS_PTGIR | GNAS | PTGIR | EC | Fibro |
| HSP90AA1_LRP1 | HSP90AA1 | LRP1 | EC | Fibro |
| HSP90B1_LRP1 | HSP90B1 | LRP1 | EC | Fibro |
| HSPG2_ITGB1 | HSPG2 | ITGB1 | EC | Fibro |
| HSPG2_LRP1 | HSPG2 | LRP1 | EC | Fibro |
| LGALS1_ITGB1 | LGALS1 | ITGB1 | EC | Fibro |
| LUM_ITGB1 | LUM | ITGB1 | EC | Fibro |
| MFGE8_PDGFRB | MFGE8 | PDGFRB | EC | Fibro |
| PSAP_LRP1 | PSAP | LRP1 | EC | Fibro |
| TIMP1_CD63 | TIMP1 | CD63 | EC | Fibro |
| VWF_LRP1 | VWF | LRP1 | EC | Fibro |
| APP_CAV1 | APP | CAV1 | EC | CM 4 |
| CALM1_CACNA1C | CALM1 | CACNA1C | EC | CM 4 |
| CALM1_INSR | CALM1 | INSR | EC | CM 4 |
| CALM1_RYR2 | CALM1 | RYR2 | EC | CM 4 |
| CALM2_CACNA1C | CALM2 | CACNA1C | EC | CM 4 |
| CALM2_INSR | CALM2 | INSR | EC | CM 4 |
| COL1A1_CD36 | COL1A1 | CD36 | EC | CM 4 |
| COL1A1_ITGB1 | COL1A1 | ITGB1 | EC | CM 4 |
| COL1A2_CD36 | COL1A2 | CD36 | EC | CM 4 |
| COL1A2_ITGB1 | COL1A2 | ITGB1 | EC | CM 4 |
| COL3A1_ITGB1 | COL3A1 | ITGB1 | EC | CM 4 |
| COL6A1_ITGB1 | COL6A1 | ITGB1 | EC | CM 4 |
| COL6A2_ITGB1 | COL6A2 | ITGB1 | EC | CM 4 |
| FN1_ITGB1 | FN1 | ITGB1 | EC | CM 4 |
| HSPG2_ITGB1 | HSPG2 | ITGB1 | EC | CM 4 |
| LGALS1_ITGB1 | LGALS1 | ITGB1 | EC | CM 4 |
| LUM_ITGB1 | LUM | ITGB1 | EC | CM 4 |
| TIMP1_CD63 | TIMP1 | CD63 | EC | CM 4 |
| CALM1_CACNA1C | CALM1 | CACNA1C | EC | CM 8 |
| CALM1_INSR | CALM1 | INSR | EC | CM 8 |
| CALM1_RYR2 | CALM1 | RYR2 | EC | CM 8 |
| CALM2_CACNA1C | CALM2 | CACNA1C | EC | CM 8 |
| CALM2_INSR | CALM2 | INSR | EC | CM 8 |
| COL1A1_CD36 | COL1A1 | CD36 | EC | CM 8 |
| COL1A1_ITGB1 | COL1A1 | ITGB1 | EC | CM 8 |
| COL1A2_CD36 | COL1A2 | CD36 | EC | CM 8 |
| COL1A2_ITGB1 | COL1A2 | ITGB1 | EC | CM 8 |
| COL3A1_ITGB1 | COL3A1 | ITGB1 | EC | CM 8 |
| COL6A1_ITGB1 | COL6A1 | ITGB1 | EC | CM 8 |
| COL6A2_ITGB1 | COL6A2 | ITGB1 | EC | CM 8 |
| FN1_ITGB1 | FN1 | ITGB1 | EC | CM 8 |
| HSPG2_ITGB1 | HSPG2 | ITGB1 | EC | CM 8 |
| LGALS1_ITGB1 | LGALS1 | ITGB1 | EC | CM 8 |
| LUM_ITGB1 | LUM | ITGB1 | EC | CM 8 |
| TIMP1_CD63 | TIMP1 | CD63 | EC | CM 8 |

| *Non-obstructive* | | | | |
|---|---|---|---|---|
| **Pair_Name** | **Ligand** | **Receptor** | **L_cell** | **R_cell** |
| CALM1_RYR2 | CALM1 | RYR2 | EC | Fibro |
| FN1_ITGB1 | FN1 | ITGB1 | EC | Fibro |
| LGALS1_ITGB1 | LGALS1 | ITGB1 | EC | Fibro |
| S100A1_RYR2 | S100A1 | RYR2 | EC | Fibro |
| CALM1_RYR2 | CALM1 | RYR2 | EC | CM 4 |
| FN1_ITGB1 | FN1 | ITGB1 | EC | CM 4 |
| LGALS1_ITGB1 | LGALS1 | ITGB1 | EC | CM 4 |
| S100A1_RYR2 | S100A1 | RYR2 | EC | CM 4 |
| CALM1_CACNA1C | CALM1 | CACNA1C | EC | CM 8 |
| CALM1_RYR2 | CALM1 | RYR2 | EC | CM 8 |
| FN1_ITGB1 | FN1 | ITGB1 | EC | CM 8 |
| LGALS1_ITGB1 | LGALS1 | ITGB1 | EC | CM 8 |
| S100A1_RYR2 | S100A1 | RYR2 | EC | CM 8 |

### Supplemental Table 11. Cardiomyocyte Cluster 2, 5, Fibroblast and Smooth Muscle Cell Communication to Cardiomyocyte Cluster 9

| *Normal* | | | | |
|---|---|---|---|---|
| **Pair_Name** | **Ligand** | **Receptor** | **L_cell** | **R_cell** |
| COL1A2_CD36 | COL1A2 | CD36 | CM 2 | CM 9 |
| TIMP1_CD63 | TIMP1 | CD63 | CM 2 | CM 9 |
| CALM1_RYR2 | CALM1 | RYR2 | CM 5 | CM 9 |
| COL1A2_CD36 | COL1A2 | CD36 | CM 5 | CM 9 |
| S100A1_RYR2 | S100A1 | RYR2 | CM 5 | CM 9 |
| TIMP1_CD63 | TIMP1 | CD63 | CM 5 | CM 9 |
| COL1A1_CD36 | COL1A1 | CD36 | Fibro | CM 9 |
| COL1A2_CD36 | COL1A2 | CD36 | Fibro | CM 9 |
| TIMP1_CD63 | TIMP1 | CD63 | Fibro | CM 9 |
| COL1A2_CD36 | COL1A2 | CD36 | SMC | CM 9 |
| TIMP1_CD63 | TIMP1 | CD63 | SMC | CM 9 |

| *Non-obstructive* | | | | |
|---|---|---|---|---|
| **Pair_Name** | **Ligand** | **Receptor** | **L_cell** | **R_cell** |
| CALM1_CACNA1C | CALM1 | CACNA1C | CM 2 | CM 9 |
| CALM1_PDE1C | CALM1 | PDE1C | CM 2 | CM 9 |
| CALM1_RYR2 | CALM1 | RYR2 | CM 2 | CM 9 |
| CALM2_CACNA1C | CALM2 | CACNA1C | CM 2 | CM 9 |
| CALM2_PDE1C | CALM2 | PDE1C | CM 2 | CM 9 |
| COL1A2_CD36 | COL1A2 | CD36 | CM 2 | CM 9 |
| COL1A2_ITGB1 | COL1A2 | ITGB1 | CM 2 | CM 9 |
| COL6A2_ITGB1 | COL6A2 | ITGB1 | CM 2 | CM 9 |
| FN1_ITGB1 | FN1 | ITGB1 | CM 2 | CM 9 |
| LGALS1_ITGB1 | LGALS1 | ITGB1 | CM 2 | CM 9 |
| LUM_ITGB1 | LUM | ITGB1 | CM 2 | CM 9 |
| S100A1_RYR2 | S100A1 | RYR2 | CM 2 | CM 9 |
| TIMP1_CD63 | TIMP1 | CD63 | CM 2 | CM 9 |
| CALM1_CACNA1C | CALM1 | CACNA1C | CM 5 | CM 9 |
| CALM1_PDE1C | CALM1 | PDE1C | CM 5 | CM 9 |
| CALM1_RYR2 | CALM1 | RYR2 | CM 5 | CM 9 |
| CALM2_CACNA1C | CALM2 | CACNA1C | CM 5 | CM 9 |
| CALM2_PDE1C | CALM2 | PDE1C | CM 5 | CM 9 |
| CALM3_CACNA1C | CALM3 | CACNA1C | CM 5 | CM 9 |
| CALM3_PDE1C | CALM3 | PDE1C | CM 5 | CM 9 |
| CALM3_RYR2 | CALM3 | RYR2 | CM 5 | CM 9 |
| COL1A2_CD36 | COL1A2 | CD36 | CM 5 | CM 9 |
| COL1A2_ITGB1 | COL1A2 | ITGB1 | CM 5 | CM 9 |
| COL6A1_ITGB1 | COL6A1 | ITGB1 | CM 5 | CM 9 |
| COL6A2_ITGB1 | COL6A2 | ITGB1 | CM 5 | CM 9 |
| FN1_ITGB1 | FN1 | ITGB1 | CM 5 | CM 9 |
| LAMB2_ITGB1 | LAMB2 | ITGB1 | CM 5 | CM 9 |
| LGALS1_ITGB1 | LGALS1 | ITGB1 | CM 5 | CM 9 |
| LGALS3BP_ITGB1 | LGALS3BP | ITGB1 | CM 5 | CM 9 |
| LUM_ITGB1 | LUM | ITGB1 | CM 5 | CM 9 |
| S100A1_RYR2 | S100A1 | RYR2 | CM 5 | CM 9 |
| TGM2_ITGB1 | TGM2 | ITGB1 | CM 5 | CM 9 |
| TIMP1_CD63 | TIMP1 | CD63 | CM 5 | CM 9 |
| CALM1_CACNA1C | CALM1 | CACNA1C | Fibro | CM 9 |
| CALM1_PDE1C | CALM1 | PDE1C | Fibro | CM 9 |
| CALM1_RYR2 | CALM1 | RYR2 | Fibro | CM 9 |
| COL1A2_CD36 | COL1A2 | CD36 | Fibro | CM 9 |
| COL1A2_ITGB1 | COL1A2 | ITGB1 | Fibro | CM 9 |
| COL3A1_ITGB1 | COL3A1 | ITGB1 | Fibro | CM 9 |
| COL4A1_ITGB1 | COL4A1 | ITGB1 | Fibro | CM 9 |
| COL6A1_ITGB1 | COL6A1 | ITGB1 | Fibro | CM 9 |
| COL6A2_ITGB1 | COL6A2 | ITGB1 | Fibro | CM 9 |
| COL6A3_ITGB1 | COL6A3 | ITGB1 | Fibro | CM 9 |
| FN1_ITGB1 | FN1 | ITGB1 | Fibro | CM 9 |
| LAMA2_ITGB1 | LAMA2 | ITGB1 | Fibro | CM 9 |
| LGALS1_ITGB1 | LGALS1 | ITGB1 | Fibro | CM 9 |
| LUM_ITGB1 | LUM | ITGB1 | Fibro | CM 9 |
| S100A1_RYR2 | S100A1 | RYR2 | Fibro | CM 9 |
| CALM1_CACNA1C | CALM1 | CACNA1C | SMC | CM 9 |
| CALM1_PDE1C | CALM1 | PDE1C | SMC | CM 9 |
| CALM1_RYR2 | CALM1 | RYR2 | SMC | CM 9 |
| CALM2_CACNA1C | CALM2 | CACNA1C | SMC | CM 9 |
| CALM2_PDE1C | CALM2 | PDE1C | SMC | CM 9 |
| COL1A2_CD36 | COL1A2 | CD36 | SMC | CM 9 |
| COL1A2_ITGB1 | COL1A2 | ITGB1 | SMC | CM 9 |
| COL4A1_ITGB1 | COL4A1 | ITGB1 | SMC | CM 9 |
| COL6A1_ITGB1 | COL6A1 | ITGB1 | SMC | CM 9 |
| COL6A2_ITGB1 | COL6A2 | ITGB1 | SMC | CM 9 |
| FN1_ITGB1 | FN1 | ITGB1 | SMC | CM 9 |
| LGALS1_ITGB1 | LGALS1 | ITGB1 | SMC | CM 9 |
| S100A1_RYR2 | S100A1 | RYR2 | SMC | CM 9 |

Supplemental Table 12. Cardiomyocyte Cluster 5 Communication to Dendritic Cells

| *Normal* | | | | |
|---|---|---|---|---|
| **Pair_Name** | **Ligand** | **Receptor** | **L_cell** | **R_cell** |
| COL1A2_CD36 | COL1A2 | CD36 | CM 5 | DC |
| LGALS1_PTPRC | LGALS1 | PTPRC | CM 5 | DC |
| TIMP1_CD63 | TIMP1 | CD63 | CM 5 | DC |

| *Non-obstructive* | | | | |
|---|---|---|---|---|
| **Pair_Name** | **Ligand** | **Receptor** | **L_cell** | **R_cell** |
| CALM1_RYR2 | CALM1 | RYR2 | CM 5 | DC |
| CALM3_RYR2 | CALM3 | RYR2 | CM 5 | DC |
| COL1A2_CD36 | COL1A2 | CD36 | CM 5 | DC |
| COL1A2_ITGB1 | COL1A2 | ITGB1 | CM 5 | DC |
| COL6A1_ITGB1 | COL6A1 | ITGB1 | CM 5 | DC |
| COL6A2_ITGB1 | COL6A2 | ITGB1 | CM 5 | DC |
| FN1_ITGB1 | FN1 | ITGB1 | CM 5 | DC |
| LAMB2_ITGB1 | LAMB2 | ITGB1 | CM 5 | DC |
| LGALS1_ITGB1 | LGALS1 | ITGB1 | CM 5 | DC |
| LGALS3BP_ITGB1 | LGALS3BP | ITGB1 | CM 5 | DC |
| LUM_ITGB1 | LUM | ITGB1 | CM 5 | DC |
| MIF_CD74 | MIF | CD74 | CM 5 | DC |
| S100A1_RYR2 | S100A1 | RYR2 | CM 5 | DC |
| TGM2_ITGB1 | TGM2 | ITGB1 | CM 5 | DC |
| TIMP1_CD63 | TIMP1 | CD63 | CM 5 | DC |

### Supplemental Table 13. Fibroblast Cluster 2 and 5 Communication to Fibroblast Clusters 2-6.

| Normal | | | | |
|---|---|---|---|---|
| Pair_Name | Ligand | Receptor | L_cell | R_cell |
| APP_LRP1 | APP | LRP1 | Fibro 2 | Fibro 2 |
| C3_CD81 | C3 | CD81 | Fibro 2 | Fibro 2 |
| C3_LRP1 | C3 | LRP1 | Fibro 2 | Fibro 2 |
| CALR_LRP1 | CALR | LRP1 | Fibro 2 | Fibro 2 |
| CD99_CD81 | CD99 | CD81 | Fibro 2 | Fibro 2 |
| COL18A1_ITGB1 | COL18A1 | ITGB1 | Fibro 2 | Fibro 2 |
| COL1A1_ITGB1 | COL1A1 | ITGB1 | Fibro 2 | Fibro 2 |
| COL1A2_ITGB1 | COL1A2 | ITGB1 | Fibro 2 | Fibro 2 |
| COL3A1_ITGB1 | COL3A1 | ITGB1 | Fibro 2 | Fibro 2 |
| COL4A1_ITGB1 | COL4A1 | ITGB1 | Fibro 2 | Fibro 2 |
| COL5A1_ITGB1 | COL5A1 | ITGB1 | Fibro 2 | Fibro 2 |
| COL5A2_ITGB1 | COL5A2 | ITGB1 | Fibro 2 | Fibro 2 |
| COL6A1_ITGB1 | COL6A1 | ITGB1 | Fibro 2 | Fibro 2 |
| COL6A2_ITGB1 | COL6A2 | ITGB1 | Fibro 2 | Fibro 2 |
| COL6A3_ITGB1 | COL6A3 | ITGB1 | Fibro 2 | Fibro 2 |
| CTGF_LRP1 | CTGF | LRP1 | Fibro 2 | Fibro 2 |
| FBLN1_ITGB1 | FBLN1 | ITGB1 | Fibro 2 | Fibro 2 |
| FBN1_ITGB1 | FBN1 | ITGB1 | Fibro 2 | Fibro 2 |
| FN1_ITGB1 | FN1 | ITGB1 | Fibro 2 | Fibro 2 |
| FN1_SDC2 | FN1 | SDC2 | Fibro 2 | Fibro 2 |
| HLA-A_APLP2 | HLA-A | APLP2 | Fibro 2 | Fibro 2 |
| HSP90AA1_LRP1 | HSP90AA1 | LRP1 | Fibro 2 | Fibro 2 |
| HSP90B1_LRP1 | HSP90B1 | LRP1 | Fibro 2 | Fibro 2 |
| HSPG2_ITGB1 | HSPG2 | ITGB1 | Fibro 2 | Fibro 2 |
| HSPG2_LRP1 | HSPG2 | LRP1 | Fibro 2 | Fibro 2 |
| LAMA2_ITGB1 | LAMA2 | ITGB1 | Fibro 2 | Fibro 2 |
| LAMB1_ITGB1 | LAMB1 | ITGB1 | Fibro 2 | Fibro 2 |
| LAMC1_ITGB1 | LAMC1 | ITGB1 | Fibro 2 | Fibro 2 |
| LGALS1_ITGB1 | LGALS1 | ITGB1 | Fibro 2 | Fibro 2 |
| LGALS3BP_ITGB1 | LGALS3BP | ITGB1 | Fibro 2 | Fibro 2 |
| LRPAP1_LRP1 | LRPAP1 | LRP1 | Fibro 2 | Fibro 2 |
| LUM_ITGB1 | LUM | ITGB1 | Fibro 2 | Fibro 2 |
| MFGE8_PDGFRB | MFGE8 | PDGFRB | Fibro 2 | Fibro 2 |
| MMP2_SDC2 | MMP2 | SDC2 | Fibro 2 | Fibro 2 |
| NID1_ITGB1 | NID1 | ITGB1 | Fibro 2 | Fibro 2 |
| PSAP_LRP1 | PSAP | LRP1 | Fibro 2 | Fibro 2 |
| SERPINE2_LRP1 | SERPINE2 | LRP1 | Fibro 2 | Fibro 2 |
| SERPING1_LRP1 | SERPING1 | LRP1 | Fibro 2 | Fibro 2 |
| TFPI_LRP1 | TFPI | LRP1 | Fibro 2 | Fibro 2 |
| THBS2_ITGB1 | THBS2 | ITGB1 | Fibro 2 | Fibro 2 |
| TIMP1_CD63 | TIMP1 | CD63 | Fibro 2 | Fibro 2 |
| TIMP2_ITGB1 | TIMP2 | ITGB1 | Fibro 2 | Fibro 2 |
| VCAN_ITGB1 | VCAN | ITGB1 | Fibro 2 | Fibro 2 |
| APP_LRP1 | APP | LRP1 | Fibro 2 | Fibro 3 |
| C3_CD81 | C3 | CD81 | Fibro 2 | Fibro 3 |
| C3_LRP1 | C3 | LRP1 | Fibro 2 | Fibro 3 |
| CALR_LRP1 | CALR | LRP1 | Fibro 2 | Fibro 3 |
| CD99_CD81 | CD99 | CD81 | Fibro 2 | Fibro 3 |
| COL18A1_ITGB1 | COL18A1 | ITGB1 | Fibro 2 | Fibro 3 |
| COL1A1_ITGB1 | COL1A1 | ITGB1 | Fibro 2 | Fibro 3 |
| COL1A2_ITGB1 | COL1A2 | ITGB1 | Fibro 2 | Fibro 3 |
| COL3A1_ITGB1 | COL3A1 | ITGB1 | Fibro 2 | Fibro 3 |
| COL4A1_ITGB1 | COL4A1 | ITGB1 | Fibro 2 | Fibro 3 |
| COL5A1_ITGB1 | COL5A1 | ITGB1 | Fibro 2 | Fibro 3 |
| COL5A2_ITGB1 | COL5A2 | ITGB1 | Fibro 2 | Fibro 3 |
| COL6A1_ITGB1 | COL6A1 | ITGB1 | Fibro 2 | Fibro 3 |
| COL6A2_ITGB1 | COL6A2 | ITGB1 | Fibro 2 | Fibro 3 |
| COL6A3_ITGB1 | COL6A3 | ITGB1 | Fibro 2 | Fibro 3 |
| CTGF_LRP1 | CTGF | LRP1 | Fibro 2 | Fibro 3 |
| FBLN1_ITGB1 | FBLN1 | ITGB1 | Fibro 2 | Fibro 3 |
| FBN1_ITGB1 | FBN1 | ITGB1 | Fibro 2 | Fibro 3 |
| FN1_ITGB1 | FN1 | ITGB1 | Fibro 2 | Fibro 3 |
| HSP90AA1_LRP1 | HSP90AA1 | LRP1 | Fibro 2 | Fibro 3 |
| HSP90B1_LRP1 | HSP90B1 | LRP1 | Fibro 2 | Fibro 3 |
| HSPG2_ITGB1 | HSPG2 | ITGB1 | Fibro 2 | Fibro 3 |
| HSPG2_LRP1 | HSPG2 | LRP1 | Fibro 2 | Fibro 3 |
| LAMA2_ITGB1 | LAMA2 | ITGB1 | Fibro 2 | Fibro 3 |
| LAMB1_ITGB1 | LAMB1 | ITGB1 | Fibro 2 | Fibro 3 |
| LAMC1_ITGB1 | LAMC1 | ITGB1 | Fibro 2 | Fibro 3 |
| LGALS1_ITGB1 | LGALS1 | ITGB1 | Fibro 2 | Fibro 3 |
| LGALS3BP_ITGB1 | LGALS3BP | ITGB1 | Fibro 2 | Fibro 3 |
| LRPAP1_LRP1 | LRPAP1 | LRP1 | Fibro 2 | Fibro 3 |
| LUM_ITGB1 | LUM | ITGB1 | Fibro 2 | Fibro 3 |
| MFGE8_PDGFRB | MFGE8 | PDGFRB | Fibro 2 | Fibro 3 |
| NID1_ITGB1 | NID1 | ITGB1 | Fibro 2 | Fibro 3 |
| PSAP_LRP1 | PSAP | LRP1 | Fibro 2 | Fibro 3 |
| SERPINE2_LRP1 | SERPINE2 | LRP1 | Fibro 2 | Fibro 3 |
| SERPING1_LRP1 | SERPING1 | LRP1 | Fibro 2 | Fibro 3 |
| TFPI_LRP1 | TFPI | LRP1 | Fibro 2 | Fibro 3 |
| THBS2_ITGB1 | THBS2 | ITGB1 | Fibro 2 | Fibro 3 |
| TIMP1_CD63 | TIMP1 | CD63 | Fibro 2 | Fibro 3 |
| TIMP2_ITGB1 | TIMP2 | ITGB1 | Fibro 2 | Fibro 3 |
| VCAN_ITGB1 | VCAN | ITGB1 | Fibro 2 | Fibro 3 |
| APP_LRP1 | APP | LRP1 | Fibro 2 | Fibro 4 |
| C3_CD81 | C3 | CD81 | Fibro 2 | Fibro 4 |
| C3_LRP1 | C3 | LRP1 | Fibro 2 | Fibro 4 |
| CALR_LRP1 | CALR | LRP1 | Fibro 2 | Fibro 4 |
| CD99_CD81 | CD99 | CD81 | Fibro 2 | Fibro 4 |
| COL18A1_ITGB1 | COL18A1 | ITGB1 | Fibro 2 | Fibro 4 |
| COL1A1_ITGB1 | COL1A1 | ITGB1 | Fibro 2 | Fibro 4 |
| COL1A2_ITGB1 | COL1A2 | ITGB1 | Fibro 2 | Fibro 4 |
| COL3A1_ITGB1 | COL3A1 | ITGB1 | Fibro 2 | Fibro 4 |
| COL4A1_ITGB1 | COL4A1 | ITGB1 | Fibro 2 | Fibro 4 |
| COL5A1_ITGB1 | COL5A1 | ITGB1 | Fibro 2 | Fibro 4 |
| COL5A2_ITGB1 | COL5A2 | ITGB1 | Fibro 2 | Fibro 4 |
| COL6A1_ITGB1 | COL6A1 | ITGB1 | Fibro 2 | Fibro 4 |
| COL6A2_ITGB1 | COL6A2 | ITGB1 | Fibro 2 | Fibro 4 |
| COL6A3_ITGB1 | COL6A3 | ITGB1 | Fibro 2 | Fibro 4 |
| CTGF_LRP1 | CTGF | LRP1 | Fibro 2 | Fibro 4 |
| FBLN1_ITGB1 | FBLN1 | ITGB1 | Fibro 2 | Fibro 4 |
| FBN1_ITGB1 | FBN1 | ITGB1 | Fibro 2 | Fibro 4 |
| FN1_ITGB1 | FN1 | ITGB1 | Fibro 2 | Fibro 4 |
| HSP90AA1_LRP1 | HSP90AA1 | LRP1 | Fibro 2 | Fibro 4 |
| HSP90B1_LRP1 | HSP90B1 | LRP1 | Fibro 2 | Fibro 4 |
| HSPG2_ITGB1 | HSPG2 | ITGB1 | Fibro 2 | Fibro 4 |
| HSPG2_LRP1 | HSPG2 | LRP1 | Fibro 2 | Fibro 4 |
| LAMA2_ITGB1 | LAMA2 | ITGB1 | Fibro 2 | Fibro 4 |
| LAMB1_ITGB1 | LAMB1 | ITGB1 | Fibro 2 | Fibro 4 |
| LAMC1_ITGB1 | LAMC1 | ITGB1 | Fibro 2 | Fibro 4 |
| LGALS1_ITGB1 | LGALS1 | ITGB1 | Fibro 2 | Fibro 4 |
| LGALS3BP_ITGB1 | LGALS3BP | ITGB1 | Fibro 2 | Fibro 4 |
| LRPAP1_LRP1 | LRPAP1 | LRP1 | Fibro 2 | Fibro 4 |
| LUM_ITGB1 | LUM | ITGB1 | Fibro 2 | Fibro 4 |
| MFGE8_PDGFRB | MFGE8 | PDGFRB | Fibro 2 | Fibro 4 |
| NID1_ITGB1 | NID1 | ITGB1 | Fibro 2 | Fibro 4 |
| PSAP_LRP1 | PSAP | LRP1 | Fibro 2 | Fibro 4 |
| SERPINE2_LRP1 | SERPINE2 | LRP1 | Fibro 2 | Fibro 4 |
| SERPING1_LRP1 | SERPING1 | LRP1 | Fibro 2 | Fibro 4 |
| TFPI_LRP1 | TFPI | LRP1 | Fibro 2 | Fibro 4 |
| THBS2_ITGB1 | THBS2 | ITGB1 | Fibro 2 | Fibro 4 |
| TIMP1_CD63 | TIMP1 | CD63 | Fibro 2 | Fibro 4 |
| TIMP2_ITGB1 | TIMP2 | ITGB1 | Fibro 2 | Fibro 4 |
| VCAN_ITGB1 | VCAN | ITGB1 | Fibro 2 | Fibro 4 |
| APP_LRP1 | APP | LRP1 | Fibro 2 | Fibro 5 |
| C3_CD81 | C3 | CD81 | Fibro 2 | Fibro 5 |
| C3_LRP1 | C3 | LRP1 | Fibro 2 | Fibro 5 |
| CALR_LRP1 | CALR | LRP1 | Fibro 2 | Fibro 5 |
| CD99_CD81 | CD99 | CD81 | Fibro 2 | Fibro 5 |
| COL18A1_ITGB1 | COL18A1 | ITGB1 | Fibro 2 | Fibro 5 |
| COL1A1_CD36 | COL1A1 | CD36 | Fibro 2 | Fibro 5 |
| COL1A1_ITGB1 | COL1A1 | ITGB1 | Fibro 2 | Fibro 5 |
| COL1A2_CD36 | COL1A2 | CD36 | Fibro 2 | Fibro 5 |
| COL1A2_ITGB1 | COL1A2 | ITGB1 | Fibro 2 | Fibro 5 |
| COL3A1_ITGB1 | COL3A1 | ITGB1 | Fibro 2 | Fibro 5 |
| COL4A1_ITGB1 | COL4A1 | ITGB1 | Fibro 2 | Fibro 5 |
| COL5A1_ITGB1 | COL5A1 | ITGB1 | Fibro 2 | Fibro 5 |
| COL5A2_ITGB1 | COL5A2 | ITGB1 | Fibro 2 | Fibro 5 |
| COL6A1_ITGB1 | COL6A1 | ITGB1 | Fibro 2 | Fibro 5 |
| COL6A2_ITGB1 | COL6A2 | ITGB1 | Fibro 2 | Fibro 5 |
| COL6A3_ITGB1 | COL6A3 | ITGB1 | Fibro 2 | Fibro 5 |
| CTGF_LRP1 | CTGF | LRP1 | Fibro 2 | Fibro 5 |
| FBLN1_ITGB1 | FBLN1 | ITGB1 | Fibro 2 | Fibro 5 |
| FBN1_ITGB1 | FBN1 | ITGB1 | Fibro 2 | Fibro 5 |
| FN1_ITGB1 | FN1 | ITGB1 | Fibro 2 | Fibro 5 |
| HSP90AA1_LRP1 | HSP90AA1 | LRP1 | Fibro 2 | Fibro 5 |
| HSP90B1_LRP1 | HSP90B1 | LRP1 | Fibro 2 | Fibro 5 |
| HSPG2_ITGB1 | HSPG2 | ITGB1 | Fibro 2 | Fibro 5 |
| HSPG2_LRP1 | HSPG2 | LRP1 | Fibro 2 | Fibro 5 |
| LAMA2_ITGB1 | LAMA2 | ITGB1 | Fibro 2 | Fibro 5 |
| LAMB1_ITGB1 | LAMB1 | ITGB1 | Fibro 2 | Fibro 5 |
| LAMC1_ITGB1 | LAMC1 | ITGB1 | Fibro 2 | Fibro 5 |
| LGALS1_ITGB1 | LGALS1 | ITGB1 | Fibro 2 | Fibro 5 |
| LGALS3BP_ITGB1 | LGALS3BP | ITGB1 | Fibro 2 | Fibro 5 |
| LRPAP1_LRP1 | LRPAP1 | LRP1 | Fibro 2 | Fibro 5 |
| LUM_ITGB1 | LUM | ITGB1 | Fibro 2 | Fibro 5 |
| MFGE8_PDGFRB | MFGE8 | PDGFRB | Fibro 2 | Fibro 5 |
| NID1_ITGB1 | NID1 | ITGB1 | Fibro 2 | Fibro 5 |
| PSAP_LRP1 | PSAP | LRP1 | Fibro 2 | Fibro 5 |
| SERPINE2_LRP1 | SERPINE2 | LRP1 | Fibro 2 | Fibro 5 |
| SERPING1_LRP1 | SERPING1 | LRP1 | Fibro 2 | Fibro 5 |
| TFPI_LRP1 | TFPI | LRP1 | Fibro 2 | Fibro 5 |
| THBS2_CD36 | THBS2 | CD36 | Fibro 2 | Fibro 5 |
| THBS2_ITGB1 | THBS2 | ITGB1 | Fibro 2 | Fibro 5 |
| TIMP1_CD63 | TIMP1 | CD63 | Fibro 2 | Fibro 5 |
| TIMP2_ITGB1 | TIMP2 | ITGB1 | Fibro 2 | Fibro 5 |
| VCAN_ITGB1 | VCAN | ITGB1 | Fibro 2 | Fibro 5 |
| APP_LRP1 | APP | LRP1 | Fibro 2 | Fibro 6 |
| C3_CD81 | C3 | CD81 | Fibro 2 | Fibro 6 |
| C3_LRP1 | C3 | LRP1 | Fibro 2 | Fibro 6 |
| CALR_LRP1 | CALR | LRP1 | Fibro 2 | Fibro 6 |
| CD99_CD81 | CD99 | CD81 | Fibro 2 | Fibro 6 |
| COL18A1_ITGB1 | COL18A1 | ITGB1 | Fibro 2 | Fibro 6 |
| COL1A1_ITGB1 | COL1A1 | ITGB1 | Fibro 2 | Fibro 6 |
| COL1A2_ITGB1 | COL1A2 | ITGB1 | Fibro 2 | Fibro 6 |
| COL3A1_ITGB1 | COL3A1 | ITGB1 | Fibro 2 | Fibro 6 |
| COL4A1_ITGB1 | COL4A1 | ITGB1 | Fibro 2 | Fibro 6 |
| COL5A1_ITGB1 | COL5A1 | ITGB1 | Fibro 2 | Fibro 6 |
| COL5A2_ITGB1 | COL5A2 | ITGB1 | Fibro 2 | Fibro 6 |
| COL6A1_ITGB1 | COL6A1 | ITGB1 | Fibro 2 | Fibro 6 |
| COL6A2_ITGB1 | COL6A2 | ITGB1 | Fibro 2 | Fibro 6 |
| COL6A3_ITGB1 | COL6A3 | ITGB1 | Fibro 2 | Fibro 6 |
| CTGF_LRP1 | CTGF | LRP1 | Fibro 2 | Fibro 6 |
| FBLN1_ITGB1 | FBLN1 | ITGB1 | Fibro 2 | Fibro 6 |
| FBN1_ITGB1 | FBN1 | ITGB1 | Fibro 2 | Fibro 6 |
| FN1_ITGB1 | FN1 | ITGB1 | Fibro 2 | Fibro 6 |
| HSP90AA1_LRP1 | HSP90AA1 | LRP1 | Fibro 2 | Fibro 6 |
| HSP90B1_LRP1 | HSP90B1 | LRP1 | Fibro 2 | Fibro 6 |
| HSPG2_ITGB1 | HSPG2 | ITGB1 | Fibro 2 | Fibro 6 |
| HSPG2_LRP1 | HSPG2 | LRP1 | Fibro 2 | Fibro 6 |
| LAMA2_ITGB1 | LAMA2 | ITGB1 | Fibro 2 | Fibro 6 |
| LAMB1_ITGB1 | LAMB1 | ITGB1 | Fibro 2 | Fibro 6 |
| LAMC1_ITGB1 | LAMC1 | ITGB1 | Fibro 2 | Fibro 6 |
| LGALS1_ITGB1 | LGALS1 | ITGB1 | Fibro 2 | Fibro 6 |
| LGALS3BP_ITGB1 | LGALS3BP | ITGB1 | Fibro 2 | Fibro 6 |
| LRPAP1_LRP1 | LRPAP1 | LRP1 | Fibro 2 | Fibro 6 |
| LUM_ITGB1 | LUM | ITGB1 | Fibro 2 | Fibro 6 |
| MFGE8_PDGFRB | MFGE8 | PDGFRB | Fibro 2 | Fibro 6 |
| NID1_ITGB1 | NID1 | ITGB1 | Fibro 2 | Fibro 6 |
| PSAP_LRP1 | PSAP | LRP1 | Fibro 2 | Fibro 6 |
| SERPINE2_LRP1 | SERPINE2 | LRP1 | Fibro 2 | Fibro 6 |
| SERPING1_LRP1 | SERPING1 | LRP1 | Fibro 2 | Fibro 6 |
| TFPI_LRP1 | TFPI | LRP1 | Fibro 2 | Fibro 6 |
| THBS2_ITGB1 | THBS2 | ITGB1 | Fibro 2 | Fibro 6 |
| TIMP1_CD63 | TIMP1 | CD63 | Fibro 2 | Fibro 6 |
| TIMP2_ITGB1 | TIMP2 | ITGB1 | Fibro 2 | Fibro 6 |
| VCAN_ITGB1 | VCAN | ITGB1 | Fibro 2 | Fibro 6 |
| APP_LRP1 | APP | LRP1 | Fibro 5 | Fibro 2 |
| COL1A1_ITGB1 | COL1A1 | ITGB1 | Fibro 5 | Fibro 2 |
| COL1A2_ITGB1 | COL1A2 | ITGB1 | Fibro 5 | Fibro 2 |
| COL3A1_ITGB1 | COL3A1 | ITGB1 | Fibro 5 | Fibro 2 |
| COL6A1_ITGB1 | COL6A1 | ITGB1 | Fibro 5 | Fibro 2 |
| COL6A2_ITGB1 | COL6A2 | ITGB1 | Fibro 5 | Fibro 2 |
| COL6A3_ITGB1 | COL6A3 | ITGB1 | Fibro 5 | Fibro 2 |
| FBLN1_ITGB1 | FBLN1 | ITGB1 | Fibro 5 | Fibro 2 |
| FBN1_ITGB1 | FBN1 | ITGB1 | Fibro 5 | Fibro 2 |
| FN1_ITGB1 | FN1 | ITGB1 | Fibro 5 | Fibro 2 |
| FN1_SDC2 | FN1 | SDC2 | Fibro 5 | Fibro 2 |
| HLA-A_APLP2 | HLA-A | APLP2 | Fibro 5 | Fibro 2 |
| HSPG2_ITGB1 | HSPG2 | ITGB1 | Fibro 5 | Fibro 2 |
| HSPG2_LRP1 | HSPG2 | LRP1 | Fibro 5 | Fibro 2 |
| LAMA2_ITGB1 | LAMA2 | ITGB1 | Fibro 5 | Fibro 2 |
| LAMC1_ITGB1 | LAMC1 | ITGB1 | Fibro 5 | Fibro 2 |
| LGALS1_ITGB1 | LGALS1 | ITGB1 | Fibro 5 | Fibro 2 |
| LUM_ITGB1 | LUM | ITGB1 | Fibro 5 | Fibro 2 |
| MFGE8_PDGFRB | MFGE8 | PDGFRB | Fibro 5 | Fibro 2 |
| MMP2_SDC2 | MMP2 | SDC2 | Fibro 5 | Fibro 2 |
| PSAP_LRP1 | PSAP | LRP1 | Fibro 5 | Fibro 2 |
| SERPINE2_LRP1 | SERPINE2 | LRP1 | Fibro 5 | Fibro 2 |
| SERPING1_LRP1 | SERPING1 | LRP1 | Fibro 5 | Fibro 2 |
| TFPI_LRP1 | TFPI | LRP1 | Fibro 5 | Fibro 2 |
| TIMP1_CD63 | TIMP1 | CD63 | Fibro 5 | Fibro 2 |
| VCAN_ITGB1 | VCAN | ITGB1 | Fibro 5 | Fibro 2 |
| APP_LRP1 | APP | LRP1 | Fibro 5 | Fibro 3 |
| COL1A1_ITGB1 | COL1A1 | ITGB1 | Fibro 5 | Fibro 3 |
| COL1A2_ITGB1 | COL1A2 | ITGB1 | Fibro 5 | Fibro 3 |
| COL3A1_ITGB1 | COL3A1 | ITGB1 | Fibro 5 | Fibro 3 |
| COL6A1_ITGB1 | COL6A1 | ITGB1 | Fibro 5 | Fibro 3 |
| COL6A2_ITGB1 | COL6A2 | ITGB1 | Fibro 5 | Fibro 3 |
| COL6A3_ITGB1 | COL6A3 | ITGB1 | Fibro 5 | Fibro 3 |
| FBLN1_ITGB1 | FBLN1 | ITGB1 | Fibro 5 | Fibro 3 |
| FBN1_ITGB1 | FBN1 | ITGB1 | Fibro 5 | Fibro 3 |
| FN1_ITGB1 | FN1 | ITGB1 | Fibro 5 | Fibro 3 |
| HSPG2_ITGB1 | HSPG2 | ITGB1 | Fibro 5 | Fibro 3 |
| HSPG2_LRP1 | HSPG2 | LRP1 | Fibro 5 | Fibro 3 |
| LAMA2_ITGB1 | LAMA2 | ITGB1 | Fibro 5 | Fibro 3 |
| LAMC1_ITGB1 | LAMC1 | ITGB1 | Fibro 5 | Fibro 3 |
| LGALS1_ITGB1 | LGALS1 | ITGB1 | Fibro 5 | Fibro 3 |
| LUM_ITGB1 | LUM | ITGB1 | Fibro 5 | Fibro 3 |
| MFGE8_PDGFRB | MFGE8 | PDGFRB | Fibro 5 | Fibro 3 |
| PSAP_LRP1 | PSAP | LRP1 | Fibro 5 | Fibro 3 |
| SERPINE2_LRP1 | SERPINE2 | LRP1 | Fibro 5 | Fibro 3 |
| SERPING1_LRP1 | SERPING1 | LRP1 | Fibro 5 | Fibro 3 |
| TFPI_LRP1 | TFPI | LRP1 | Fibro 5 | Fibro 3 |
| TIMP1_CD63 | TIMP1 | CD63 | Fibro 5 | Fibro 3 |
| VCAN_ITGB1 | VCAN | ITGB1 | Fibro 5 | Fibro 3 |
| APP_LRP1 | APP | LRP1 | Fibro 5 | Fibro 4 |
| COL1A1_ITGB1 | COL1A1 | ITGB1 | Fibro 5 | Fibro 4 |
| COL1A2_ITGB1 | COL1A2 | ITGB1 | Fibro 5 | Fibro 4 |
| COL3A1_ITGB1 | COL3A1 | ITGB1 | Fibro 5 | Fibro 4 |
| COL6A1_ITGB1 | COL6A1 | ITGB1 | Fibro 5 | Fibro 4 |
| COL6A2_ITGB1 | COL6A2 | ITGB1 | Fibro 5 | Fibro 4 |
| COL6A3_ITGB1 | COL6A3 | ITGB1 | Fibro 5 | Fibro 4 |
| FBLN1_ITGB1 | FBLN1 | ITGB1 | Fibro 5 | Fibro 4 |
| FBN1_ITGB1 | FBN1 | ITGB1 | Fibro 5 | Fibro 4 |
| FN1_ITGB1 | FN1 | ITGB1 | Fibro 5 | Fibro 4 |
| HSPG2_ITGB1 | HSPG2 | ITGB1 | Fibro 5 | Fibro 4 |
| HSPG2_LRP1 | HSPG2 | LRP1 | Fibro 5 | Fibro 4 |
| LAMA2_ITGB1 | LAMA2 | ITGB1 | Fibro 5 | Fibro 4 |
| LAMC1_ITGB1 | LAMC1 | ITGB1 | Fibro 5 | Fibro 4 |
| LGALS1_ITGB1 | LGALS1 | ITGB1 | Fibro 5 | Fibro 4 |
| LUM_ITGB1 | LUM | ITGB1 | Fibro 5 | Fibro 4 |
| MFGE8_PDGFRB | MFGE8 | PDGFRB | Fibro 5 | Fibro 4 |
| PSAP_LRP1 | PSAP | LRP1 | Fibro 5 | Fibro 4 |
| SERPINE2_LRP1 | SERPINE2 | LRP1 | Fibro 5 | Fibro 4 |
| SERPING1_LRP1 | SERPING1 | LRP1 | Fibro 5 | Fibro 4 |
| TFPI_LRP1 | TFPI | LRP1 | Fibro 5 | Fibro 4 |
| TIMP1_CD63 | TIMP1 | CD63 | Fibro 5 | Fibro 4 |
| VCAN_ITGB1 | VCAN | ITGB1 | Fibro 5 | Fibro 4 |
| APP_LRP1 | APP | LRP1 | Fibro 5 | Fibro 5 |
| COL1A1_CD36 | COL1A1 | CD36 | Fibro 5 | Fibro 5 |
| COL1A1_ITGB1 | COL1A1 | ITGB1 | Fibro 5 | Fibro 5 |
| COL1A2_CD36 | COL1A2 | CD36 | Fibro 5 | Fibro 5 |
| COL1A2_ITGB1 | COL1A2 | ITGB1 | Fibro 5 | Fibro 5 |
| COL3A1_ITGB1 | COL3A1 | ITGB1 | Fibro 5 | Fibro 5 |
| COL6A1_ITGB1 | COL6A1 | ITGB1 | Fibro 5 | Fibro 5 |
| COL6A2_ITGB1 | COL6A2 | ITGB1 | Fibro 5 | Fibro 5 |
| COL6A3_ITGB1 | COL6A3 | ITGB1 | Fibro 5 | Fibro 5 |
| FBLN1_ITGB1 | FBLN1 | ITGB1 | Fibro 5 | Fibro 5 |
| FBN1_ITGB1 | FBN1 | ITGB1 | Fibro 5 | Fibro 5 |
| FN1_ITGB1 | FN1 | ITGB1 | Fibro 5 | Fibro 5 |
| HSPG2_ITGB1 | HSPG2 | ITGB1 | Fibro 5 | Fibro 5 |
| HSPG2_LRP1 | HSPG2 | LRP1 | Fibro 5 | Fibro 5 |
| LAMA2_ITGB1 | LAMA2 | ITGB1 | Fibro 5 | Fibro 5 |
| LAMC1_ITGB1 | LAMC1 | ITGB1 | Fibro 5 | Fibro 5 |
| LGALS1_ITGB1 | LGALS1 | ITGB1 | Fibro 5 | Fibro 5 |
| LUM_ITGB1 | LUM | ITGB1 | Fibro 5 | Fibro 5 |
| MFGE8_PDGFRB | MFGE8 | PDGFRB | Fibro 5 | Fibro 5 |
| PSAP_LRP1 | PSAP | LRP1 | Fibro 5 | Fibro 5 |
| SERPINE2_LRP1 | SERPINE2 | LRP1 | Fibro 5 | Fibro 5 |
| SERPING1_LRP1 | SERPING1 | LRP1 | Fibro 5 | Fibro 5 |
| TFPI_LRP1 | TFPI | LRP1 | Fibro 5 | Fibro 5 |
| TIMP1_CD63 | TIMP1 | CD63 | Fibro 5 | Fibro 5 |
| VCAN_ITGB1 | VCAN | ITGB1 | Fibro 5 | Fibro 5 |
| APP_LRP1 | APP | LRP1 | Fibro 5 | Fibro 6 |
| COL1A1_ITGB1 | COL1A1 | ITGB1 | Fibro 5 | Fibro 6 |
| COL1A2_ITGB1 | COL1A2 | ITGB1 | Fibro 5 | Fibro 6 |
| COL3A1_ITGB1 | COL3A1 | ITGB1 | Fibro 5 | Fibro 6 |
| COL6A1_ITGB1 | COL6A1 | ITGB1 | Fibro 5 | Fibro 6 |
| COL6A2_ITGB1 | COL6A2 | ITGB1 | Fibro 5 | Fibro 6 |
| COL6A3_ITGB1 | COL6A3 | ITGB1 | Fibro 5 | Fibro 6 |
| FBLN1_ITGB1 | FBLN1 | ITGB1 | Fibro 5 | Fibro 6 |
| FBN1_ITGB1 | FBN1 | ITGB1 | Fibro 5 | Fibro 6 |
| FN1_ITGB1 | FN1 | ITGB1 | Fibro 5 | Fibro 6 |
| HSPG2_ITGB1 | HSPG2 | ITGB1 | Fibro 5 | Fibro 6 |
| HSPG2_LRP1 | HSPG2 | LRP1 | Fibro 5 | Fibro 6 |
| LAMA2_ITGB1 | LAMA2 | ITGB1 | Fibro 5 | Fibro 6 |
| LAMC1_ITGB1 | LAMC1 | ITGB1 | Fibro 5 | Fibro 6 |
| LGALS1_ITGB1 | LGALS1 | ITGB1 | Fibro 5 | Fibro 6 |
| LUM_ITGB1 | LUM | ITGB1 | Fibro 5 | Fibro 6 |
| MFGE8_PDGFRB | MFGE8 | PDGFRB | Fibro 5 | Fibro 6 |
| PSAP_LRP1 | PSAP | LRP1 | Fibro 5 | Fibro 6 |
| SERPINE2_LRP1 | SERPINE2 | LRP1 | Fibro 5 | Fibro 6 |
| SERPING1_LRP1 | SERPING1 | LRP1 | Fibro 5 | Fibro 6 |
| TFPI_LRP1 | TFPI | LRP1 | Fibro 5 | Fibro 6 |
| TIMP1_CD63 | TIMP1 | CD63 | Fibro 5 | Fibro 6 |
| VCAN_ITGB1 | VCAN | ITGB1 | Fibro 5 | Fibro 6 |

| Non-obstructive | | | | |
|---|---|---|---|---|
| Pair_Name | Ligand | Receptor | L_cell | R_cell |
| A2M_LRP1 | A2M | LRP1 | Fibro 2 | Fibro 2 |
| CALM1_RYR2 | CALM1 | RYR2 | Fibro 2 | Fibro 2 |
| CCN2_LRP1 | CCN2 | LRP1 | Fibro 2 | Fibro 2 |
| COL1A1_CD36 | COL1A1 | CD36 | Fibro 2 | Fibro 2 |
| COL1A1_ITGB1 | COL1A1 | ITGB1 | Fibro 2 | Fibro 2 |
| COL1A2_CD36 | COL1A2 | CD36 | Fibro 2 | Fibro 2 |
| COL1A2_ITGB1 | COL1A2 | ITGB1 | Fibro 2 | Fibro 2 |
| COL3A1_ITGB1 | COL3A1 | ITGB1 | Fibro 2 | Fibro 2 |
| COL4A1_ITGB1 | COL4A1 | ITGB1 | Fibro 2 | Fibro 2 |
| COL6A1_ITGB1 | COL6A1 | ITGB1 | Fibro 2 | Fibro 2 |
| COL6A2_ITGB1 | COL6A2 | ITGB1 | Fibro 2 | Fibro 2 |
| COL6A3_ITGB1 | COL6A3 | ITGB1 | Fibro 2 | Fibro 2 |
| FN1_ITGB1 | FN1 | ITGB1 | Fibro 2 | Fibro 2 |
| HSP90AA1_LRP1 | HSP90AA1 | LRP1 | Fibro 2 | Fibro 2 |
| HSP90B1_LRP1 | HSP90B1 | LRP1 | Fibro 2 | Fibro 2 |
| LAMA2_ITGB1 | LAMA2 | ITGB1 | Fibro 2 | Fibro 2 |
| LAMB1_ITGB1 | LAMB1 | ITGB1 | Fibro 2 | Fibro 2 |
| LGALS1_ITGB1 | LGALS1 | ITGB1 | Fibro 2 | Fibro 2 |
| LPL_LRP1 | LPL | LRP1 | Fibro 2 | Fibro 2 |
| LUM_ITGB1 | LUM | ITGB1 | Fibro 2 | Fibro 2 |
| PSAP_LRP1 | PSAP | LRP1 | Fibro 2 | Fibro 2 |
| S100A1_RYR2 | S100A1 | RYR2 | Fibro 2 | Fibro 2 |
| TIMP1_CD63 | TIMP1 | CD63 | Fibro 2 | Fibro 2 |
| TIMP2_ITGB1 | TIMP2 | ITGB1 | Fibro 2 | Fibro 2 |
| VCAN_ITGB1 | VCAN | ITGB1 | Fibro 2 | Fibro 2 |
| CALM1_RYR2 | CALM1 | RYR2 | Fibro 2 | Fibro 3 |
| COL1A1_CD36 | COL1A1 | CD36 | Fibro 2 | Fibro 3 |
| COL1A1_ITGB1 | COL1A1 | ITGB1 | Fibro 2 | Fibro 3 |
| COL1A2_CD36 | COL1A2 | CD36 | Fibro 2 | Fibro 3 |
| COL1A2_ITGB1 | COL1A2 | ITGB1 | Fibro 2 | Fibro 3 |
| COL3A1_ITGB1 | COL3A1 | ITGB1 | Fibro 2 | Fibro 3 |
| COL4A1_ITGB1 | COL4A1 | ITGB1 | Fibro 2 | Fibro 3 |
| COL6A1_ITGB1 | COL6A1 | ITGB1 | Fibro 2 | Fibro 3 |
| COL6A2_ITGB1 | COL6A2 | ITGB1 | Fibro 2 | Fibro 3 |
| COL6A3_ITGB1 | COL6A3 | ITGB1 | Fibro 2 | Fibro 3 |
| FN1_ITGB1 | FN1 | ITGB1 | Fibro 2 | Fibro 3 |
| LAMA2_ITGB1 | LAMA2 | ITGB1 | Fibro 2 | Fibro 3 |
| LAMB1_ITGB1 | LAMB1 | ITGB1 | Fibro 2 | Fibro 3 |
| LGALS1_ITGB1 | LGALS1 | ITGB1 | Fibro 2 | Fibro 3 |
| LUM_ITGB1 | LUM | ITGB1 | Fibro 2 | Fibro 3 |
| S100A1_RYR2 | S100A1 | RYR2 | Fibro 2 | Fibro 3 |
| TIMP1_CD63 | TIMP1 | CD63 | Fibro 2 | Fibro 3 |
| TIMP2_ITGB1 | TIMP2 | ITGB1 | Fibro 2 | Fibro 3 |
| VCAN_ITGB1 | VCAN | ITGB1 | Fibro 2 | Fibro 3 |
| CALM1_RYR2 | CALM1 | RYR2 | Fibro 2 | Fibro 4 |
| COL1A1_CD36 | COL1A1 | CD36 | Fibro 2 | Fibro 4 |
| COL1A1_ITGB1 | COL1A1 | ITGB1 | Fibro 2 | Fibro 4 |
| COL1A2_CD36 | COL1A2 | CD36 | Fibro 2 | Fibro 4 |
| COL1A2_ITGB1 | COL1A2 | ITGB1 | Fibro 2 | Fibro 4 |
| COL3A1_ITGB1 | COL3A1 | ITGB1 | Fibro 2 | Fibro 4 |
| COL4A1_ITGB1 | COL4A1 | ITGB1 | Fibro 2 | Fibro 4 |
| COL6A1_ITGB1 | COL6A1 | ITGB1 | Fibro 2 | Fibro 4 |
| COL6A2_ITGB1 | COL6A2 | ITGB1 | Fibro 2 | Fibro 4 |
| COL6A3_ITGB1 | COL6A3 | ITGB1 | Fibro 2 | Fibro 4 |
| FN1_ITGB1 | FN1 | ITGB1 | Fibro 2 | Fibro 4 |
| LAMA2_ITGB1 | LAMA2 | ITGB1 | Fibro 2 | Fibro 4 |
| LAMB1_ITGB1 | LAMB1 | ITGB1 | Fibro 2 | Fibro 4 |
| LGALS1_ITGB1 | LGALS1 | ITGB1 | Fibro 2 | Fibro 4 |
| LUM_ITGB1 | LUM | ITGB1 | Fibro 2 | Fibro 4 |
| S100A1_RYR2 | S100A1 | RYR2 | Fibro 2 | Fibro 4 |
| TIMP1_CD63 | TIMP1 | CD63 | Fibro 2 | Fibro 4 |
| TIMP2_ITGB1 | TIMP2 | ITGB1 | Fibro 2 | Fibro 4 |
| VCAN_ITGB1 | VCAN | ITGB1 | Fibro 2 | Fibro 4 |
| CALM1_RYR2 | CALM1 | RYR2 | Fibro 2 | Fibro 5 |
| COL1A1_CD36 | COL1A1 | CD36 | Fibro 2 | Fibro 5 |
| COL1A1_ITGB1 | COL1A1 | ITGB1 | Fibro 2 | Fibro 5 |
| COL1A2_CD36 | COL1A2 | CD36 | Fibro 2 | Fibro 5 |
| COL1A2_ITGB1 | COL1A2 | ITGB1 | Fibro 2 | Fibro 5 |
| COL3A1_ITGB1 | COL3A1 | ITGB1 | Fibro 2 | Fibro 5 |
| COL4A1_ITGB1 | COL4A1 | ITGB1 | Fibro 2 | Fibro 5 |
| COL6A1_ITGB1 | COL6A1 | ITGB1 | Fibro 2 | Fibro 5 |
| COL6A2_ITGB1 | COL6A2 | ITGB1 | Fibro 2 | Fibro 5 |
| COL6A3_ITGB1 | COL6A3 | ITGB1 | Fibro 2 | Fibro 5 |
| FN1_ITGB1 | FN1 | ITGB1 | Fibro 2 | Fibro 5 |
| LAMA2_ITGB1 | LAMA2 | ITGB1 | Fibro 2 | Fibro 5 |
| LAMB1_ITGB1 | LAMB1 | ITGB1 | Fibro 2 | Fibro 5 |
| LGALS1_ITGB1 | LGALS1 | ITGB1 | Fibro 2 | Fibro 5 |
| LUM_ITGB1 | LUM | ITGB1 | Fibro 2 | Fibro 5 |
| S100A1_RYR2 | S100A1 | RYR2 | Fibro 2 | Fibro 5 |
| TIMP1_CD63 | TIMP1 | CD63 | Fibro 2 | Fibro 5 |
| TIMP2_ITGB1 | TIMP2 | ITGB1 | Fibro 2 | Fibro 5 |
| VCAN_ITGB1 | VCAN | ITGB1 | Fibro 2 | Fibro 5 |
| CALM1_RYR2 | CALM1 | RYR2 | Fibro 2 | Fibro 6 |
| COL1A1_CD36 | COL1A1 | CD36 | Fibro 2 | Fibro 6 |
| COL1A1_ITGB1 | COL1A1 | ITGB1 | Fibro 2 | Fibro 6 |
| COL1A2_CD36 | COL1A2 | CD36 | Fibro 2 | Fibro 6 |
| COL1A2_ITGB1 | COL1A2 | ITGB1 | Fibro 2 | Fibro 6 |
| COL3A1_ITGB1 | COL3A1 | ITGB1 | Fibro 2 | Fibro 6 |
| COL4A1_ITGB1 | COL4A1 | ITGB1 | Fibro 2 | Fibro 6 |
| COL6A1_ITGB1 | COL6A1 | ITGB1 | Fibro 2 | Fibro 6 |
| COL6A2_ITGB1 | COL6A2 | ITGB1 | Fibro 2 | Fibro 6 |
| COL6A3_ITGB1 | COL6A3 | ITGB1 | Fibro 2 | Fibro 6 |
| FN1_ITGB1 | FN1 | ITGB1 | Fibro 2 | Fibro 6 |
| LAMA2_ITGB1 | LAMA2 | ITGB1 | Fibro 2 | Fibro 6 |
| LAMB1_ITGB1 | LAMB1 | ITGB1 | Fibro 2 | Fibro 6 |
| LGALS1_ITGB1 | LGALS1 | ITGB1 | Fibro 2 | Fibro 6 |
| LUM_ITGB1 | LUM | ITGB1 | Fibro 2 | Fibro 6 |
| S100A1_RYR2 | S100A1 | RYR2 | Fibro 2 | Fibro 6 |
| TIMP1_CD63 | TIMP1 | CD63 | Fibro 2 | Fibro 6 |
| TIMP2_ITGB1 | TIMP2 | ITGB1 | Fibro 2 | Fibro 6 |
| VCAN_ITGB1 | VCAN | ITGB1 | Fibro 2 | Fibro 6 |
| A2M_LRP1 | A2M | LRP1 | Fibro 5 | Fibro 2 |
| CALM1_RYR2 | CALM1 | RYR2 | Fibro 5 | Fibro 2 |
| FN1_ITGB1 | FN1 | ITGB1 | Fibro 5 | Fibro 2 |
| HSP90AA1_LRP1 | HSP90AA1 | LRP1 | Fibro 5 | Fibro 2 |
| LPL_LRP1 | LPL | LRP1 | Fibro 5 | Fibro 2 |
| LUM_ITGB1 | LUM | ITGB1 | Fibro 5 | Fibro 2 |
| PSAP_LRP1 | PSAP | LRP1 | Fibro 5 | Fibro 2 |
| S100A1_RYR2 | S100A1 | RYR2 | Fibro 5 | Fibro 2 |
| CALM1_RYR2 | CALM1 | RYR2 | Fibro 5 | Fibro 3 |
| FN1_ITGB1 | FN1 | ITGB1 | Fibro 5 | Fibro 3 |
| LUM_ITGB1 | LUM | ITGB1 | Fibro 5 | Fibro 3 |
| S100A1_RYR2 | S100A1 | RYR2 | Fibro 5 | Fibro 3 |
| CALM1_RYR2 | CALM1 | RYR2 | Fibro 5 | Fibro 4 |
| FN1_ITGB1 | FN1 | ITGB1 | Fibro 5 | Fibro 4 |
| LUM_ITGB1 | LUM | ITGB1 | Fibro 5 | Fibro 4 |
| S100A1_RYR2 | S100A1 | RYR2 | Fibro 5 | Fibro 4 |
| CALM1_RYR2 | CALM1 | RYR2 | Fibro 5 | Fibro 5 |
| FN1_ITGB1 | FN1 | ITGB1 | Fibro 5 | Fibro 5 |
| LUM_ITGB1 | LUM | ITGB1 | Fibro 5 | Fibro 5 |
| S100A1_RYR2 | S100A1 | RYR2 | Fibro 5 | Fibro 5 |
| CALM1_RYR2 | CALM1 | RYR2 | Fibro 5 | Fibro 6 |
| FN1_ITGB1 | FN1 | ITGB1 | Fibro 5 | Fibro 6 |
| LUM_ITGB1 | LUM | ITGB1 | Fibro 5 | Fibro 6 |
| S100A1_RYR2 | S100A1 | RYR2 | Fibro 5 | Fibro 6 |

### Supplemental Table 14. Increased Communication From Fibroblast and Cardiomyocyte Subtypes to Cardiomyocyte Cluster 9

| *Normal* | | | | |
|---|---|---|---|---|
| **Pair_Name** | **Ligand** | **Receptor** | **L_cell** | **R_cell** |
| COL1A1_CD36 | COL1A1 | CD36 | Fibro 1 | CM 9 |
| COL1A2_CD36 | COL1A2 | CD36 | Fibro 1 | CM 9 |
| TIMP1_CD63 | TIMP1 | CD63 | Fibro 1 | CM 9 |
| COL1A1_CD36 | COL1A1 | CD36 | Fibro 2 | CM 9 |
| COL1A2_CD36 | COL1A2 | CD36 | Fibro 2 | CM 9 |
| THBS2_CD36 | THBS2 | CD36 | Fibro 2 | CM 9 |
| TIMP1_CD63 | TIMP1 | CD63 | Fibro 2 | CM 9 |
| COL1A2_CD36 | COL1A2 | CD36 | Fibro 3 | CM 9 |
| TIMP1_CD63 | TIMP1 | CD63 | Fibro 3 | CM 9 |
| COL1A1_CD36 | COL1A1 | CD36 | Fibro 4 | CM 9 |
| COL1A2_CD36 | COL1A2 | CD36 | Fibro 4 | CM 9 |
| TIMP1_CD63 | TIMP1 | CD63 | Fibro 4 | CM 9 |
| COL1A1_CD36 | COL1A1 | CD36 | Fibro 5 | CM 9 |
| COL1A2_CD36 | COL1A2 | CD36 | Fibro 5 | CM 9 |
| TIMP1_CD63 | TIMP1 | CD63 | Fibro 5 | CM 9 |
| COL1A2_CD36 | COL1A2 | CD36 | CM 2 | CM 9 |
| TIMP1_CD63 | TIMP1 | CD63 | CM 2 | CM 9 |
| COL1A2_CD36 | COL1A2 | CD36 | CM 3 | CM 9 |
| TIMP1_CD63 | TIMP1 | CD63 | CM 3 | CM 9 |
| CALM1_RYR2 | CALM1 | RYR2 | CM 5 | CM 9 |
| COL1A2_CD36 | COL1A2 | CD36 | CM 5 | CM 9 |
| S100A1_RYR2 | S100A1 | RYR2 | CM 5 | CM 9 |
| TIMP1_CD63 | TIMP1 | CD63 | CM 5 | CM 9 |

| *Non-obstructive* | | | | |
|---|---|---|---|---|
| **Pair_Name** | **Ligand** | **Receptor** | **L_cell** | **R_cell** |
| CALM1_CACNA1C | CALM1 | CACNA1C | Fibro 1 | CM 9 |
| CALM1_PDE1C | CALM1 | PDE1C | Fibro 1 | CM 9 |
| CALM1_RYR2 | CALM1 | RYR2 | Fibro 1 | CM 9 |
| COL1A2_CD36 | COL1A2 | CD36 | Fibro 1 | CM 9 |
| COL1A2_ITGB1 | COL1A2 | ITGB1 | Fibro 1 | CM 9 |
| COL4A1_ITGB1 | COL4A1 | ITGB1 | Fibro 1 | CM 9 |
| COL6A1_ITGB1 | COL6A1 | ITGB1 | Fibro 1 | CM 9 |
| COL6A2_ITGB1 | COL6A2 | ITGB1 | Fibro 1 | CM 9 |
| COL6A3_ITGB1 | COL6A3 | ITGB1 | Fibro 1 | CM 9 |
| FN1_ITGB1 | FN1 | ITGB1 | Fibro 1 | CM 9 |
| LAMA2_ITGB1 | LAMA2 | ITGB1 | Fibro 1 | CM 9 |
| LGALS1_ITGB1 | LGALS1 | ITGB1 | Fibro 1 | CM 9 |
| LUM_ITGB1 | LUM | ITGB1 | Fibro 1 | CM 9 |
| S100A1_RYR2 | S100A1 | RYR2 | Fibro 1 | CM 9 |
| CALM1_CACNA1C | CALM1 | CACNA1C | Fibro 2 | CM 9 |
| CALM1_PDE1C | CALM1 | PDE1C | Fibro 2 | CM 9 |
| CALM1_RYR2 | CALM1 | RYR2 | Fibro 2 | CM 9 |
| CALM2_CACNA1C | CALM2 | CACNA1C | Fibro 2 | CM 9 |
| CALM2_PDE1C | CALM2 | PDE1C | Fibro 2 | CM 9 |
| COL1A1_CD36 | COL1A1 | CD36 | Fibro 2 | CM 9 |
| COL1A1_ITGB1 | COL1A1 | ITGB1 | Fibro 2 | CM 9 |
| COL1A2_CD36 | COL1A2 | CD36 | Fibro 2 | CM 9 |
| COL1A2_ITGB1 | COL1A2 | ITGB1 | Fibro 2 | CM 9 |
| COL3A1_ITGB1 | COL3A1 | ITGB1 | Fibro 2 | CM 9 |
| COL4A1_ITGB1 | COL4A1 | ITGB1 | Fibro 2 | CM 9 |
| COL6A1_ITGB1 | COL6A1 | ITGB1 | Fibro 2 | CM 9 |
| COL6A2_ITGB1 | COL6A2 | ITGB1 | Fibro 2 | CM 9 |
| COL6A3_ITGB1 | COL6A3 | ITGB1 | Fibro 2 | CM 9 |
| FN1_ITGB1 | FN1 | ITGB1 | Fibro 2 | CM 9 |
| LAMA2_ITGB1 | LAMA2 | ITGB1 | Fibro 2 | CM 9 |
| LAMB1_ITGB1 | LAMB1 | ITGB1 | Fibro 2 | CM 9 |
| LGALS1_ITGB1 | LGALS1 | ITGB1 | Fibro 2 | CM 9 |
| LUM_ITGB1 | LUM | ITGB1 | Fibro 2 | CM 9 |
| S100A1_RYR2 | S100A1 | RYR2 | Fibro 2 | CM 9 |
| TIMP1_CD63 | TIMP1 | CD63 | Fibro 2 | CM 9 |
| TIMP2_ITGB1 | TIMP2 | ITGB1 | Fibro 2 | CM 9 |
| VCAN_ITGB1 | VCAN | ITGB1 | Fibro 2 | CM 9 |
| CALM1_CACNA1C | CALM1 | CACNA1C | Fibro 3 | CM 9 |
| CALM1_PDE1C | CALM1 | PDE1C | Fibro 3 | CM 9 |
| CALM1_RYR2 | CALM1 | RYR2 | Fibro 3 | CM 9 |
| COL1A2_CD36 | COL1A2 | CD36 | Fibro 3 | CM 9 |
| COL1A2_ITGB1 | COL1A2 | ITGB1 | Fibro 3 | CM 9 |
| COL3A1_ITGB1 | COL3A1 | ITGB1 | Fibro 3 | CM 9 |
| COL4A1_ITGB1 | COL4A1 | ITGB1 | Fibro 3 | CM 9 |
| COL6A1_ITGB1 | COL6A1 | ITGB1 | Fibro 3 | CM 9 |
| COL6A2_ITGB1 | COL6A2 | ITGB1 | Fibro 3 | CM 9 |
| COL6A3_ITGB1 | COL6A3 | ITGB1 | Fibro 3 | CM 9 |
| FN1_ITGB1 | FN1 | ITGB1 | Fibro 3 | CM 9 |
| LAMA2_ITGB1 | LAMA2 | ITGB1 | Fibro 3 | CM 9 |
| LAMB1_ITGB1 | LAMB1 | ITGB1 | Fibro 3 | CM 9 |
| LGALS1_ITGB1 | LGALS1 | ITGB1 | Fibro 3 | CM 9 |
| LUM_ITGB1 | LUM | ITGB1 | Fibro 3 | CM 9 |
| S100A1_RYR2 | S100A1 | RYR2 | Fibro 3 | CM 9 |
| CALM1_CACNA1C | CALM1 | CACNA1C | Fibro 4 | CM 9 |
| CALM1_PDE1C | CALM1 | PDE1C | Fibro 4 | CM 9 |
| CALM1_RYR2 | CALM1 | RYR2 | Fibro 4 | CM 9 |
| COL1A2_CD36 | COL1A2 | CD36 | Fibro 4 | CM 9 |
| COL1A2_ITGB1 | COL1A2 | ITGB1 | Fibro 4 | CM 9 |
| COL4A1_ITGB1 | COL4A1 | ITGB1 | Fibro 4 | CM 9 |
| COL6A1_ITGB1 | COL6A1 | ITGB1 | Fibro 4 | CM 9 |
| COL6A3_ITGB1 | COL6A3 | ITGB1 | Fibro 4 | CM 9 |
| FN1_ITGB1 | FN1 | ITGB1 | Fibro 4 | CM 9 |
| LGALS1_ITGB1 | LGALS1 | ITGB1 | Fibro 4 | CM 9 |
| LUM_ITGB1 | LUM | ITGB1 | Fibro 4 | CM 9 |
| S100A1_RYR2 | S100A1 | RYR2 | Fibro 4 | CM 9 |
| CALM1_CACNA1C | CALM1 | CACNA1C | Fibro 5 | CM 9 |
| CALM1_PDE1C | CALM1 | PDE1C | Fibro 5 | CM 9 |
| CALM1_RYR2 | CALM1 | RYR2 | Fibro 5 | CM 9 |
| FN1_ITGB1 | FN1 | ITGB1 | Fibro 5 | CM 9 |
| LUM_ITGB1 | LUM | ITGB1 | Fibro 5 | CM 9 |
| S100A1_RYR2 | S100A1 | RYR2 | Fibro 5 | CM 9 |
| CALM1_CACNA1C | CALM1 | CACNA1C | CM 2 | CM 9 |
| CALM1_PDE1C | CALM1 | PDE1C | CM 2 | CM 9 |
| CALM1_RYR2 | CALM1 | RYR2 | CM 2 | CM 9 |
| CALM2_CACNA1C | CALM2 | CACNA1C | CM 2 | CM 9 |
| CALM2_PDE1C | CALM2 | PDE1C | CM 2 | CM 9 |
| COL1A2_CD36 | COL1A2 | CD36 | CM 2 | CM 9 |
| COL1A2_ITGB1 | COL1A2 | ITGB1 | CM 2 | CM 9 |
| COL6A2_ITGB1 | COL6A2 | ITGB1 | CM 2 | CM 9 |
| FN1_ITGB1 | FN1 | ITGB1 | CM 2 | CM 9 |
| LGALS1_ITGB1 | LGALS1 | ITGB1 | CM 2 | CM 9 |
| LUM_ITGB1 | LUM | ITGB1 | CM 2 | CM 9 |
| S100A1_RYR2 | S100A1 | RYR2 | CM 2 | CM 9 |
| TIMP1_CD63 | TIMP1 | CD63 | CM 2 | CM 9 |
| CALM1_CACNA1C | CALM1 | CACNA1C | CM 3 | CM 9 |
| CALM1_PDE1C | CALM1 | PDE1C | CM 3 | CM 9 |
| CALM1_RYR2 | CALM1 | RYR2 | CM 3 | CM 9 |
| CALM2_CACNA1C | CALM2 | CACNA1C | CM 3 | CM 9 |
| CALM2_PDE1C | CALM2 | PDE1C | CM 3 | CM 9 |
| LAMA2_ITGB1 | LAMA2 | ITGB1 | CM 3 | CM 9 |
| LGALS1_ITGB1 | LGALS1 | ITGB1 | CM 3 | CM 9 |
| NCAM1_CACNA1C | NCAM1 | CACNA1C | CM 3 | CM 9 |
| S100A1_RYR2 | S100A1 | RYR2 | CM 3 | CM 9 |
| TGM2_ITGB1 | TGM2 | ITGB1 | CM 3 | CM 9 |
| VEGFA_ITGB1 | VEGFA | ITGB1 | CM 3 | CM 9 |
| CALM1_CACNA1C | CALM1 | CACNA1C | CM 5 | CM 9 |
| CALM1_PDE1C | CALM1 | PDE1C | CM 5 | CM 9 |
| CALM1_RYR2 | CALM1 | RYR2 | CM 5 | CM 9 |
| CALM2_CACNA1C | CALM2 | CACNA1C | CM 5 | CM 9 |
| CALM2_PDE1C | CALM2 | PDE1C | CM 5 | CM 9 |
| CALM3_CACNA1C | CALM3 | CACNA1C | CM 5 | CM 9 |
| CALM3_PDE1C | CALM3 | PDE1C | CM 5 | CM 9 |
| CALM3_RYR2 | CALM3 | RYR2 | CM 5 | CM 9 |
| COL1A2_CD36 | COL1A2 | CD36 | CM 5 | CM 9 |
| COL1A2_ITGB1 | COL1A2 | ITGB1 | CM 5 | CM 9 |
| COL6A1_ITGB1 | COL6A1 | ITGB1 | CM 5 | CM 9 |
| COL6A2_ITGB1 | COL6A2 | ITGB1 | CM 5 | CM 9 |
| FN1_ITGB1 | FN1 | ITGB1 | CM 5 | CM 9 |
| LAMB2_ITGB1 | LAMB2 | ITGB1 | CM 5 | CM 9 |
| LGALS1_ITGB1 | LGALS1 | ITGB1 | CM 5 | CM 9 |
| LGALS3BP_ITGB1 | LGALS3BP | ITGB1 | CM 5 | CM 9 |
| LUM_ITGB1 | LUM | ITGB1 | CM 5 | CM 9 |
| S100A1_RYR2 | S100A1 | RYR2 | CM 5 | CM 9 |
| TGM2_ITGB1 | TGM2 | ITGB1 | CM 5 | CM 9 |
| TIMP1_CD63 | TIMP1 | CD63 | CM 5 | CM 9 |
